# Ophthalmic imaging as a measure of cardiovascular and neurological health: a multi-omic analysis of deep-learning derived phenotypes

**DOI:** 10.1101/2025.08.04.25332962

**Authors:** Thomas H. Julian, Haoran Dou, Jinming Duan, Jinghan Huang, Esther Yoo, David J. Green, Andrew Strange, Elham Alhathli, UK Biobank Eye and Vision Consortium, Matthew Sperrin, Pearse A Keane, Emily Y. Chew, Bernard Keavney, Tomas W. Fitzgerald, Johnathan Cooper-Knock, Ewan Birney, Alejandro F. Frangi, Panagiotis I. Sergouniotis

## Abstract

The eye is a recognised source of biomarkers for cardiovascular and neurodegenerative disease risk. Here, we characterise the breadth of these associations and identify biological axes that may mediate them. Using UK Biobank data, we developed a multi-omic analysis pipeline integrating physiological, radiomic, metabolomic, and genomic information. We trained adversarial autoencoders (Ret-AAE) to represent optical coherence tomography (OCT) images and colour fundus photographs as 256-dimensional embeddings. Ret-AAE derived embeddings were associated with a range of cardiovascular and neurodegenerative diseases, including ischaemic heart disease, cerebrovascular disease, Parkinson’s disease, and dementia. Examining associations across diverse omics datasets, we provide evidence linking ophthalmic imaging features to neurological and cardiovascular anatomy and function, lipid metabolism, and gene sets associated with neurodegenerative pathology. Collectively, our findings demonstrate that ophthalmic features reflect complex, multisystem biological processes, and reinforce the role of the eye as a composite indicator of systemic health.

## Introduction

The eye is uniquely positioned among human organs in that it permits direct, non-invasive visualisation of both the vasculature and the central nervous system.^1–3^ Given this property, ophthalmic structures have long been recognised as potential indicators of systemic health, particularly in relation to neurodegenerative and cardiovascular disease.

In clinical practice, this relationship is well established and materially important. Ophthalmic findings often prompt investigations into systemic conditions: retinal features may lead to assessment of blood pressure or diabetic status; papilledema to neuroimaging; and signs of ocular ischemia to evaluation of carotid artery patency.^4^ These associations, however, reflect overt disease, *i.e.* manifestations that are already visible to the human eye and reflect established disease.

The role of the eye in public health is now increasingly recognised as being significantly broader than previously understood. Several developments have elevated interest in the field: (i) advances in ophthalmic imaging technologies have offered better resolution of disease-relevant tissues ^5^; (ii) retinal imaging is becoming increasingly widespread - with modalities such as colour fundus photography (CFP) and optical coherence tomography (OCT) now available not only in specialist eye clinics but also in many optometry practices and community-based healthcare settings; and (iii) machine learning models have become increasingly effective at representing complex structures in medical imaging data.^6^ Inspired by these advances, deep learning models utilising ophthalmic imaging - developed by our group and others - have been applied to predict a range of neurodegenerative and cardiovascular conditions including myocardial infarction, heart failure, Alzheimer’s disease, stroke, and Parkinson’s disease.^2,7–11^

Despite significant progress in this area, several key challenges remain. These include: providing explainability of the relationship between the eye, heart and brain; determining the breadth of disease that can be predicted; incorporating multi-modal data in models; integrating temporally embedded, sequential imaging data to enable modelling of longitudinal health trajectories; improving model validity across diverse populations; exploring the cost effectiveness of these technologies for primary prevention; and solving logistical technology implementation challenges required for deployment. In this study, we address two of these challenges: (1) delineating the spectrum of neurodegenerative/cardiovascular diseases that are linked to ophthalmic imaging features; and (2) characterising the aberrant biological processes detectable in ophthalmic images that may be relevant to systemic disease risk. Previous work has principally focussed on exploring the mechanisms linking ophthalmic features to systemic disease using a narrow range of expert-defined imaging traits, such as OCT layer thicknesses, and vessel widths and tortuosity measures in CFPs. This approach leads to repeated study of features we *already* know are important and may neglect more complex or abstract structural features that are challenging to define using human labels. Accordingly, we introduce **Ret-AAE** (retinal adversarial autoencoder), an autoencoder framework that compresses OCT and CFP images into 256-dimensional vector embeddings (hereafter, ‘Ret-AAE embeddings’), allowing exploration of oculo-systemic associations in a manner free of prior assumptions.

To address challenge (1), we investigated the breadth of cardiovascular and neurodegenerative disease associations with ophthalmic features using two complementary analyses: (a) exploration of the association between multimodality Ret-AAE embeddings and baseline neurodegenerative/cardiovascular conditions, where variations likely reflect the presence of diagnosed disease or its associated systemic correlates; and (b) prospective associations between multimodality Ret-AAE embeddings and incident disease, establishing the extent to which retinal features may serve as early indicators of future pathology. With a view to addressing challenge (2), we explored the plethora of factors that may mediate the link between retinal imaging features and neurological/cardiovascular health, generating mechanistic hypotheses for further validation. To this end, we developed an hypothesis-free multi-omic pipeline leveraging UK Biobank (UKB) data, integrating physiological, functional, radiomic, metabolomic, and genomic data to interrogate the architecture of these oculo-systemic associations (**Figure 1**).

**Figure 1:**
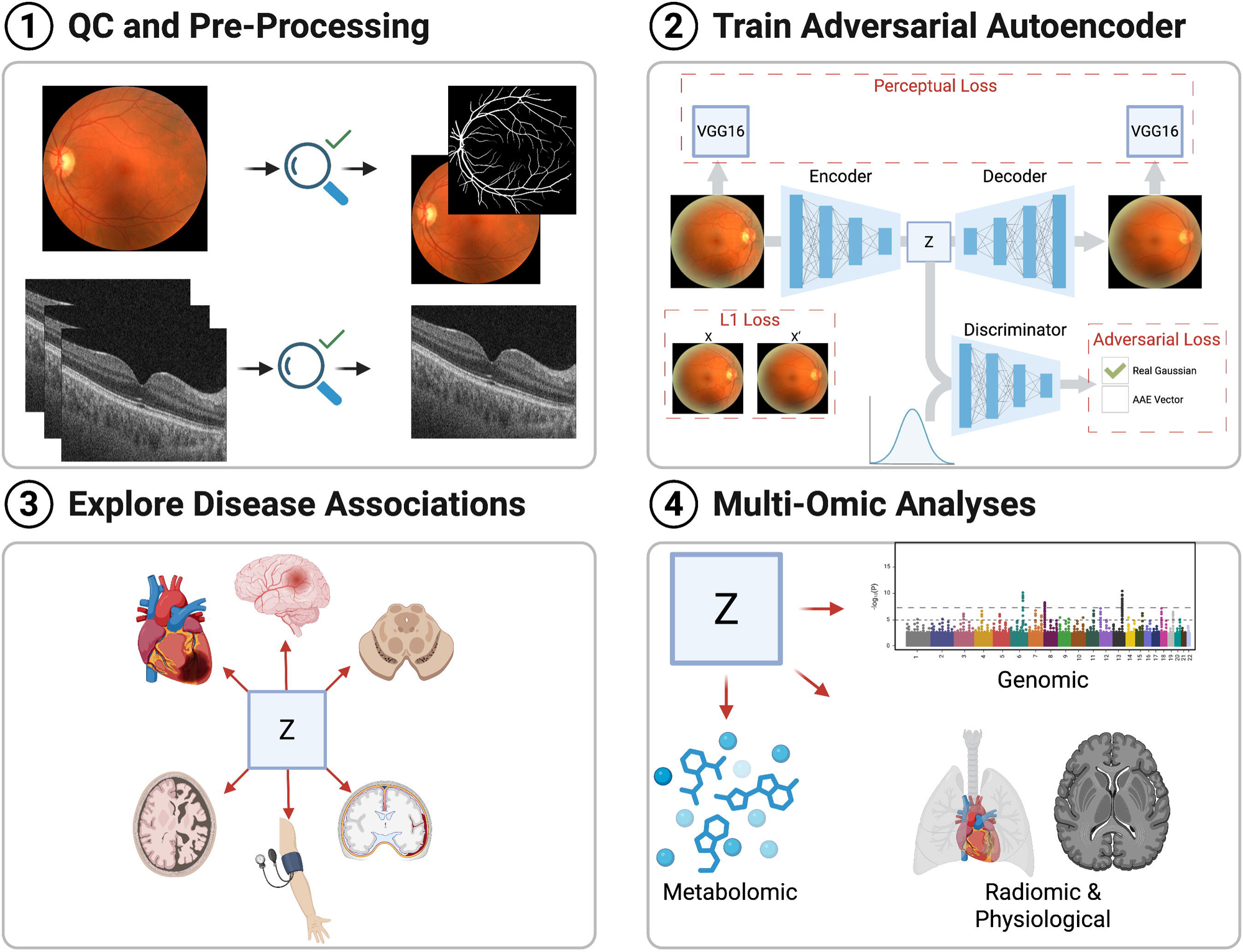
An overview of the four key components to this analysis. **(1)** Data is quality controlled prior to analysis. **(2)** The optical coherence tomography and colour fundus photograph images are then used to train an adversarial autoencoder. **(3-4)** The latent features (‘embeddings’) projected by the encoder (denoted *z*) are then utilised to (a) explore ophthalmic associations with disease, and (b) study potential biological and genetic mechanisms that link ophthalmic features to systemic disease.

## Results

### Ret-AAE maps ophthalmic images to a low-dimensional vector whilst preserving critical anatomical features

After quality control, a total of 63,946 images were available to train the CFP-AAE (13,570 and 14,024 for test and validation respectively). Further, a total of 88,972 images were available to train the OCT-AAE (19,018 and 19,621 for test and validation, respectively). The structural similarity index metric (SSIM) for CFP reconstructions was 0.89 and the peak signal to noise ratio (PSNR) was 30.65 dB. The SSIM for the OCT images was 0.72 whilst PSNR was 29.19 dB. The speckled (noisy) nature of OCT images was not preserved during reconstruction and images were smoothed, which may account for the lower reconstruction metrics relative to CFPs despite perceptually superior anatomical structure preservation. Eight randomly sampled example reconstructions for OCT and CFPs are presented in **Supplementary Figures 1-2**. Most ophthalmic imaging embeddings (latent features) approximated a Gaussian distribution (**Supplementary Figures 3-4**).

### Population characteristics of those with sufficiently high-quality images for phenotypic and genotypic analysis

Phenotypic and genotypic associations were conducted in subjects with sufficiently high-quality left eye CFP (N=42,001) or OCT (N=68,942) images, although the total number of subjects for each analysis varied from test to test due to missingness. The overall cohort with such images were significantly younger than those with no ophthalmic imaging (Day one of the imaging study median ages: OCT imaging = 58, CFP imaging = 55, no imaging = 59, Kruskal-Wallis *p*-value ≈ 0), differed in terms of genetic sex (OCT imaging = 52.8% female, CFP imaging = 54.8% female, no imaging = 54.4% female, χ2 *p*-value=2.62E-14); were less likely to identify as White ethnicity (OCT imaging = 92%, CFP imaging = 94%, no imaging = 96%, χ2 *p*-value ≈ 0), and were less wealthy (median Townsend score for OCT = −1.84, CFP = −1.89, no imaging = −2.27, Kruskal-Wallis *p*-value = 1.83E-175). Visualisations of the population characteristics are presented in **Supplementary Figures 5-8**.

### Ret-AAE embeddings are associated with ophthalmic anatomy, function, genetics and disease

We performed three ‘positive control’ studies to explore the representation of retinal features within Ret-AAE embeddings (denoted ‘*z_i_’,* where *i* is the relevant latent dimension): (i) Pearson correlation analysis to assess associations between ophthalmic imaging embeddings and ophthalmic anatomical or functional features; (ii) Welch’s t-test to compare ophthalmic imaging embeddings based on the presence or absence of ophthalmic disease; and (iii) Cox proportional hazard modelling to examine whether the embeddings predict future ophthalmic disease. Additionally, expecting to reproduce the findings of genome wide-association studies (GWAS) for conventional ophthalmic imaging traits, we conducted GWAS of the 512 Ret-AAE embeddings.

These ‘positive control’ studies yielded results demonstrating that our models effectively capture and represent key ophthalmic features. See the “controlling for type one error” section of this manuscript for the details of our approach to multiple comparisons.

The Pearson correlation study of latent features against 50 ophthalmic traits (e.g. conventional ophthalmic imaging-derived traits, visual acuity, refractive error, intraocular pressure, and corneal biomechanics). Reassuringly, we detected extensive multiple-testing corrected significant associations between Ret-AAE embeddings and the breadth of ophthalmic features analysed (**Supplementary Tables 1.1-1.3**, **Supplementary Figures 9-11)**. Exploring the association between Ret-AAE embeddings and baseline ophthalmic disease, we revealed that Ret-AEE embeddings are associated with all ophthalmic diseases tested (**Supplementary Figures 12-13).** The largest number of associations exceeding a Bonferroni significance threshold were observed for cataracts, vitreous disorders, glaucoma, retinal detachments and breaks, and other retinal disorders. Additionally, we found that CFP- and OCT-derived embeddings predicted post-imaging ophthalmic disease, including disorders of the lens, choroid, retina and vitreous (**Supplementary Figures 14-15**). Complete positive control analyses are available in **Supplementary Tables 1.2-1.7**.

With a view to exploring the biology captured by Ret-AAE embeddings, we conducted a series of genetic analyses. The OCT-derived embedding discovery GWAS contained 30,863 subjects, with replication in 6,847 subjects (see Methods). The CFP-derived embedding discovery GWAS included 19,109 individuals, with replication in 4,402 subjects. These GWAS detected significant associations with numerous genetic variants identified in studies of conventional ophthalmic imaging traits and revealed previously unreported variants of interest.

Following fine-mapping (GCTA COJO), the GWAS of the 256 OCT-derived embeddings highlighted genome-wide significant associations for 33 different single nucleotide variants across 39 embeddings. ^12^ Of these, 9 replicated in the validation study (across 13 embeddings, **Supplementary Figure 16**, **Supplementary Table 1.8**). Several GWAS hits are implicated in pigmentation, including *TYR*, *OCA2*, *DCT*, and *TSPAN10*.^13–16^ Additionally, we identified significant signals in genes with core roles in retinal development/physiology, namely *RDH5* (which encodes an enzyme with roles in the visual cycle) and *VSX2* (a retinal transcription factor). Most detected variants have been previously identified in conventional ophthalmic trait GWAS, with the exception of *OSTF1* (a protein produced by osteoclasts, which has previously been linked to retinitis pigmentosa risk) and *DCT* (previously linked to albinism).^17,18^ Linkage disequilibrium (LD) score regression indicated that seven (2.73%) OCT-derived embeddings had a polygenic architecture, with a heritable component (range = 0.39% - 5.13%, median = 3.46%), which may reflect statistical power, multifactorial inheritance patterns, environmental influence, technical factors, and/or that some embeddings are distributed according to right eye features (note the GWAS was conducted using left eye features).

GWAS of the 256 CFP-derived embeddings identified 51 independent genome-wide significant variants across 80 embeddings. Of these, 13 replicated in the validation study (across 53 embeddings) (**Supplementary Table 1.9**). Again, several of these variants were in genes with key roles in pigmentation (*TYR*, *OCA2*, *DCT,* and *IRF4*)^19^. Additionally, we identified a vasculature-associated signal in *PDE3A.* This gene encodes a protein that regulates vascular smooth muscle contraction and relaxation, which was previously linked to retinal vasculature features, aortic root size, and blood pressure.^20^ There were novel structural imaging-genetics associations with *TMCC2,* (a gene associated with machine learning derived age-gap measures), *RXFP3* (a gene associated with aging-related disorders), and *IL1RAP* (a component of the interleukin 1 receptor complex, associated with ocular disease).^21–23^ Overall, 23 embeddings (8.98%) had a polygenic architecture with the heritability of polygenic CFP-derived embeddings exceeding that of OCTs, and ranging from 3.12% to 15.5% (median = 5.75%).

Gene set analysis using Multi-marker Analysis of GenoMic Annotation (MAGMA) revealed 33 multiple-testing corrected significant gene sets with associations spanning 25 CFP-derived embeddings; and 26 gene sets with associations spanning 22 OCT-derived embeddings.^24^ Of these, four CFP- and six OCT-associated gene sets replicated in the validation study. For OCT-derived embeddings, replicated gene sets included ‘KEGG MEDICUS variant duplication or mutation activated Fms-like tyrosine kinase 3 (FLT3) to Jak-STAT signaling pathway’, associated with a choroid localised embedding (*z_153_*, beta = 1.71, *p*-value = 6.00E-06); ‘KEGG MEDICUS reference regulation of complement cascade *CFHR’*, associated with a choroid/retinal pigment epithelium localised embedding (*z_126_*, beta = 2.23, *p*-value = 9.00E-06); and ‘WikiPathways (WP) FOXP3 in COVID19’, also associated with a choroid localised embedding (*z_153_*, beta = 0.86, *p-*value = 9.00E-06). The gene sets identified fit with our understanding of the importance of pigmentation, the complement system, and fibroblast growth factor receptors in ophthalmic biology; as well as revealing new pathways warranting further exploration in dedicated studies.^15,25^ For CFP-derived embeddings, the replicated pathways included ‘Reactome melanin biosynthesis’, which was associated with fundal background and optic nerve head localised embeddings (*z_142_*, and *z_148_*, beta = 1.86-2.27, *p*-value = 5.56E-10 - 1.81E-07), and ‘Reactome phospholipase C mediated cascade fibroblast growth factor receptor 2 (FGFR2*)’,* associated with a vasculature localised embedding (*z_173_*, beta 0.9, *p-*value = 6.66E-06). Cluster maps of MAGMA results are presented in **Supplementary Figures 17-18** and the complete results are presented in **Supplementary Tables 1.10-1.11**. Further Ret-AAE embedding-associated pathways with relevance to neurodegenerative disease were identified, and are discussed in the relevant section later.

Collectively, our GWAS findings illustrate that Ret-AAE embeddings are associated with established ophthalmic disease-related genes as well as genes associated with biological age discrepancies, pigmentation, and vascular physiology. The complete GCTA COJO results can be viewed in **Supplementary Table 1.12-1.13**. Heritability analyses are presented in **Supplementary Tables 1.14-1.15**.

### Associations between ophthalmic imaging features and cardiovascular disease

CFP- and OCT-derived embeddings were associated with numerous cardiovascular diseases at baseline (i.e., at the time of first ophthalmic imaging), following correction for age and genetic sex (**Supplementary Table 1.16-1.17**). The largest number of significant associations were seen for ischaemic heart disease and hypertension (**Figure 2)**. A smaller number of significant associations were seen for cerebrovascular disease and heart failure. There were no significant associations between Ret-AAE and cardiomyopathy. Gradient□weighted Class Activation Mapping (Grad-CAM, **Supplementary Figures 19-48**) analyses suggested that cardiovascular disease-associated embeddings (e.g. CFP *z_9_, z_20_, z_63_, z_148_, z_154_, z_218_*, OCT *z_173_, z_209_, z_218_*) localised to the retinal vasculature, the optic nerve head, the choroidal layer, neurosensory retina, and background features (potentially pigmentation or choroidal features).

**Figure 2:**
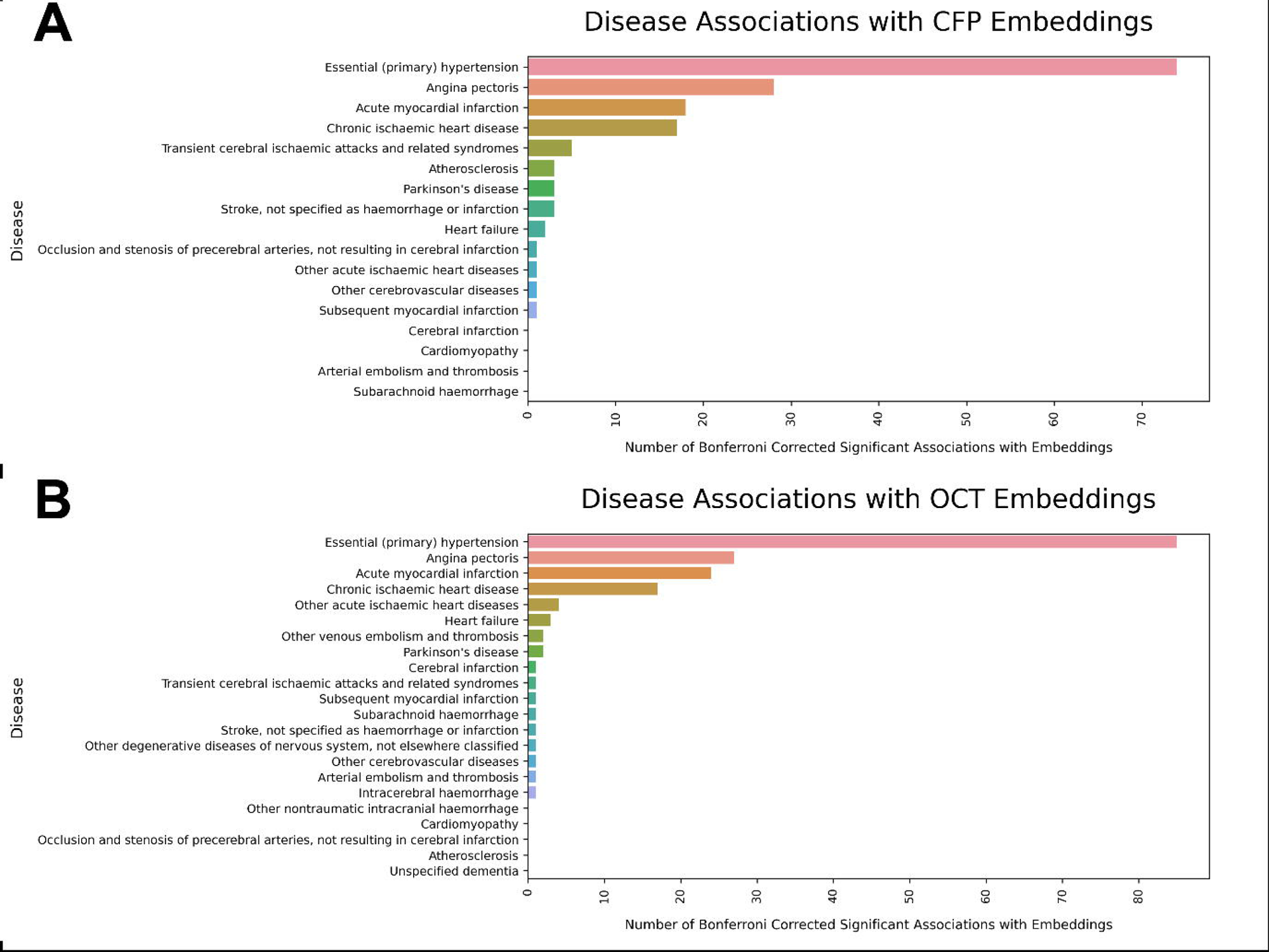
A bar plot showing the number of multiple-testing corrected significant associations between **(A)** CFP-derived & **(B)** OCT-derived embeddings and neurodegenerative/ cardiovascular diseases at the time of baseline imaging. The individual p values are presented in the supplementary tables and can be referred to in order to appraise specific tests and the strength of significance.

CFP- and OCT-derived embeddings were predictive of future risk of a wide range of cardiovascular diseases including hypertension, stroke, angina pectoris, ischaemic heart disease, and heart failure (**Figure 3, Supplementary Tables 1.18-1.19)**. Although essential hypertension was associated with numerous CFP-derived embeddings localising to the vasculature, optic nerve head, and fundal background (e.g. *z_6_*, *z_148_*, and *z_208_*), it was also associated with just one OCT-derived embedding, which appeared to localise to the borders of the macula (*z_105_*). Amongst OCT-derived embeddings, heart failure emerged as the most frequently associated disorder. This was most strongly associated with embeddings localising to the ellipsoid zone, retinal pigment epithelium, ganglion cell layer, retinal nerve fibre layer, and to some extent the choroid (*z_209_* and *z_218_*). The contrasting disease prediction profiles of CFP- and OCT-derived embeddings brings attention to the value that multi-modal disease prediction models may hold as a means of measuring diverse cardiovascular health outcomes.

**Figure 3:**
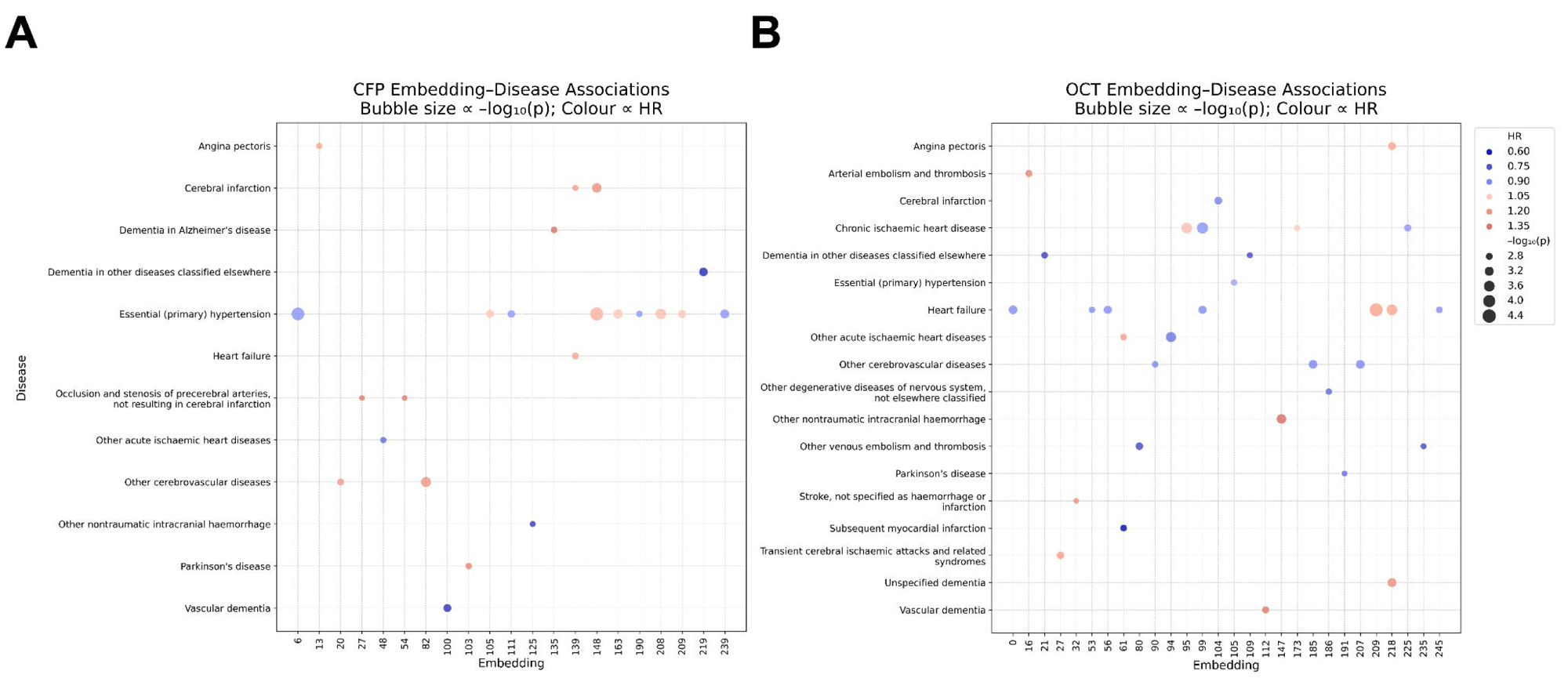
A bubble plot illustrating the results of our CFP-derived **(A)** and OCT-derived **(B)** embedding Cox proportional hazards analysis. Only embeddings and disorders with at least one multiple testing corrected significant result are plotted here. The size of the bubbles indicates the level of significance (larger bubbles = greater significance, unadjusted). The bubbles are colour coded according to the hazards ratio (HR). The individual p values are presented in the supplementary tables and can be referred to in order to appraise specific tests and the strength of significance.

Using LD score regression, we estimated the extent of genetic correlation between Ret-AAE embeddings and cardiovascular disease. This analysis has substantial limitations, given that it can only be conducted on the minority of embeddings with a polygenic genetic architecture and only for traits for which there are well-powered, publicly available GWAS summary statistics. Accordingly, genetic correlation analyses were undertaken against angina, myocardial infarction, heart failure, stroke, and systolic blood pressure. We found a few CFP embeddings that were at least nominally associated with cardiovascular disease. All of these localised to the vascular tree and included: embedding *z_220_*, which was moderately genetically correlated with myocardial infarction after multiple testing correction (r_xy_ = 0.38, *p*-value = 0.003); embedding *z_72_* which was nominally significantly genetically correlated with myocardial infarction (r_xy_ = 0.22, *p*-value = 0.01); and embeddings *z_68_* and *z_111_*, which were nominally genetically correlated with heart failure (r_xy_ = −0.32 and 0.34, *p*-value = 0.04 and 0.04, respectively). These correlations offer some evidence that there is genetic overlap between ophthalmic features and cardiovascular disease. The complete LD score regression results are available in **Supplementary Tables 1.20-1.21**.

Next, we sought to explore how Ret-AEE embeddings relate to cardiovascular anatomy, physiology, and function. To that end, we calculated the Pearson correlation coefficient of these embeddings against cardiovascular traits. HDBSCAN clustering revealed four cardiovascular trait groups (**Supplementary Table 1.22**). ‘Cluster 0’ was comprised of electrocardiogram (ECG) intervals (‘PP interval’, ‘RR interval’, ‘QT interval’); ‘cluster 1’ grouped heart rate measures; ‘cluster 2’ included central pulse pressure, peripheral pulse pressure, and stroke volume; and ‘cluster 3’ encompassed blood pressure measures (**Supplementary Figure 49**). Associations were of low magnitude but high significance (CFP: 0.06 > r_xy_ > −0.12; OCT: 0.08 > r_xy_ > −0.20, excluding small sample sizes). The embeddings were principally associated with ‘noise’ defined features (those which do not fit into a cluster, coded −1), ‘cluster 2’ features, and ‘cluster 3’ features (**Figure 4)**. The significant ‘noise’ results were led by pulse wave arterial stiffness index, pulse wave reflection index, and blood pressure measurements. Notable non-clustered associations included body surface area, left ventricular stroke volume, end-diastolic volume, and end-systolic volumes. Overall, 24 cardiovascular traits were associated with at least one CFP-derived embedding, and 21 with OCT-derived embeddings. The embeddings with the most cardiovascular associations were vascular traits (OCT-embedding *z_199_*, choroid localised; and CFP-embedding *z_106_*, vascular tree localised). The traits with the most significant associations with embeddings are presented in **Supplementary Figures 50-51**. For the full breadth of results see **Supplementary Tables 1.23-1.24**.

**Figure 4:**
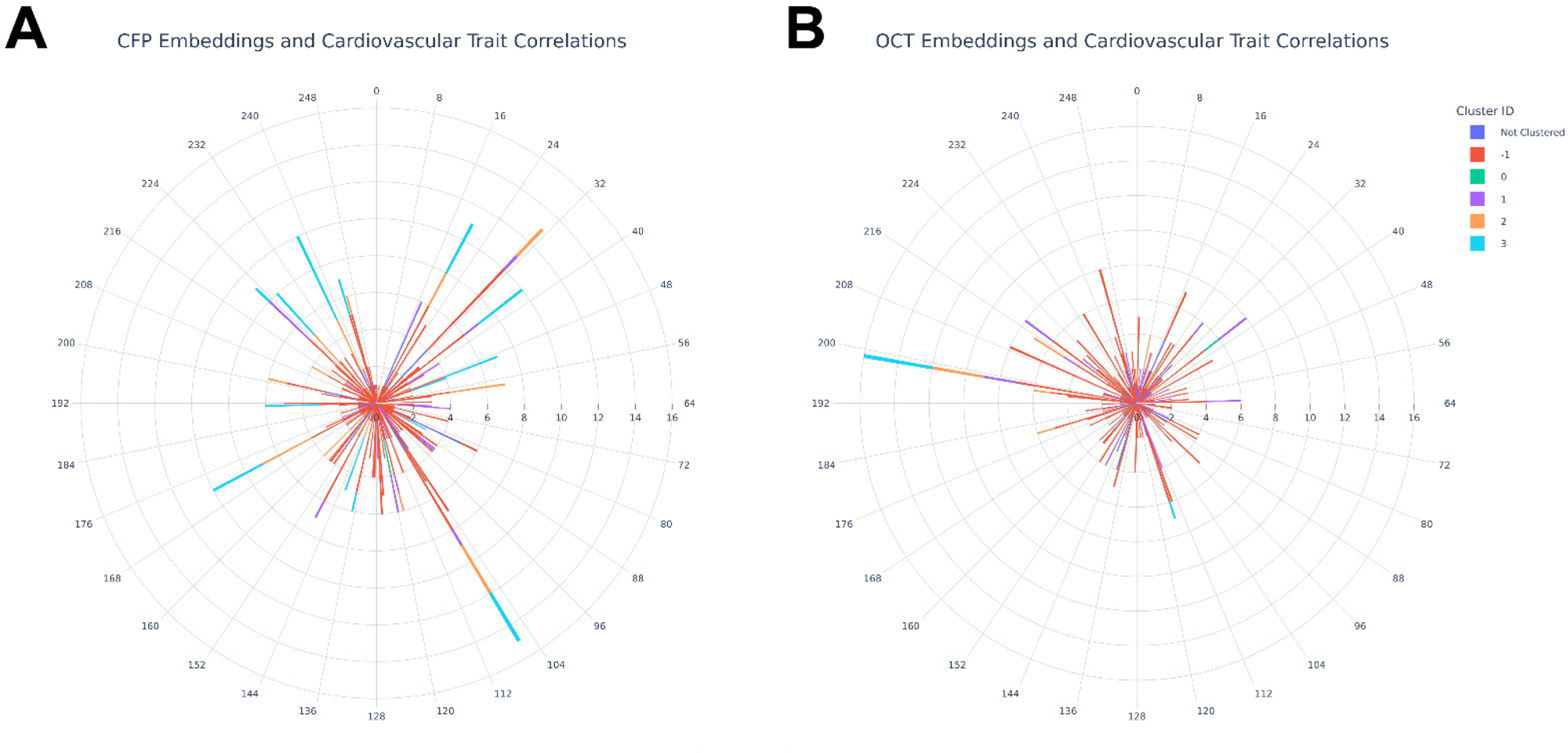
Polar plots displaying the number of significant relationships (corrected for multiple comparisons) between **(A)** CFP-derived embeddings, **(B)** OCT-derived embeddings, and cardiovascular traits. The circumferential axis indicates the embedding number. The radial axis indicates the number of multiple testing corrected significant Pearson correlations that the embedding has with cardiovascular traits. The bars are colour coded according to the cluster of traits the significant relationship belongs to. ‘Cluster 0’ was comprised of electrocardiogram (ECG) intervals (‘PP interval’, ‘RR interval’, ‘QT interval’); ‘cluster 1’ grouped heart rate measures; ‘cluster 2’ included central pulse pressure, peripheral pulse pressure, and stroke volume; and ‘cluster 3’ encompassed blood pressure measures. The individual p values are presented in the supplementary tables and can be referred to for appraisal of specific tests and the strength of significance. HDBSCAN clustering has been used to simplify the presentation of results by grouping traits into statistically defined categories, but readers may wish to explore the more granular trait-wise results in the supplementary content.

To identify ophthalmic imaging associated metabolomic signatures which may be of relevance to the association between ophthalmic anatomy and systemic disease, we performed canonical correlation analysis (CCA) between Ret-AAE embeddings and 251 circulating metabolites. The CFP-metabolomic CCA included 16,917 subjects whilst the OCT-metabolomic analysis included 27,849 individuals. For CFP-derived embeddings, two canonical modes were retained following inspection of the Scree plot (**Supplementary Figures 52-54**). Both modes demonstrated moderate correlation between CFP-derived embeddings and metabolomic traits, with the analyses dominated by high density lipoprotein (HDL) and low-density lipoprotein (LDL) compositional measures (Mode 1: r = 0.44, permutation *p*-value < 0.001; Mode 2: (r = 0.39, *p*-value < 0.001). For the OCT embedding–metabolomics CCA, the first canonical mode was evaluated via permutation testing (**Supplementary Figure 55-56**). This revealed moderate correlation (r = 0.36, *p*-value < 0.001), and was again dominated by HDL, LDL, and very low-density lipoprotein (VLDL)-related features. Notably, ‘cholesterol in large HDL’ had the highest CCA weighting by a substantial margin. The complete results of CCA are presented in **Supplementary Tables 1.25-1.27**. Seeking to validate our results under statistical assumptions that allow for high correlation amongst metabolomic data, we conducted a further embedding-metabolic association study using canonical sparse partial least squares (SPLS). SPLS re-affirmed the association between CFP embeddings, and HDL and LDL compositional measures; whilst SPLS for OCT embeddings recapitulated associations between embeddings and HDL, LDL and VLDL-related metabolites, whilst also indicating a substantial contribution of creatinine to the model which was not identified in CCA. Although we did not explore the relationship with renal parameters in this study, the creatinine feature importance is in keeping with a recognised link between OCT features and renal health. SPLS hyperparameter testing, loading, and cluster plots are presented in **Supplementary Figures 57-64**.^26^ These results suggest that anatomical features represented by Ret-AAE embeddings are associated with lipid metabolism. Given the recognised centralism of lipid metabolism in cardiovascular disease, these findings highlight circulating lipoprotein composition as a shared biological axis between ophthalmic structure and systemic cardiometabolic health.^27,28^

### Associations between ophthalmic imaging features and neurodegenerative disease

Ret-AAE derived embeddings were associated with Parkinson’s disease at baseline imaging, at a level exceeding a multiple comparisons-corrected threshold (**Supplementary Tables 1.16-1.17)**. The embeddings most strongly associated with Parkinson’s disease (e.g. CFP *z_148_*, OCT *z_15_*) localised to the optic nerve head and ellipsoid zone (photoreceptor layer).

Ret-AAE embeddings were also associated with future risk of several neurodegenerative diseases, after correction for age and genetic sex. Both CFP- and OCT-derived embeddings were predictive of future risk of Parkinson’s disease, vascular dementia, and dementia in other diseases (inclusive of Pick disease, Creutzfeldt-Jakob disease, Parkinson’s disease, Huntington disease, human immunodeficiency virus and others). It is noted that Alzheimer’s dementia was associated with CFP-derived embeddings (e.g. vasculature and optic nerve head localised CFP-*z_103_*), but not OCT-derived embeddings.

The LD score regression analysis was highly limited by the public availability of sufficiently polygenic neurodegenerative disease GWAS. However, it was possible to explore the genetic relationship between Alzheimer’s disease and polygenic Ret-AAE embeddings - finding that there was no evidence of significant genetic correlation. Conversely, our gene set analysis revealed interesting relationships between Ret-AAE embeddings and four pathways associated with neurodegenerative disorders. CFP-derived embeddings were associated with ‘WP kynurenine pathway and links to cell senescence’ (*z_41_*, an embedding localising to numerous CFP features, beta = 0.71, p value = 8.31E-06). OCT-derived embeddings were associated with the ‘Pathway Interaction Database alpha synuclein pathway’ (*z_135_*, loosely localising to outer retina, beta = 0.74, p value = 1.00E-06), KEGG MEDICUS variant mutation-caused aberrant amyloid beta to VGCC-Ca^2+^ apoptotic pathway N01006 (*z_36_*, not localisable, beta = 1.25, *p*-value = 1.10E-05), and KEGG MEDICUS variant mutation caused aberrant amyloid beta to transport of calcium (*z_36_*, beta = 1.02, *p*-value = 1.10E-05).

HDBSCAN clustering identified four groups of neurological traits. ‘Cluster 0’ was principally constituted of grey matter volumes assessed using magnetic resonance imaging (MRI); ‘clusters 1-3’ were largely constituted of diffusion MRI values (**Supplementary Figure 65, Supplementary Table 1.28**). This analysis also revealed extensive low magnitude, but highly statistically significant correlations distributed across a small number of embeddings (CFP: 0.11 > r_xy_ > −0.12; OCT: 0.08 > r_xy_ > −0.23, excluding small sample sizes). The analysis was dominated by significant results in diffusion MRI dominated ‘clusters 2-3’, with a smaller proportion of multiple testing significant associations with the grey matter volume dominated MRI ‘cluster 0’ (**Figure 5)**. Key significant results included OCT-derived embedding *z_209_* (principally localised to the inner retinal layers, ellipsoid zone, and RPE; 222 multiple-testing corrected significant results), *z_215_* (poor localisation; 239 multiple-testing corrected significant results), and *z_218_*(retinal nerve fibre layer localised; 255 multiple-testing corrected significant results); and for CFP-derived embedding *z_148_* (optic nerve head, background feature, and projection artifact localised; 189 multiple-testing corrected significant associations). OCT-derived embeddings had substantially more significant associations with neurological traits than those from CFPs. Fluid intelligence score (the capacity to solve problems that require logic and reasoning ability, independent of acquired knowledge) and reaction times showed the largest number of significant associations with imaging latent features, followed by cerebral volume measures. **Supplementary Figures 66-67** illustrate the traits with the largest number of significant associations with embeddings. Overall, 459 neurological traits had significant associations with at least one OCT-derived embedding, and 390 with at least one CFP-derived embedding (**Supplementary Tables 1.29-1.30**).

**Figure 5:**
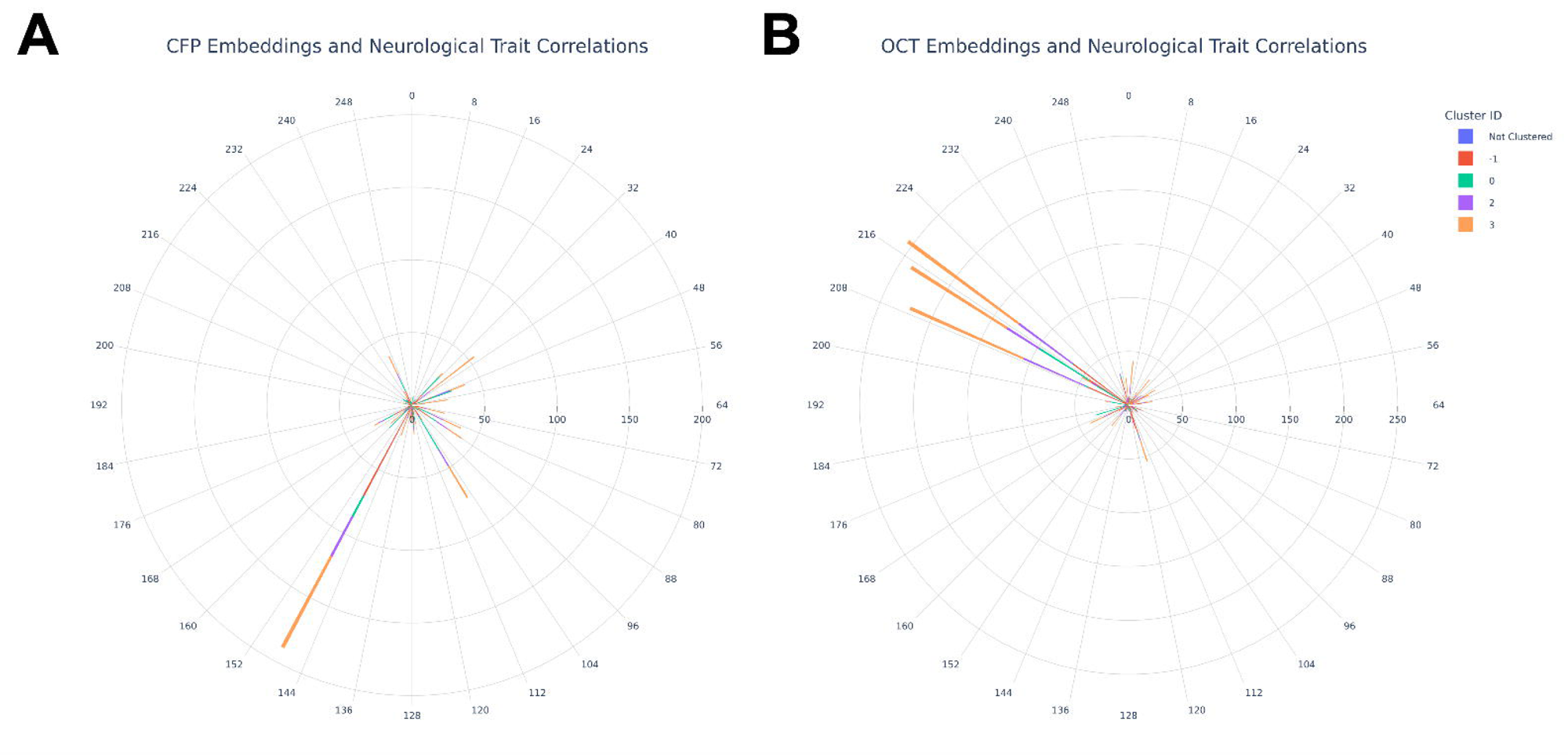
Polar plots displaying the number of significant relationships (corrected for multiple comparisons) between **(A)** CFP-derived embeddings, **(B)** OCT-derived embeddings, and neurological traits. The circumferential axis indicates the embedding number. The radial axis indicates the number of multiple testing corrected significant Pearson correlations that the embedding has with neurological traits. The bars are colour coded according to the cluster of traits the significant relationship belongs to. ‘Cluster 0’ was principally constituted of grey matter volumes assessed using magnetic resonance imaging (MRI); ‘clusters 1-3’ were largely constituted of diffusion MRI values. The individual p values are presented in the supplementary tables and can be referred to in order to appraise specific tests and the strength of significance. HDBSCAN clustering has been used to simplify the presentation of results by grouping traits into statistically defined categories, but readers may wish to explore the more granular trait-wise results in the supplementary content.

As documented previously, Ret-AAE embeddings were also correlated with lipid-related metabolites. Although this data has been presented in relation to cardiovascular disease, lipids are crucial for the normal development and function of the central nervous system, and lipid metabolism has been implicated across the breadth of neurodegenerative disease.^29,30^ Therefore, the associations between Ret-AAE embeddings and lipid metabolism are of equal interest as a potential axis of association between ophthalmic features and neurodegenerative disease.

## Discussion

In this study, we present an integrative multi-omic framework to investigate the links between deep learning-derived ophthalmic imaging features (Ret-AAE embeddings) and systemic disorders. Leveraging comprehensive physiological, anatomical, radiomic, metabolomic and genomic analyses, we provide converging evidence that the observed association between ophthalmic features and systemic disease reflect diverse processes.

We show that OCT and CFP features are associated with a range of cardiovascular and neurodegenerative disorders, both at the time of imaging and as predictors of future disease. Specifically, Ret-AAE embeddings are associated with future heart failure, hypertension, ischaemic heart disease, cerebrovascular disease, Parkinson’s disease, and dementia; mirroring existing evidence of the predictive power of ophthalmic imaging for neurodegenerative and cardiovascular diseases.^1,2,8,9,31,32^ As such, this work builds upon evidence supporting the concept of the eye as a diagnostic window into cardiovascular and neurological health. Furthermore, our study corroborates and extends existing evidence that the ophthalmic features represented in OCT and CFP images offer distinct, complementary insights into systemic health. Previous work by Zhou et al. has indicated that CFP and OCT images do not necessarily predict systemic disease with equal accuracy. In their study (RETFound), it was shown that CFPs offered the most value with respect to stroke prediction, and OCTs in relation to Parkinson’s disease.^9^ In this study, we showed that both CFP and OCT features are associated with cerebrovascular and cardiovascular outcomes, but they differ with regards to the breadth and distribution of these associations. This was apparent throughout our multi-omic analysis, where OCT features had substantially more neurological trait associations, whilst CFPs had more cardiovascular associations; and with regards to future disease risk, where OCTs led by heart failure associations and CFPs by essential hypertension associations.

Saliency maps implicated the choroid and vascular tree in relation to cardiovascular associations, and the optic nerve head, neurosensory retina, and vasculature in relation to neurological associations. Notably, the prominence of choroidal features in our analysis contrasts with the relative paucity of research into choroidal biomarkers for cardiovascular disease^2^. This suggests that further study is warranted, particularly using imaging modalities that visualise the choroid with greater resolution (e.g. swept-source OCT). In some cases, associations were mapped to projection artifacts (e.g. cataract) and CFP background features (e.g. pigmentation or the choroidal vasculature). These associations may reflect variation associated with ancestral background and age. Whilst such variation is of relevance to disease prediction, this finding highlights the importance of considering confounding anatomical factors when studying image-derived phenotypes.^33^

Genetic analyses revealed that Ret-AAE features are enriched for genes in pathways implicated in neurodegenerative disease, including those related to Parkinson’s disease (e.g. alpha-synuclein), Alzheimer’s disease (e.g. amyloid-beta-related pathways), and broader neurodegeneration (e.g. the kynurenine pathway, linked to Huntington’s disease, amyotrophic lateral sclerosis, and dementia).^34,35^ These findings are in keeping with post-mortem studies demonstrating that amyloid and alpha synuclein deposits can be demonstrated in the retina, and may be correlated with the extent of neurodegenerative disease. ^36,37,38^ Additionally, we observed novel statistically significant genetic correlation between cardiovascular traits and vascular tree localised ophthalmic features, suggesting that some ophthalmic features share a genetic architecture with cardiovascular health. Physiologically, our analyses indicated that ophthalmic features are associated with multiple cardiovascular phenotypes, including blood pressure, pulse pressures, arterial stiffness, and cardiac cycle functional volumes. Radiomic analyses revealed that there are broad associations between ophthalmic features and both global and regional cerebral volumes, as well as with cerebral tissue microstructure assessed using diffusion MRI. Metabolomic analysis demonstrated that both OCT- and CFP-derived features are correlated with lipid-related metabolites, suggesting a shared metabolomic axis between ophthalmic features and neurodegenerative/cardiovascular disease. Collectively, these findings are illustrative of the wide-ranging and complex potential intermediaries which may link ophthalmic features to systemic health. While few studies have taken a multi-omic approach to elucidate the biological underpinnings of deep learning-derived retinal phenotypes, our findings reinforce prior evidence linking ophthalmic features to neurological/cardiovascular structure, systemic physiology, metabolomic pathways, and clinical risk factors. ^10,39–42^ Furthermore, this study advances the application of deep learning-derived phenotyping approaches in large-scale biological and clinical research. ^10,39,43,44^

This study has several limitations. Whilst the UK Biobank was designed to be broadly representative of the ageing UK population, it is not without some inevitable selection biases. All reported associations should therefore be interpreted considering this limitation, which may apply to both the cohort as a whole and the missingness of individual measures within the population.^45^ Specifically, it is possible that the relationship between the eye, heart and brain may differ in cohorts with a higher burden of disease, of more heterogeneous ancestry, or of different age distribution (e.g. those below 40 years of age, who are excluded from the UK Biobank study). Secondly, Ret-AAE utilised a 2D representation (the central slice) of OCT images, rather than exploring relationships across the entire 3D macular representation. This simplification, necessitated by computational constraints, risks overlooking relationships with paracentral OCT features and macular asymmetries. We did not seek to explore pharmacological factors within this study, and it is therefore possible that some oculo-systemic associations are pharmacologically mediated, a line of enquiry which may be worthy of further study. This study sought to explore mechanistic links between ophthalmic features and systemic health, but the study methodology was not designed with clinical applications in mind, and the distinction between statistical and clinical significance should be noted. Finally, with the intention of balancing type one and type two error rates, our multiple-testing correction was applied considering each embedding as a family (see section “controlling for type one error risk”), the results may therefore benefit from follow-up in external validation studies, although the risk of type one error is reduced by the convergence of evidence across the diverse analyses conducted in this study.

In summary, this study provides a framework for interpreting the specific systemic pathways that underpin observed imaging-disease associations with respect to their genetic, metabolic, anatomical, and functional characteristics. Here, this was achieved using AAE-derived imaging features, but future research may seek to compare the performance of this approach with alternative methods, such as use of variational autoencoders or other state-of-the-art vision learners^11,46^. By offering enhanced biological interpretability of these associations, our findings can inform future studies aiming to model cardiovascular and neurological disease using ophthalmic imaging, and support the development of explainable deep learning tools for personalised medicine approaches. Finally, through delineation of the spectrum of neurodegenerative and cardiovascular disorders associated with ophthalmic imaging features, we expect this work to guide future models with respect to the diversity of health outcomes that can be modelled.

## Methods

### Ethics statement

The UK Biobank study was (UKB) conducted with the approval of the North-West Research Ethics Committee (ref 06/MRE08/65), in accordance with the principles of the Declaration of Helsinki, all participants gave written informed consent and were free to withdraw at any time.^47^ This project used data from the UKB study under approved project numbers 53144, 49978 & 11350. All retinal images displayed in this manuscript are reproduced with the permission of UKB.

### Cohort characteristics

UKB is a prospective population-based study of 502,355 participants. Baseline examinations were carried out at 22 study assessment centres between January 2006 and October 2010. Study participants have undergone a detailed assessment of demographic, lifestyle and clinical measures; provided DNA samples via blood tests; and provided a range of physical measures. A substantial subset of volunteers underwent an ophthalmic assessment (23%, N = 117,279). Of these participants, 67,664 underwent spectral domain OCT between 2006-2010 (‘instance 0’) and an additional 17,090 participants were imaged for the first time during the first repeat assessment between 2012-13 (“instance 1”) ^48^. UKB data is organised into data categories and fields, and we provide data category/field ID numbers in brackets for reproducibility. The characteristics of those with and without imaging were compared using the Kruskal-Wallis and χ2 tests.

### Imaging quality control

Low quality OCT scans were excluded on the basis of an image quality score below 40.^15^ The quality score utilised was that which is native to the Topcon OCT device. CFPs were excluded if they were of ‘reject’ quality grading assessed using a deep learning method, ‘Automorph’ (‘usable’ and ‘good’ images were retained). ^49^Automorph was trained using the EyePACS-Q dataset, using grading provided by two experts according to image illumination, artifacts, and diagnosability of ocular diseases; the full method is documented in the referenced publication.

### Image pre-processing

For OCTs, FDS files were converted to an analysable format using the oct-converter python package.^50^ OCT slice 64 was retained for analysis. CFPs were cropped and segmented using Automorph and the binary vasculature segmented images were retained for the purpose of refining the autoencoder loss function defined below. Imaging data were randomly split approximately 70:15:15 into train : test : validate allocations for the purpose of training deep learning models. Images were partitioned so that each subject’s scans were paired within the same allocation.

### Autoencoder Model Architecture and Training

#### Image transformations

Several transformations were applied to OCTs and CFPs during the autoencoder training process. To reduce computational burden, images were resized to 224×224 using bicubic interpolation. To improve model generalisability, several further transformations were applied during training, namely: random horizontal flips, random rotations (0-15 degrees), random colour jitter (brightness = 0.9-1.1, contrast = 0.9-1.1, saturation = 0.9 - 1.1).

#### Model architecture

We designed an AAE for the purpose of creating compressed representations of retinal images (CFP and OCT). The AAE architecture is regularized by matching the aggregated posterior, q(*z*), to an arbitrary prior, p(*z*). We selected a Gaussian prior N(0,1), and the resulting vector (*z*) approximates a multivariate Gaussian distribution. ^51^ This property is beneficial for downstream analyses, many of which have a normality assumption. Our AAE was trained in a laterality agnostic manner, but separate AAEs were trained for OCTs and CFPs. The model encoder consisted of each layer of the encoder consisting of a 2D convolution, a batch normalisation layer, a Leaky ReLU, a multi-scale residual block (kernel sizes 3 and 5), and an efficient multi-scale attention module.^52,53^ We projected our image to a 256-dimensional vector using a linear layer at the bottleneck. Our decoder consisted of a single linear layer for up sampling our vector, followed by 2D transposed convolutions, batch normalisation, Leaky ReLU activations, multi-scale residual blocks, and efficient multi-scale attention modules.^52,53^ Our adversarial discriminator was composed of three linear layers and utilized LeakyReLU activation functions.

#### Loss functions

For training both the OCT- and CFP-AAE, we utilised the mean absolute error (MAE, also known as L1 loss) and adversarial loss. CFPs are relatively low contrast images, which are dominated by a narrow background colour distribution (the retina). For our purposes, the retinal vasculature is an important structure which must be represented in the latent space. In our first iterations of training our model, the AAE failed to reconstruct the vasculature to an acceptable extent, due to the vasculature contributing little to the L1 loss (**Supplementary Figure 68**). Using Automorph, we segmented the CFP images and produced binary masks of the vasculature.^49^ On average, the vasculature represented just 4.07% of the pixels in CFPs. Accordingly, for the CFP AAE, we weighted the L1 loss to attribute 12.29x more weight to pixels containing blood vessels on Automorph binary masks. In doing so, CFP vasculature structures then contributed 50% of the L1 Loss on average. In addition, we implemented a perceptual loss function for the CFP AAE. In a previous systematic comparison of deep perceptual loss functions, the 4th ReLU of VGG16 (without batch normalisation) was shown to contribute to the best performance in autoencoding tasks, and, accordingly, this model was selected for our perceptual loss function.^54^ In order to calculate a perceptual loss, original images and AAE reconstructions were normalised according to ImageNet parameters before being passed to VGG16 without batch normalization.^55^ The mean squared error loss of the feature maps of the reconstructed and original images then served as the perceptual loss. Finally, with a view to normalising the latent vectors produced by the model, we utilised an adversarial loss function. Briefly, this was calculated by training a discriminator network to distinguish between the latent vectors of the AAE model and a standard normal distribution. To improve the stability of the adversarial network, we utilised one sided label smoothing.^56^ The weight provided to each constituent of the loss function was determined using a Bayesian hyperparameter sweep as described below. In our CFP AAE, terminal arterioles and venules were lost in reconstruction. Although we identified that the SSIM improved with expansion of the latent space (e.g., 128-dimensional vectors resulted in SSIM of 0.876, and 256-dimensional vectors in 0.891), in the interest of preserving the statistical and computational feasibility of downstream tasks, we did not expand the latent space dimensionality further in the pursuit of terminal vascular reconstruction.

#### Image quality metrics

Image quality metrics utilised included the PSNR, SSIM, and the L1 loss.^57^

#### Hyperparameter search

We performed a Bayesian hyperparameter sweep using the Weights and Biases platform (version 0.19.8).^58^ The SSIM in the test dataset guided the Bayesian optimization. For both CFP and OCT AAEs, the search was used to select the discriminator learning rate, the autoencoder learning rate, the number of convolutional layers, the optimizer weight decay, and the weighting of the adversarial loss. For the CFP sweep, we additionally included the perceptual loss weighting. The sweep was conducted for 35 epochs per hyperparameter combination. For the CFP AAE, the sweep-identified optimal hyperparameters included 5 convolutional layers, weight decay of 0.01 (AdamW optimiser), autoencoder learning rate of 0.0005, discriminator learning rate of 0.0001, a weighting of 0.2 for the perceptual loss, and a weighting of 0.001 to the discriminator loss.^59^ For the OCT AAE, the optimal hyperparameters included 4 convolutional layers, weight decay of 0.001 (AdamW optimizer), autoencoder learning rate of 0.0001, discriminator learning rate of 0.00005, and weighting of 0.0001 for the discriminator loss.

#### Additional AAE experimental details

Experiments were conducted using PyTorch.^60^ We utilised a learning rate to warm up over the first 15 epochs. The batch size was 32. A random seed of value 42 was used. The CFP model was trained for 400 epochs, and the OCT model was trained for 250. Our model was trained using 4 Tesla T4 GPUs. Randomly selected reconstructions were examined by two Ophthalmologists (T.H.J and P.I.S) to ensure critical structures were adequately represented in reconstructed images.

### OCT and CFP AAE phenotypic association tests

We utilised latent vectors representing the left eye (OCT and CFP) of UKB subjects as a means of reducing the dimensionality of the analysis. In keeping with previous work, the left eye was selected because the relevant images are systematically of higher quality than those from the right eye.^11,15^ The total number of subjects varied according to the requirements of the association test, and are reported in text where possible, or in the supplementary content where subject number varied according to individual phenotype. Both instance 0 and 1 ophthalmic images were used in these analyses (except where explicitly stated otherwise), and where a subject had undergone imaging during both instances only their first set of images were considered.

#### Pearson correlation analyses

The ‘positive control’ Pearson correlation analyses included all traits in the UKB eye measures category (Category 100013), excluding right eye measures.

The experimental tests featured a combination of anatomical and functional cardiovascular and neurological traits. Specifically, this featured all traits in the following UKB catalogues: brain MRI (Category 1014), cognitive function (Category 1005), heart MRI (Category 1015) carotid ultrasound (Category 101) and selected traits from the physical measures category (Category 1006). There were a total of 50 cardiovascular traits, and 892 neurological traits. The large number of neurological traits reflects the scale of derived neuroimaging features available in UKB. To enhance interpretability in plots, we utilised HDBSCAN clustering.^61^ Traits present in an overlapping, large number of subjects were clustered, and the remainder of traits were assigned to a “not clustered” category - but were still considered in subsequent analyses. Data were scaled according to the interquartile range using ‘*RobustScaler’* in SciKit Learn.^62^ Clusterable features included 35 ophthalmic traits (N=62,729), 889 neurological features (N=12,219), and 37 cardiovascular traits (N=11,295). A leaf clustering method was utilised. Cardiovascular and ophthalmic traits were fewer in number, so a minimum cluster size of 3 was used. Neurological traits were assigned a minimum cluster size of 10 given the larger number of traits.

For each analysis, outlier cardiovascular, neurological or ophthalmic values were excluded. This was achieved by excluding data greater than 2.5 standard deviations from the mean. Pearson correlation coefficients and accompanying *p*-values were then calculated using the SciPy python package.^63^

#### Cox proportional hazards and Welch’s t-test analyses

Disease outcomes were derived from the “First Occurrences” UKB catalogue (Category 1712). The UKB first occurrences were generated by mapping read code information in the ‘Primary Care data’ (Category 3000), ICD-9 and ICD-10 codes in the ‘Hospital inpatient data’ (Category 2000), ICD-10 codes in ‘Death Register records’ (Field 40001, Field 40002), and ‘self-reported medical condition codes’ (Field 20002) reported at the baseline or subsequent UKB assessment centre visit. The UKB estimates that hospital inpatient data and death are mostly complete up to 31.05.2022, after which data may be incomplete.^64^ Accordingly, we utilised 31.05.2022 as the censoring date for our time-to-event analysis. For the Cox proportional hazards, subjects who suffered a given disease event before the date of retinal imaging were excluded from analysis. Meanwhile, for our t-test, the objective was to determine if latent features differed significantly according to the presence/absence of disease at the time of scan, and so for this analysis those with disease events at the time of scan were included. Disease outcomes included (1) a positive control disease dataset inclusive of diseases that were determined to be identifiable on retinal imaging according to two Ophthalmologists (T.H.J and P.I.S), including disorders of the retina, choroid, optic nerve, and lens (19 diseases); (2) all available neurodegenerative diseases (10 diseases), (3) all available cardiovascular disorders (23 diseases). The complete list of diseases considered is presented in **Supplementary Table 1.31**.

Cox proportional hazards modelling was conducted using the lifelines Python package.^65^ The primary outcome of interest was the relationship between left eye Ret-AAE embeddings and time-to-event analyses against ophthalmic, cardiovascular and neurodegenerative diseases. Covariates included genetic sex and age at time of scan. t-tests were conducted using the SciPy package in Python.^63^ We elected to utilise the Welch’s t-test, a modification of the Student’s t-test which is more reliable in the context of unequal sample sizes of variances between two populations.^66^ Results with less than 30 samples per group were excluded from the final analysis in both analyses. We were unable to test the baseline relationship between Ret-AAE embeddings and Huntington’s disease, spinal muscular atrophy and related syndromes, Alzheimer’s disease, and vascular dementia due to insufficient cases (<30) at baseline imaging. Huntington’s disease was additionally excluded from Cox proportional hazards analysis due to insufficient cases.

#### Canonical correlation analysis

CCA was used to explore the relationship between CFP/OCT-derived embeddings and 251 metabolic biomarkers generated by Nightingale Health (Category 220).^67^ Due to subject overlap in instance 0 and 1 metabolic sampling, only instance 0 data were used in this analysis. Subjects with missing imaging or missing metabolite samples were excluded. Extreme values which may unduly influence the results were managed using Winsorization (applied to the most extreme upper and lower 1% of values)^68^. The analysis was conducted using the SciKit learn CCA model. Embeddings and metabolomic data were scaled using the native CCA scaler. To assess the statistical significance of each canonical variate, we used permutation testing to create a null distribution of correlations. The embedding matrices were held constant, whilst the metabolomic matrix was substituted for randomly permuted sequences over 1000 iterations.^69,70^ The *p*-value was calculated using the number of null correlations that exceeded the average CCA estimated on the original dataset. The number of modes to test in our permutation test was determined by constructing a Scree plot of the first 10 modes and then truncating where there is a substantial reduction to the regression coefficient.

#### Sparse partial least squares

In addition to CCA, we explored metabolomic data using a canonical sparse partial least squares (SPLS) analysis using the mixOmics (version 6.32.0) R library^71^. SPLS is a variant of partial least squares in which a subset of important features is selected for inclusion in the model according to sparsity hyperparameters, rather than including all traits, which may be inappropriate in the context of highly correlated covariates (as is the case for metabolomic data). SPLS was therefore implemented as a means of validating our CCA results in a manner more robust to the structure of our data. Data were prepared in line with the CCA analysis. Lasso penalisation was optimised using five-fold cross validation with five repeats, optimised to maximise correlation between predicted and actual components. Following this, we implemented CFP SPLS with two components; 30 embedding variables in component one, and 10 in component two; and 25 metabolite variables in component one, and 10 in component two. OCT SPLS was implemented with one component, with 10 embedding variables and 10 metabolite variables. The analysis was undertaken using the canonical mode and plots presented with respect to SPLS were all generated using mixOmics.

### Genetic Analyses

#### Choice of GWAS method

REGENIE version 4.1 was selected as the method for GWAS.^72^ For step 1, we utilised the UKB microarray data (Field 22418). For step 2, we utilised the imputed data (Field 22828).

#### Phenotypes and covariate data

The phenotypes studied were latent features for each subject’s left eye OCT and CFP image. Therefore, 256 GWAS studies were performed per subject per scan type (i.e. 512 GWAS studies in total). Covariates for GWAS were sex (Field 31), age at date of scan (Field 22006) and spherical equivalent refractive error. Spherical equivalent refractive error was calculated by identifying the most reliable refractometry result (Field 5276), identifying the spherical refractive error (Field 5085) and cylindrical refractive error (Field 5086), and performing the calculation: spherical equivalent = spherical refractive error + 0.5 x cylindrical refractive error.^15^

#### Quality control for genetic analyses

The genetic analyses in the discovery cohort were conducted in subjects in instance 0 (initial assessment visit, 2006-2010). The genetic analyses in the replication cohort were conducted in subjects who attended within instance 1 (first repeat assessment visit, 2012-13). Any overlapping subjects were removed (i.e. the cohorts are entirely independent). Analysis was limited to subjects of genetically determined European ancestry according to their principal components (Field 22006).

Subject-level genetic quality control metrics were applied, including: exclusion of subjects whose genetic sex (Field 22001) does not match their reported sex (Field 31); exclusion of subjects with sex chromosome aneuploidy (Field 22019); exclusion of subjects with genetic kinship to other UKB participants (Field 2202); exclusion of subjects who were outliers for heterozygosity or missing rate (Field 22027), and removal of individuals with more than 10% missing genotype data.^73^

Variant level quality control was conducted using plink2.^74^ For the microarray data used in REGENIE step 1, quality control included: minimum minor allele frequency of 0.01, minimum minor allele count of 20, removal of variants with >10% missingness, and removal of SNPs failing Hardy-Weinberg Equilibrium (HWE) test at p < 1e-15. For imputed data utilised in Step 2, quality control included: minimum minor allele frequency of 0.01, minimum minor allele count of 20, removal of variants with >10% missingness, removal of SNPs failing Hardy-Weinberg Equilibrium test at p < 1e-6, and removal of duplicate variants. Additionally, following GWAS, any variants with an INFO score below 0.8 were removed.

#### GCTA COJO

To refine the obtained association signals, further analyses were performed using GCTA-COJO.^12^ Genetic variants in loci that were on different chromosomes or more than 1500 Kb distant from each other were assumed to be uncorrelated. Otherwise, GCTA COJO default parameters were implemented.

#### Gene set analysis

Gene set analysis was performed using MAGMA.^24^ Gene sets were derived from the Human MSigDB Collections.^75^ Specifically, we elected to utilise the canonical pathways from the curated gene sets. The canonical pathways include the BioCarta pathways database (292 gene sets), the KEGG MEDICUS pathway database (658 gene sets), the PID pathway database (196 gene sets), the Reactome pathway database (1787 gene sets), WikiPathways pathway database (885 gene sets), and the KEGG (legacy) pathway database (186 gene sets). Variants were annotated to genes based on dbSNP version 135 variant locations and NCBI 37.3 gene definitions. The linkage disequilibrium reference file was derived from the ‘European’ subset of the 1000 Genomes Project dataset.^76^

#### Global genetic correlation analysis

The global genetic correlation between OCT/CFP latent features and cardiovascular/ neurological disease was assessed using LD score regression.^77^ The GWAS summary statistics for cardiovascular and neurological disease were sourced from publicly available studies in the same genome build used in this study (GRCh37) and containing the required information for genetic correlation.^78–83^ Only latent features with a mean chi^2^ > 1.02 were considered for LD score regression (the threshold for polygenicity). Our analysis was consistent with the methodology outlined by Bulik-Sullivan *et al*., in which genetic variants were filtered according to: presence in HapMap3; minor allele frequency > 0.01; INFO score >0.9; removal of strand-ambiguous variants and duplicated variants. LD scores were derived from the European subset of the 1000 Genomes Project dataset.^76^ The intercept was not constrained in this analysis. LD score regression was also utilised to determine the heritability of the latent features, where a heritability estimate has been provided for traits with evidence of polygenicity, but not an estimate of statistical significance as this is not output by the LDSC method.

#### Gradient-weighted class activation mapping

To enhance the interpretability of our learned embeddings, we applied Gradient□weighted Class Activation Mapping (Grad□CAM).^84^ Consistent with previous work, we observed that visualizations from the earliest convolutional layers lacked semantic relevance, whereas those from the deepest layers were overly coarse to localize meaningful image features.^85^ Consequently, we focused our primary analyses on the third convolutional layer for both CFP and OCT modalities, which provided the optimal balance between spatial resolution and semantic content. In instances where Grad□CAM failed to yield an interpretable map, we instead employed Layer□CAM or, alternatively, targeted adjacent convolutional layers; these exceptions are clearly noted in each figure legend.^86^ Saliency maps are only presented for embeddings which are referenced in text. Where possible, we describe embeddings with respect to their anatomical localisation in order to enhance interpretability (**Supplementary Figures 19-48**).

#### Controlling for type one error risk

We elected to implement family-wise error rate control (Bonferroni corrected threshold for significance) in this analysis. We examined whether autoencoder embeddings were substantially correlated to guide our definition of a family. As can be seen in **Supplementary Figure 69-70**, the embeddings demonstrated little meaningful correlation. Accordingly, each embedding was treated as its own family, and multiple testing correction was applied according to the total number of tests within each embedding. Given that this is not a **study-wide** correction (i.e., we do not also correct according to the number of embeddings, which would be prohibitively strict / cause excessive type two error given the study scale and hypothesis-free nature of this work), we assess our results and the risk of type one error in the context of the convergent evidence presented across analyses, present raw p values to allow interpretation by readers, interpret results in the context of their biological plausibility, and where possible validate results in a separate UK Biobank instance. Although this approach offers a more nuanced perspective on the results, type one error remains an area of concern, and the key study findings will benefit from external experimental validation when appropriate external multi-omic datasets become available. The Bonferroni corrected *p*-value thresholds for significance for all other analyses are presented in the appropriate supplementary table legends.

## Data Availability

All data are available in the supplementary materials.

https://github.com/TomJulian/Ret-AAE

## Model availability

Due to UK Biobank policies regarding generative models, we are unable to share the Ret-AAE decoder weights. However, we are permitted to share the model encoder and its associated weights, and we have made these publicly available on GitHub: https://github.com/TomJulian/Ret-AAE

## Acknowledgements

We acknowledge the following sources of fundings: the Medical Research Council (MRC) Clinical Research Training Fellowship (MR/Z504105/1, to T.H.J); the Intramural Research Program at the National Eye Institute/National Institute for Health (to E.Y.C); the Wellcome Trust (224643/Z/21/Z, Clinical Research Career Development Fellowship, to P.I.S); the MND Association (894-791), TargetALS, Tambourine’s ALS Breakthrough Research Fund in partnership with the Milken Institute, and the ALS Association (23-PP-664), all to J.C.K; the British Heart Foundation Manchester Research Excellence Award (RE/24/130017, to J.M.D); the EMBL European Bioinformatics Institute (EMBL-EBI) (E.Y, T.F and E.B); the UK Research & Innovation Future Leaders Fellowship (MR/T019050/1), Moorfields Eye Charity with The Rubin Foundation Charitable Trust (GR001753), and an Alcon Research Institute Senior Investigator Award, to P.A.K; the Royal Academy of Engineering under the RAEng Chair in Emerging Technologies (INSILEX CiET1919\/19), ERC Advanced Grant – UKRI Frontier Research Guarantee (INSILICO EP\/Y030494/1), the UK Centre of Excellence on in-silico Regulatory Science and Innovation (UK CEiRSI) (10139527), the National Institute for Health and Care Research (NIHR) Manchester Biomedical Research Centre (BRC) (NIHR203308), and the BHF Manchester Centre of Research Excellence (RE\/24/130017), all to A.F.F, J.H, H.D, J.M.D.

We acknowledge the contribution of the UK Biobank Eye and Vision Consortium. Members of this consortium include: Naomi Allen, Tariq Aslam, Denize Atan, Sarah Barman, Jenny Barrett, Paul Bishop, Graeme Black, Tasanee Braithwaite, Roxana Carare, Usha Chakravarthy, Michelle Chan, Sharon Chua, Alexander Day, Parul Desai, Bal Dhillon, Andrew Dick, Alexander Doney, Cathy Egan, Sarah Ennis, Paul Foster, Marcus Fruttiger, John Gallacher, David Garway-Heath, Jane Gibson, Jeremy Guggenheim, Chris Hammond, Alison Hardcastle, Simon Harding, Ruth Hogg, Pirro Hysi, Pearse Keane, Peng Tee Khaw, Anthony Khawaja, Gerassimos Lascaratos, Thomas Littlejohns, Andrew Lotery, Robert Luben, Phil Luthert, Tom Macgillivray, Sarah Mackie, Savita Madhusudhan, Bernadette Mcguinness, Gareth Mckay, Martin Mckibbin, Tony Moore, James Morgan, Eoin O’Sullivan, Richard Oram, Chris Owen, Praveen Patel, Euan Paterson, Tunde Peto, Axel Petzold, Nikolas Pontikos, Jugnoo Rahi, Alicja Rudnicka, Naveed Sattar, Jay Self, Panagiotis Sergouniotis, Sobha Sivaprasad, David Steel, Irene Stratton, Nicholas Strouthidis, Cathie Sudlow, Zihan Sun, Robyn Tapp, Dhanes Thomas, Emanuele Trucco, Adnan Tufail, Ananth Viswanathan, Veronique Vitart, Mike Weedon, Alastair K. Denniston, Peng T. Khaw, Konstantinos Balaskas, Cathy Williams, Katie Williams, Jayne Woodside, Max Yates, Jennifer Yip, Yalin Zheng.

