## Supplementary Figures for "Ophthalmic imaging as a measure of cardiovascular and neurological health: a multi-omic analysis of deep-learning derived phenotypes"

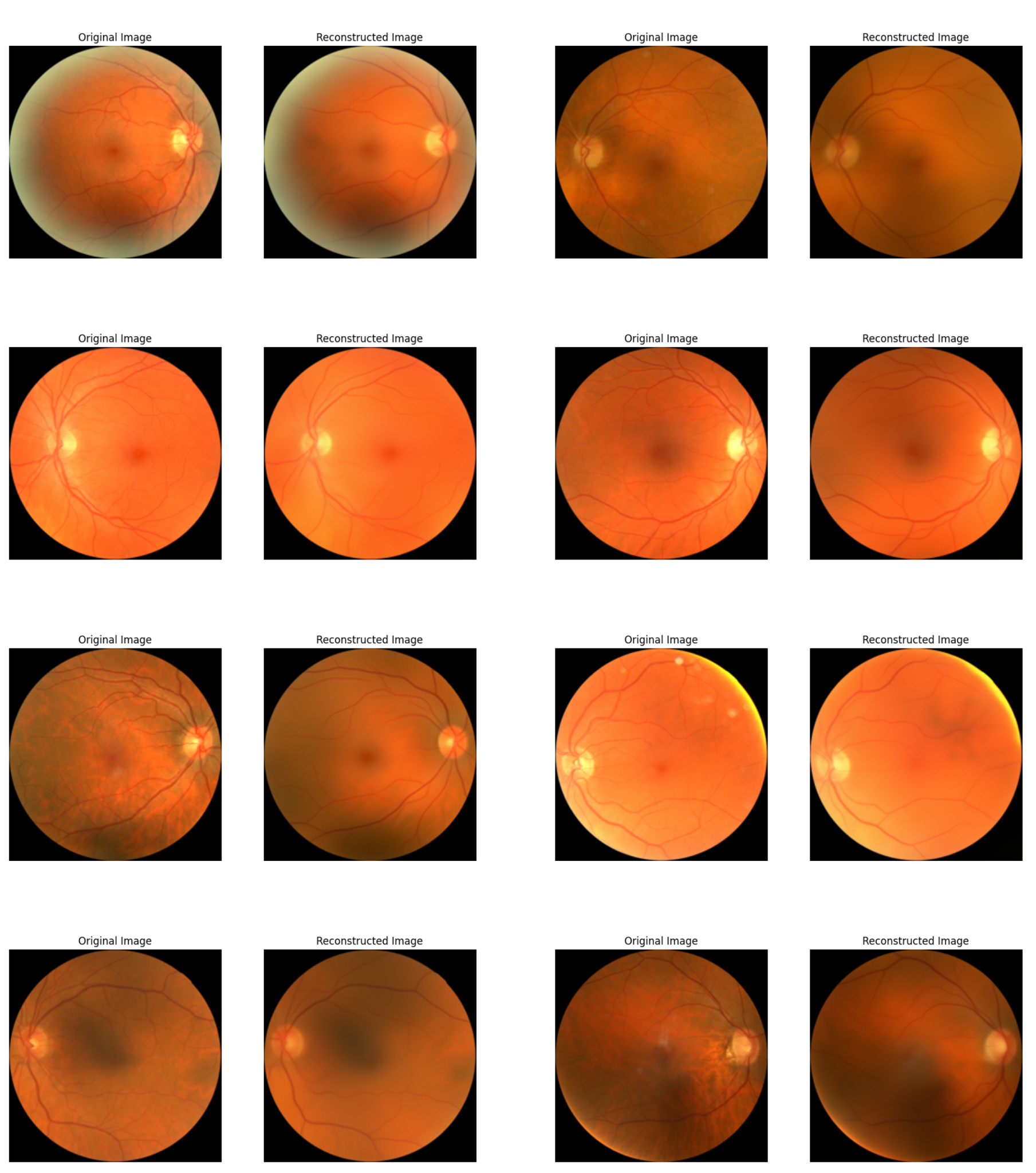

**Supplementary Figure 1:** Eight randomly selected examples of CFP reconstructions (images unseen during training). Reproduced with the permission of UK Biobank.

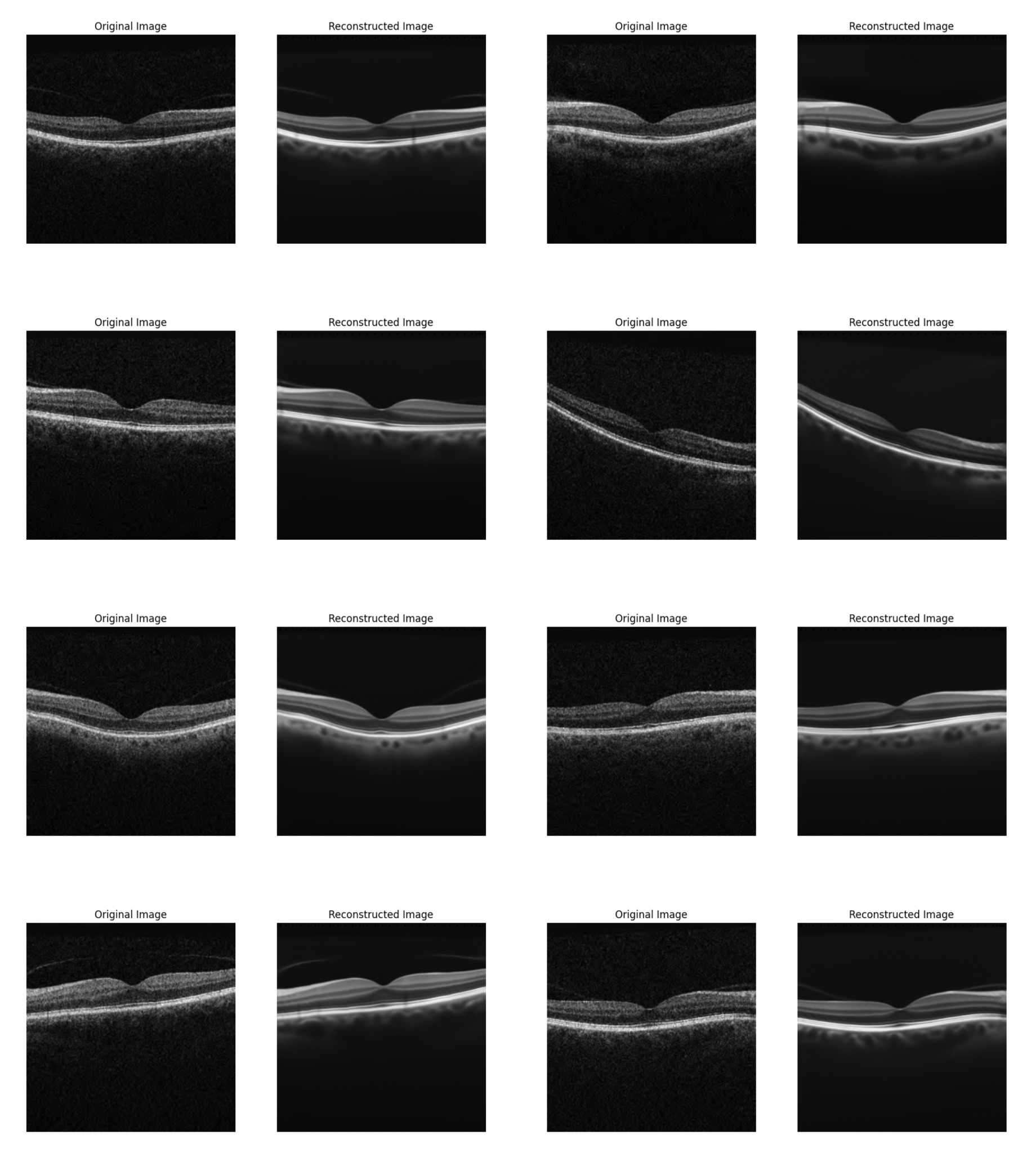

**Supplementary Figure 2:** Eight randomly selected examples of OCT reconstructions (images unseen during training). Reproduced with the permission of UK Biobank.

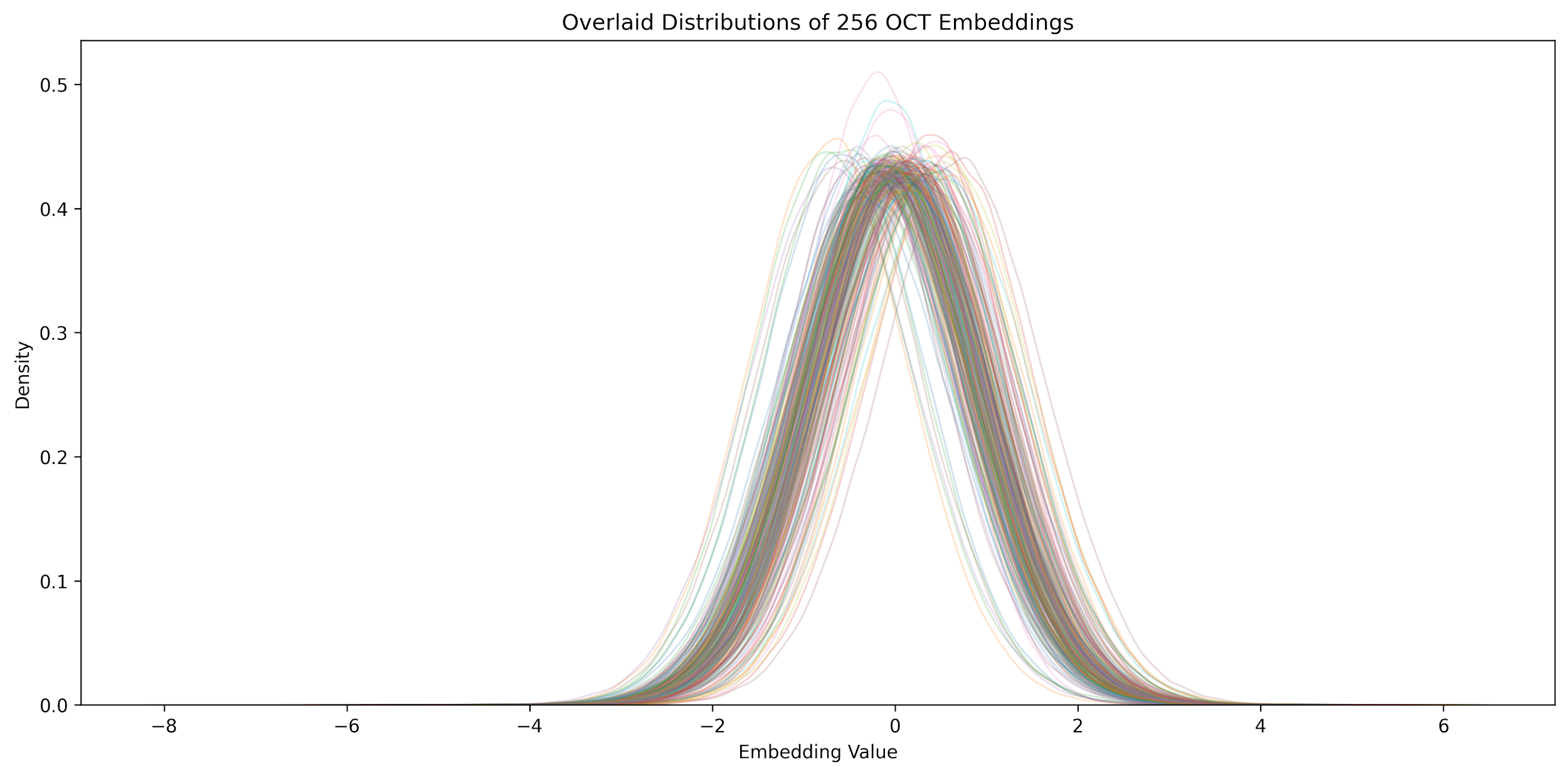
**Supplementary Figure 3**: A density plot, illustrating that most OCT embeddings approximated a Gaussian distribution.

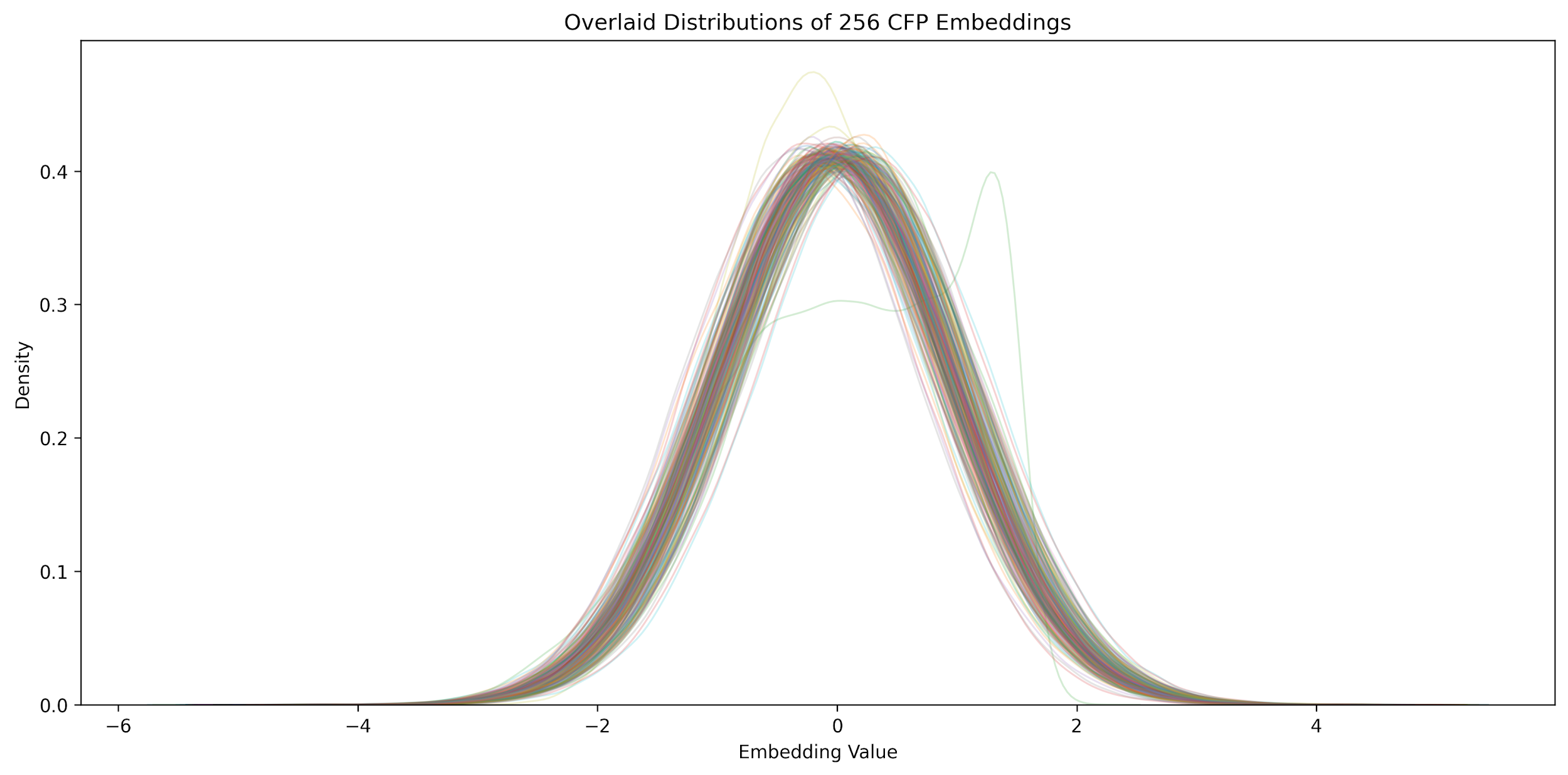
 **Supplementary Figure 4**: A density plot, illustrating that most CFP embeddings approximated a Gaussian distribution.

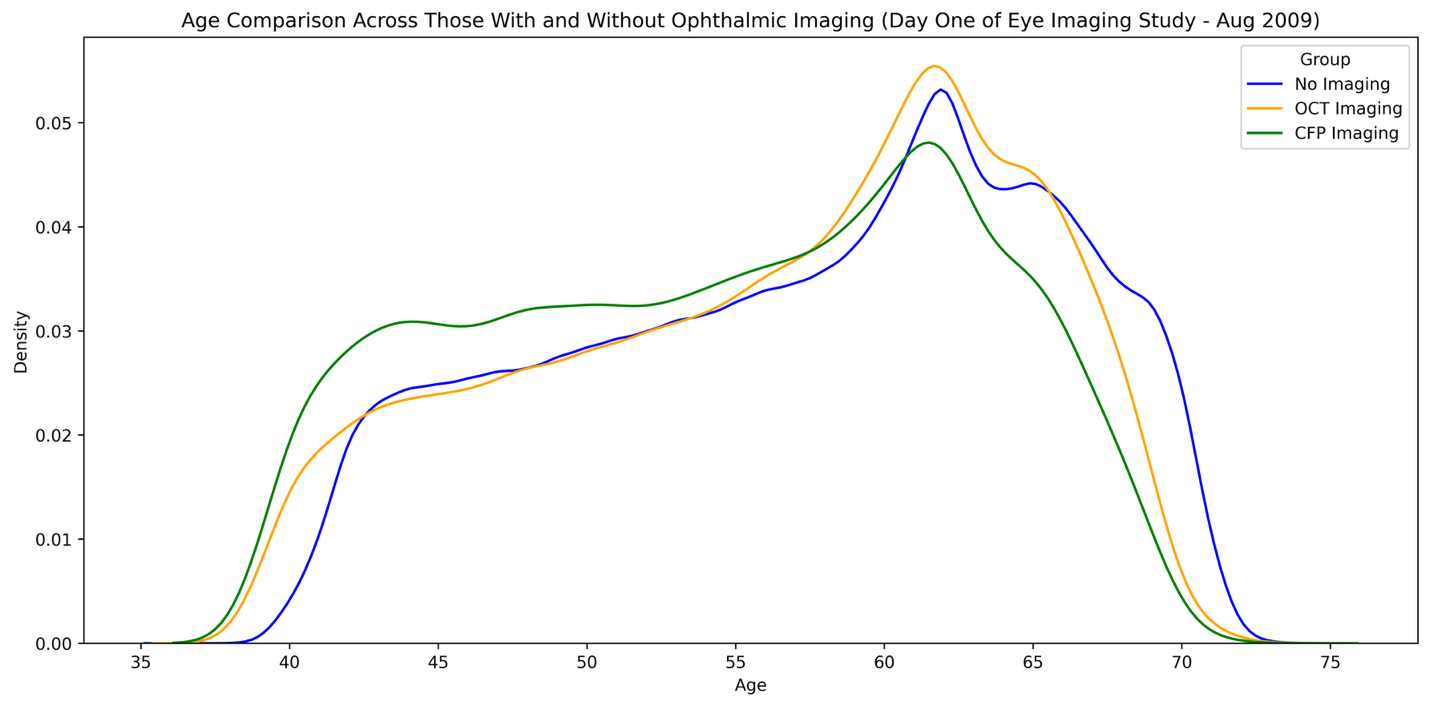

**Supplementary Figure 5**: A density plot showing the distribution of age of subjects at the commencement of the UK Biobank Eye Imaging Study. The plot compares those without imaging, those with sufficiently high-quality left eye colour fundus photograph (CFP) images, and those with sufficiently high-quality left eye optical coherence tomography (OCT) images. Quality criteria are described in the methods section of the manuscript.
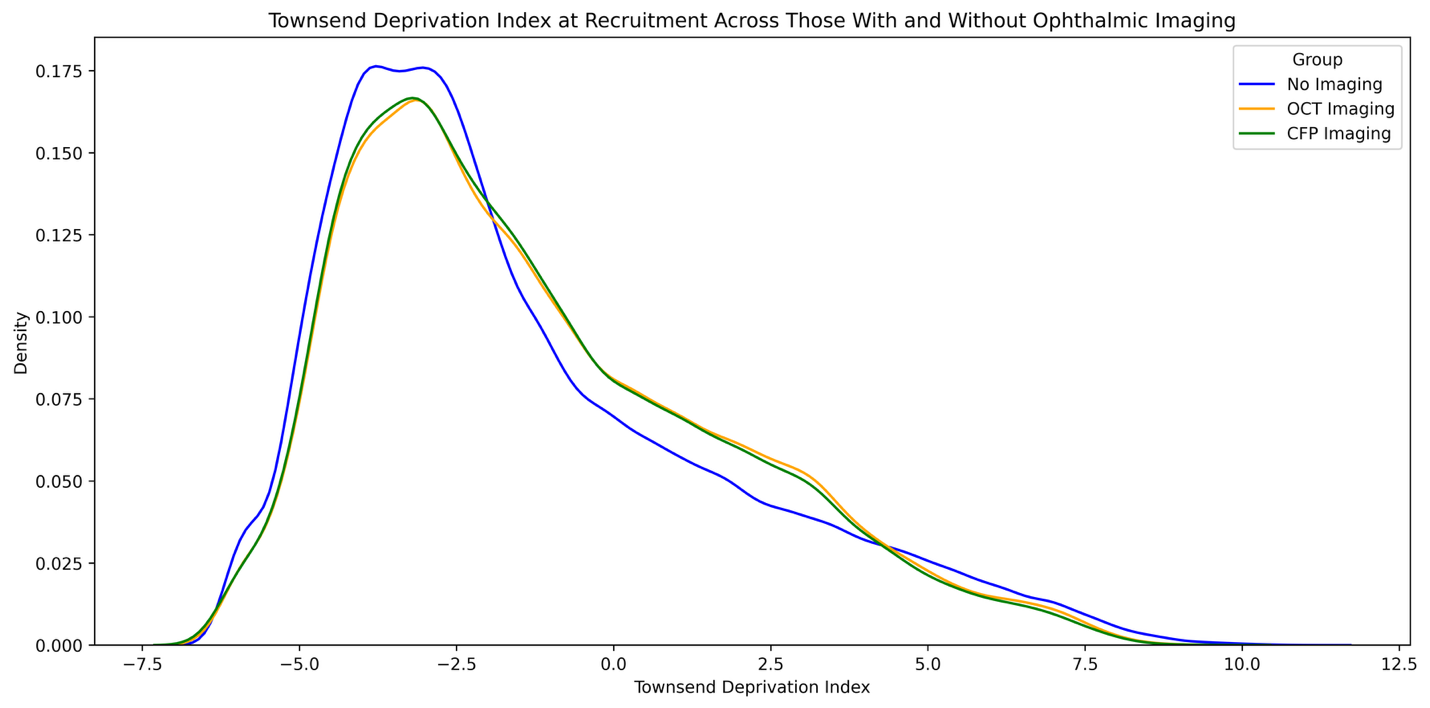
 **Supplementary Figure 6**: A density plot showing the distribution of Townsend deprivation index of UK Biobank subjects. The plot compares those without imaging, those with sufficiently high-quality left eye colour fundus photograph (CFP) images, and those with sufficiently high-quality left eye optical coherence tomography (OCT) images. Quality criteria are described in the methods section of the manuscript.
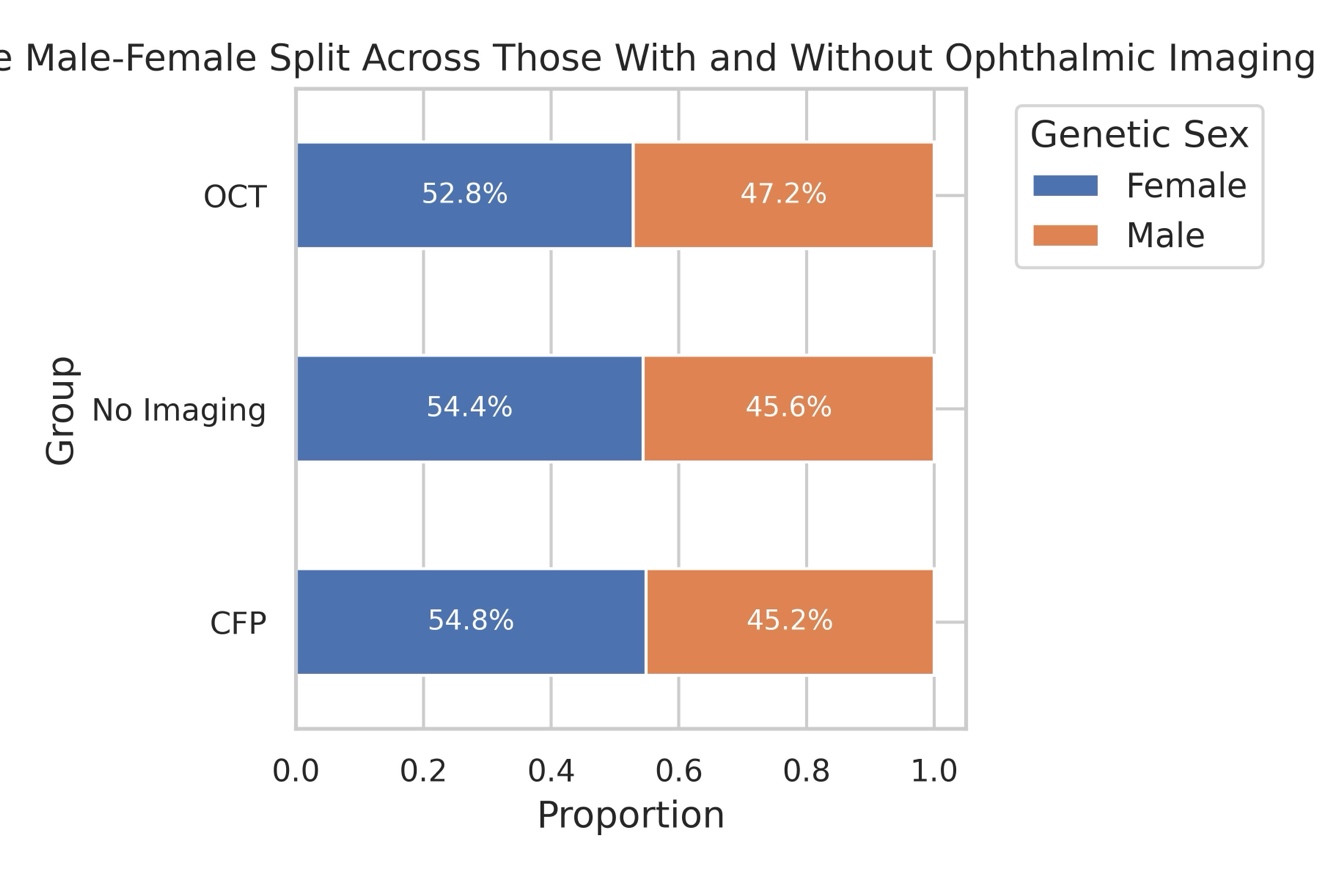

**Supplementary Figure 7:** A bar plot showing the male : female (genetic sex) split of UK Biobank subjects. The plot compares those without imaging, those with sufficiently high-quality left eye colour fundus photograph (CFP) images, and those with sufficiently high-quality left eye optical coherence tomography (OCT) images. Quality criteria are described in the methods section of the manuscript.

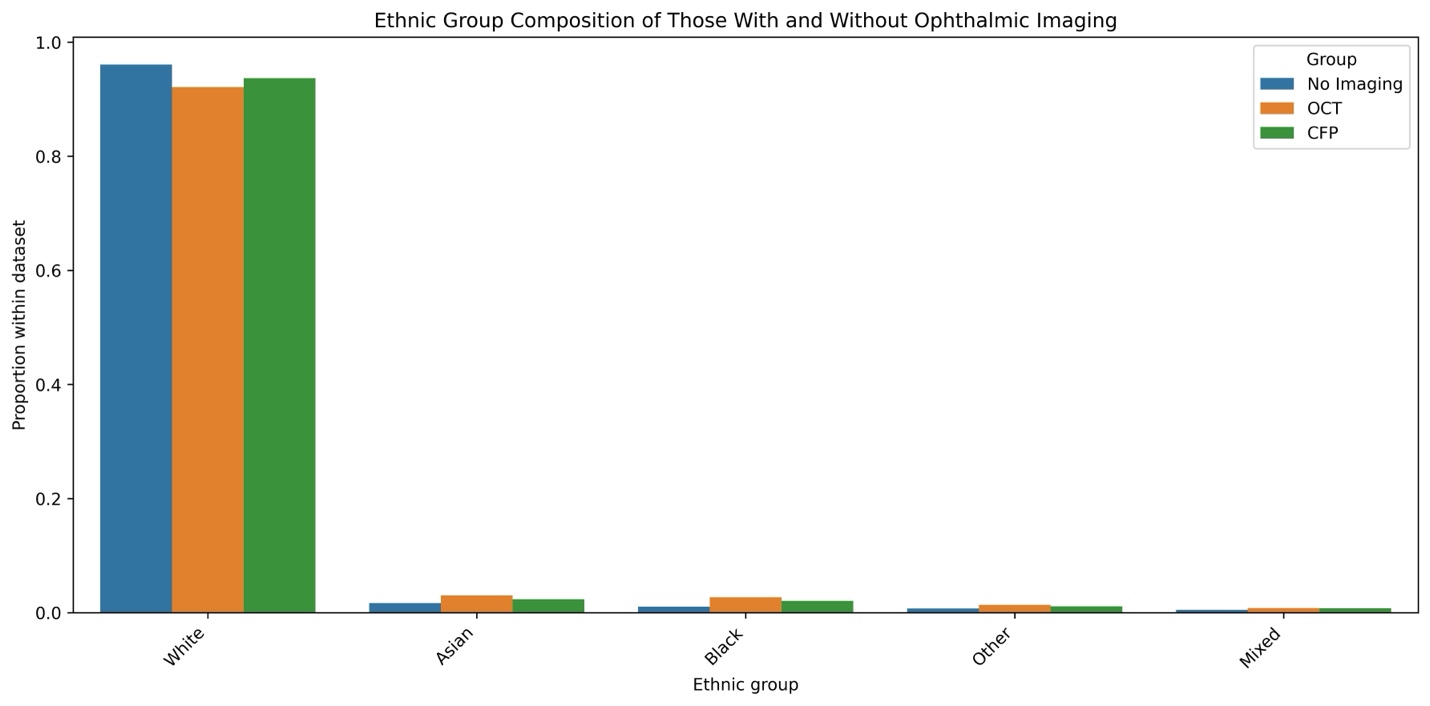

**Supplementary Figure 8:** A bar plot showing the self-reported ethnicity split of UK Biobank subjects. The plot compares those without imaging, those with sufficiently high-quality left eye colour fundus photograph (CFP) images, and those with sufficiently high-quality left eye optical coherence tomography (OCT) images. Quality criteria are described in the methods section of the manuscript. Ethnic groups were defined as per the UK 2021 census (with some truncation of full group names to allow them to fit on the plot axis). The ethnic groups as defined by the UK government are available here: <https://www.ethnicity-facts-figures.service.gov.uk/style-guide/ethnic-groups/>

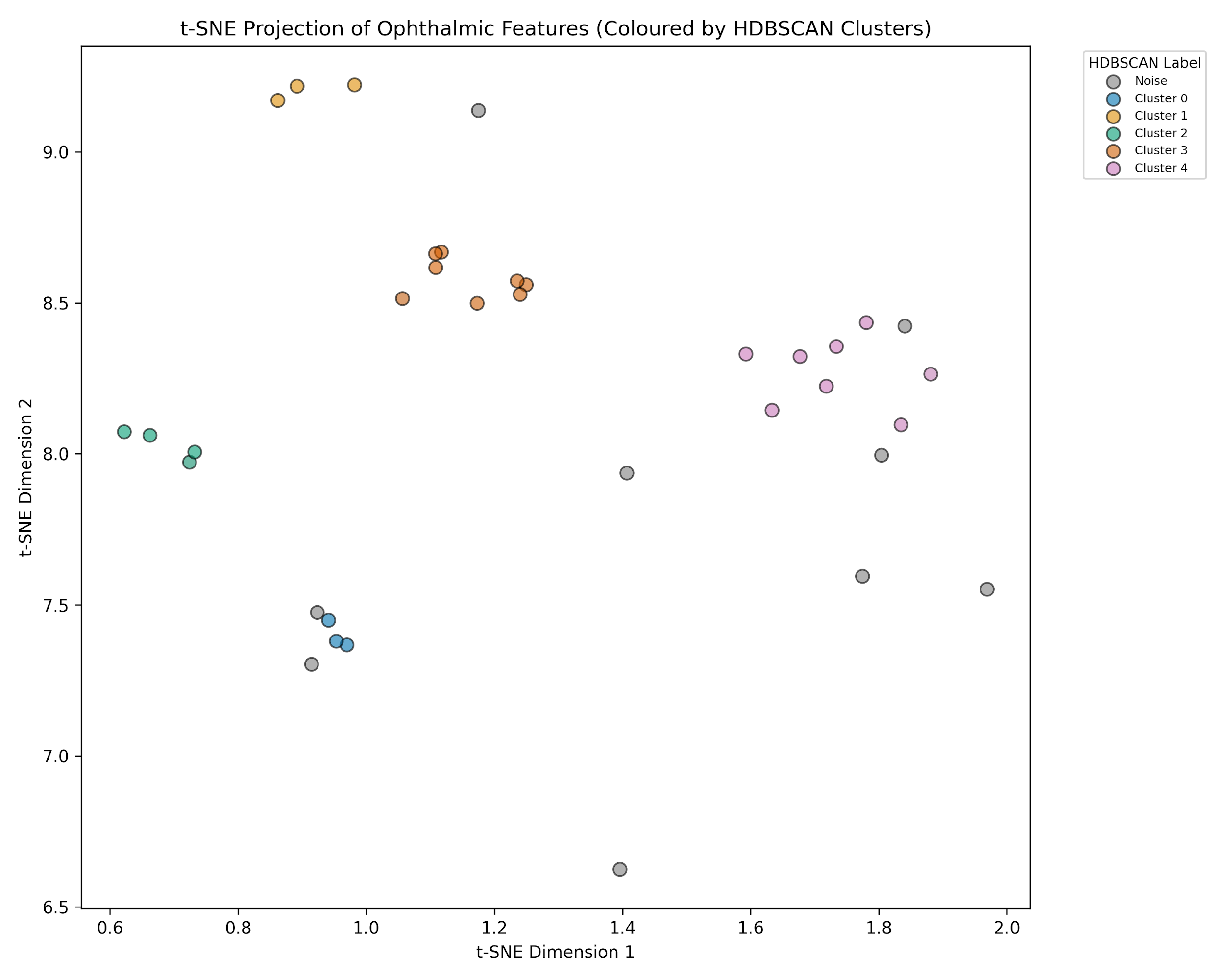

**Supplementary Figure 9:** A t-SNE projection of ophthalmic features clustered using HDBSCAN. The clusters have been color coded according to the HDBSCAN cluster.

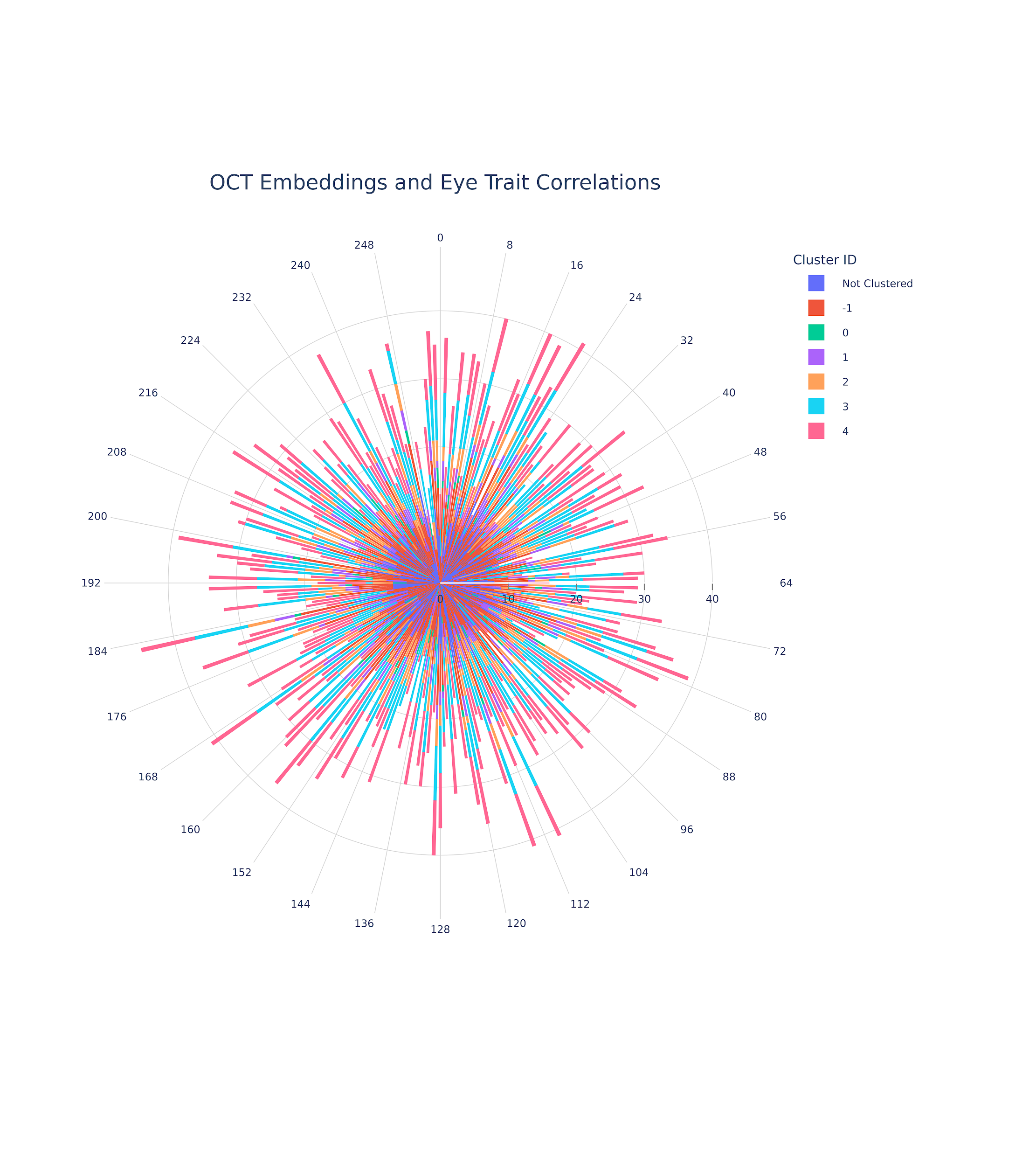
**Supplementary Figure 10:** A polar plot illustrating the relationships between ophthalmic traits and OCT embeddings. HDBSCAN identified four data clusters: ‘cluster 0’ included intraocular pressure and corneal hysteresis measures; ‘cluster 1’ included OCT derived photoreceptor layer thickness measures; ‘cluster 2’ included OCT derived thickness of the photoreceptor & retinal pigment epithelium layers; ‘cluster 3’ included a number of OCT derived thickness measures including the inner nuclear layer, outer plexiform layer, photoreceptors and retinal pigment epithelium; and ‘cluster 4’ contained OCT derived macular thickness measures. Inner retinal thickness measures (e.g. retinal nerve fibre layer and ganglion cell layer thickness). 15 traits could not be clustered due to a high level of missingness across the population. and visual acuity did not fall into HDBSCAN clusters, and were categorized as noise (cluster -1). The circumferential axis indicates the embedding number. The radial axis indicates the number of multiple testing corrected significant Pearson correlations that the embedding has with Ophthalmic traits. The bars are color coded according to the cluster of traits the significant relationship belongs to. There were more significant relationships between OCT embeddings and Ophthalmic traits than CFP embeddings and Ophthalmic traits. This is expected given that most UKB ocular traits were derived from OCT scans.
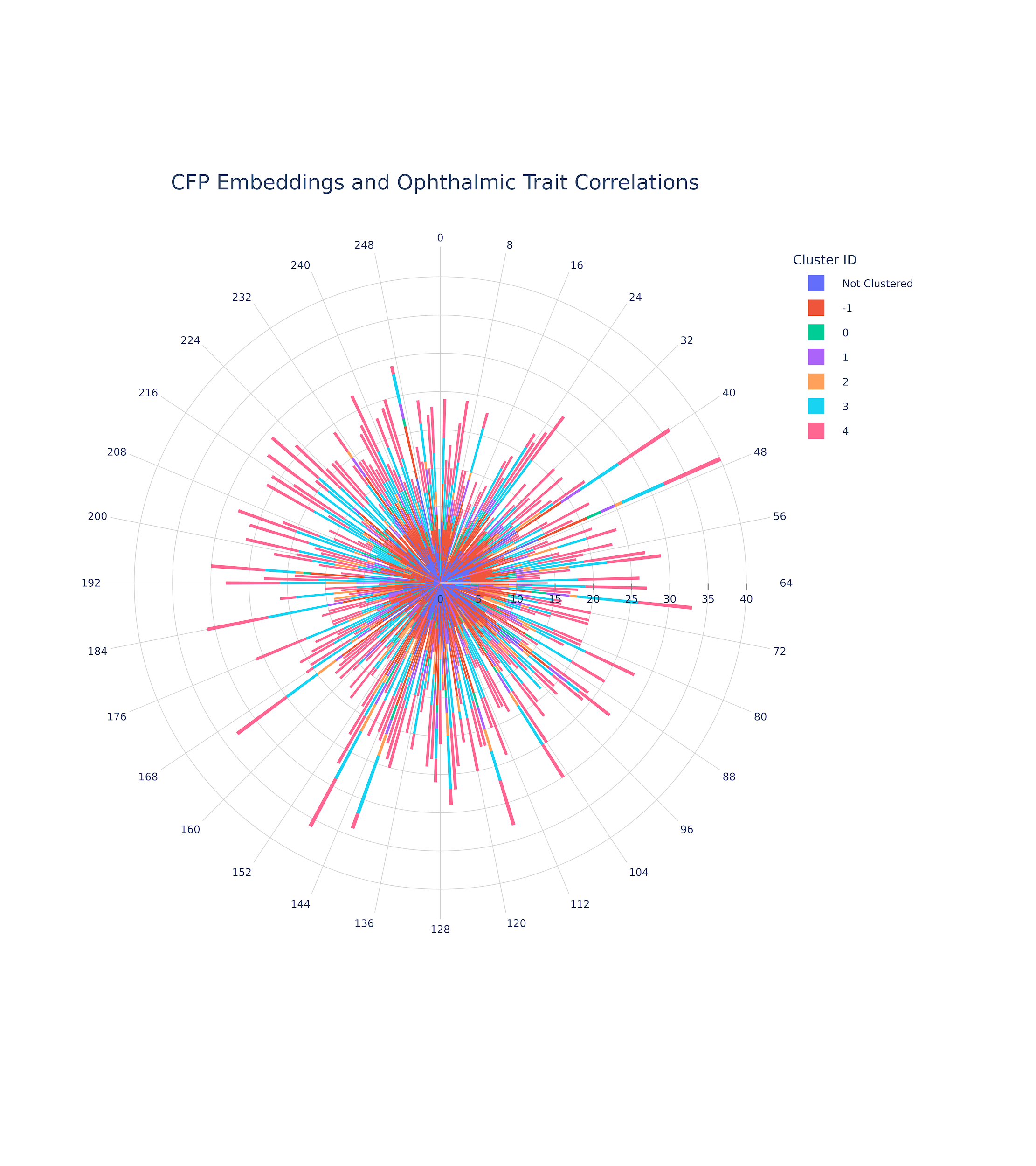
**Supplementary Figure 11:** A polar plot illustrating the relationships between ophthalmic traits and CFP embeddings. HDBSCAN identified four data clusters: ‘cluster 0’ included intraocular pressure and corneal hysteresis measures; ‘cluster 1’ included OCT derived photoreceptor layer thickness measures; ‘cluster 2’ included OCT derived thickness of the photoreceptor & retinal pigment epithelium layers; ‘cluster 3’ included a number of OCT derived thickness measures including the inner nuclear layer, outer plexiform layer, photoreceptors and retinal pigment epithelium; and ‘cluster 4’ contained OCT derived macular thickness measures. Inner retinal thickness measures (e.g. retinal nerve fibre layer and ganglion cell layer thickness). 15 traits could not be clustered due to a high level of missingness across the population. and visual acuity did not fall into HDBSCAN clusters, and were categorized as noise (cluster -1). The circumferential axis indicates the embedding number. The radial axis indicates the number of multiple testing corrected significant Pearson correlations that the embedding has with Ophthalmic traits. The bars are color coded according to the cluster of traits the significant relationship belongs to.

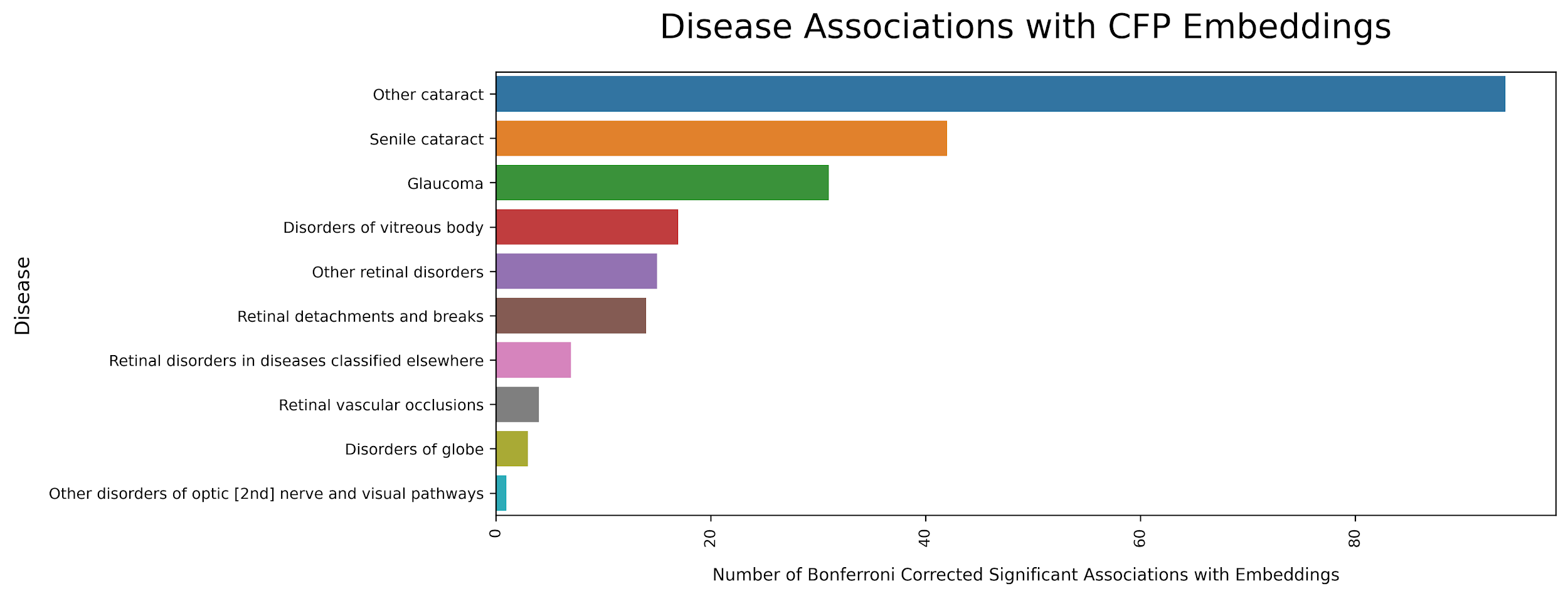
**Supplementary Figure 12:** The results of Welch’s t-test. This bar plot shows the total number of multiple testing corrected significant associations between ocular diseases and CFP embedding values. As can be seen, all ophthalmic disorders tested demonstrated associations with embeddings values. The fewest multiple testing significant results were seen for non-glaucomatous optic nerve disorders, an expected result given the retrobulbar nature of these conditions and the absence of dedicated optic nerve imaging in this study.

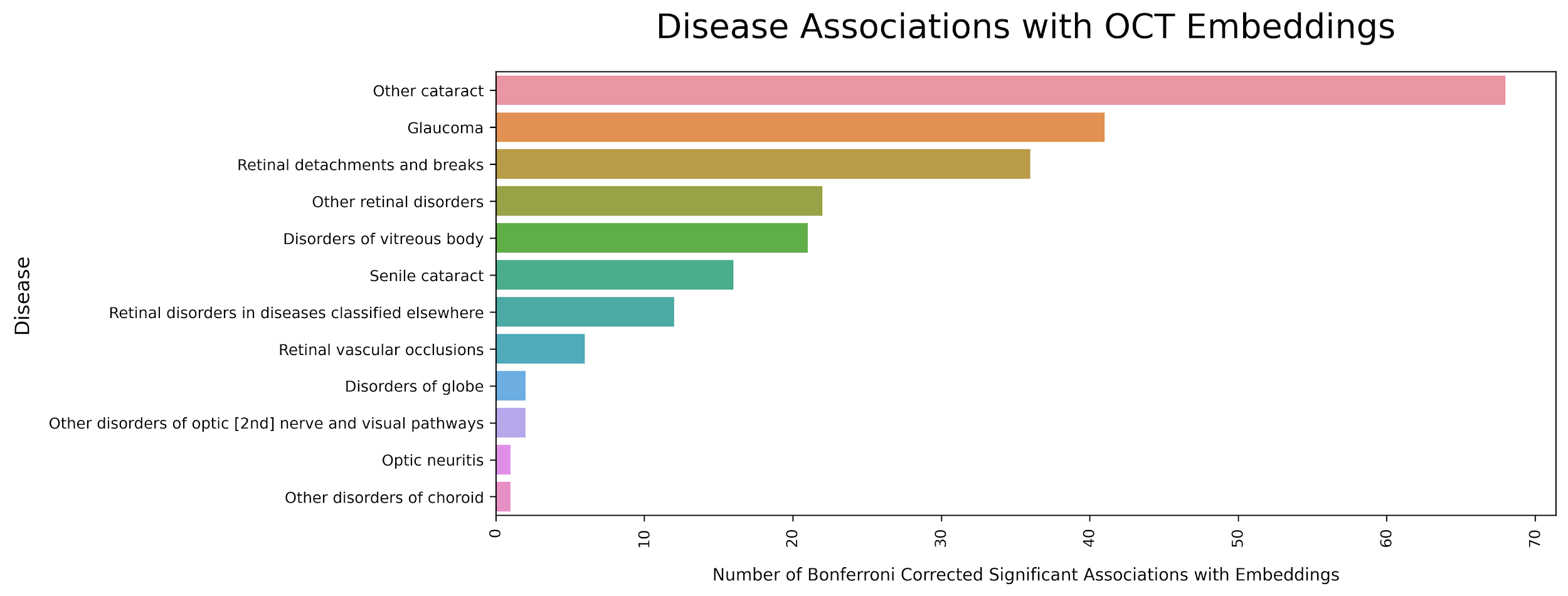
**Supplementary Figure 13:** The results of Welch’s t-test. This bar plot shows the total number of multiple testing corrected significant associations between ocular diseases and OCT embedding values. As can be seen, all ophthalmic disorders tested demonstrated associations with embeddings values. The fewest multiple testing significant results were seen for non-glaucomatous optic nerve disorders, an expected result given the retrobulbar nature of these conditions and the absence of dedicated optic nerve imaging in this study.

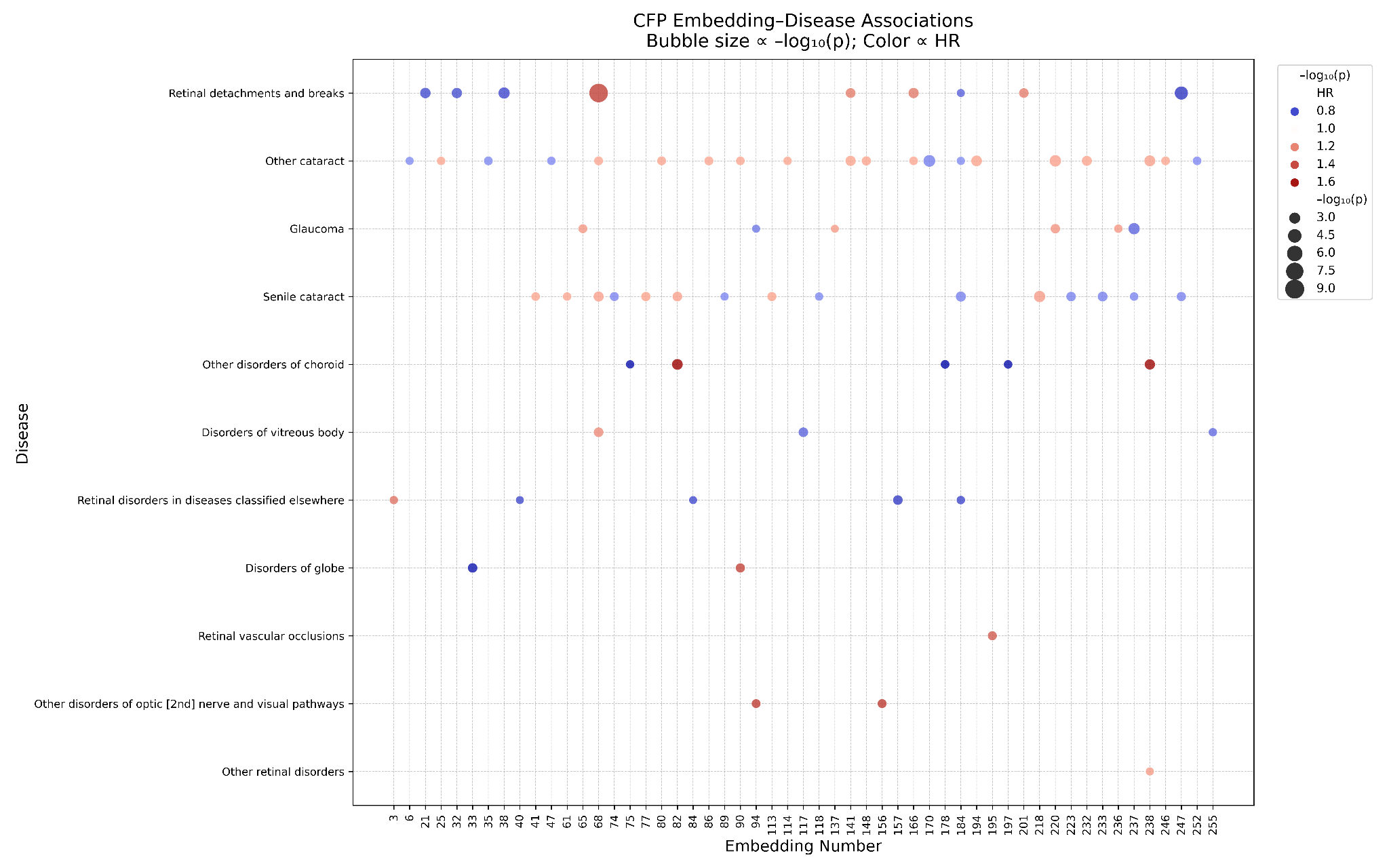
**Supplementary Figure 14:** A bubble plot illustrating the results of our CFP-ocular disease Cox Proportional Hazards analysis. Only embeddings and disorders with at least one multiple testing corrected significant result are plotted here. The size of the bubbles indicates the level of significance (larger bubbles = more significant). The bubbles are color coded according to the hazards ratio (HR).

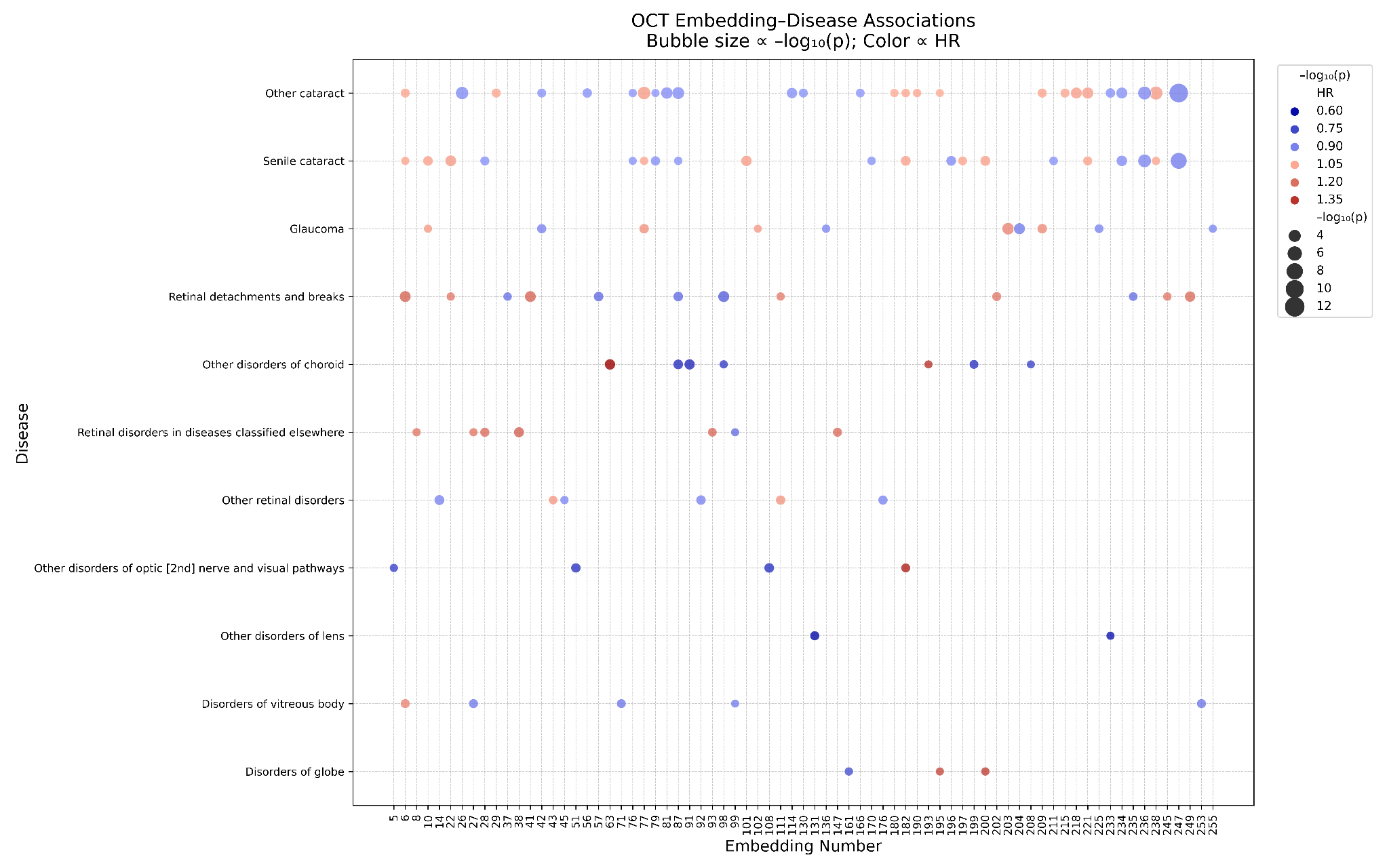

**Supplementary Figure 15:** A bubble plot illustrating the results of our OCT-ocular disease Cox Proportional Hazards analysis. Only embeddings and disorders with at least one multiple testing corrected significant result are plotted here. The size of the bubbles indicates the level of significance (larger bubbles = more significant). The bubbles are color coded according to the hazards ratio (HR).

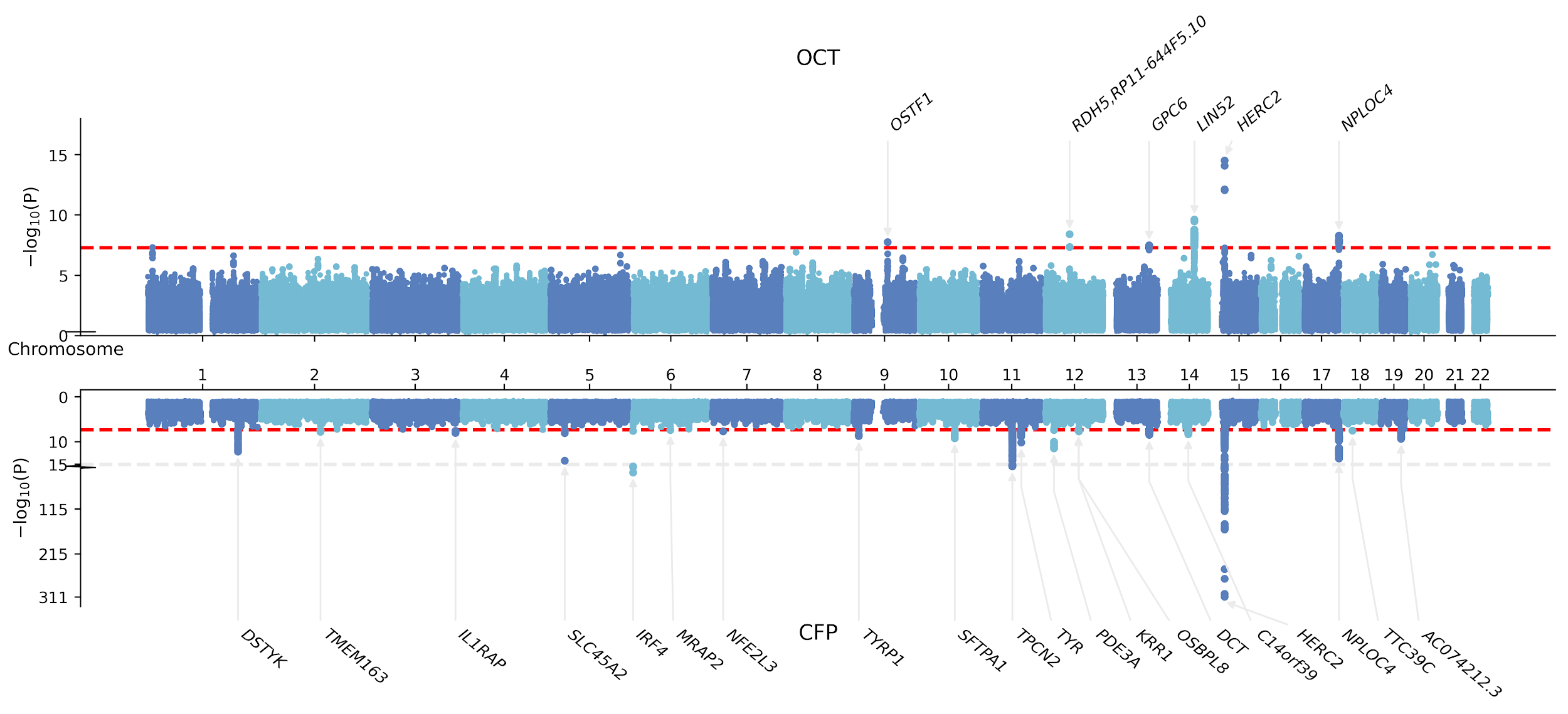

**Supplementary Figure 16:** A Miami plot of GWAS results for both CFP and OCT embeddings. In the interest of creating a single illustrative figure, this Miami plot contains the top SNPs from all embeddings identified to have a polygenic architecture using LD score regression. The superior plot is for OCT-derived embedding associations, whilst the inferior plot is for CFP-derived embedding associations. Note that due to the asymmetry in the axis, the CFP plot has been truncated, with the point of truncation indicated on the y axis. The red line indicates the genome-wide significance threshold, and all annotated variants exceed this threshold.

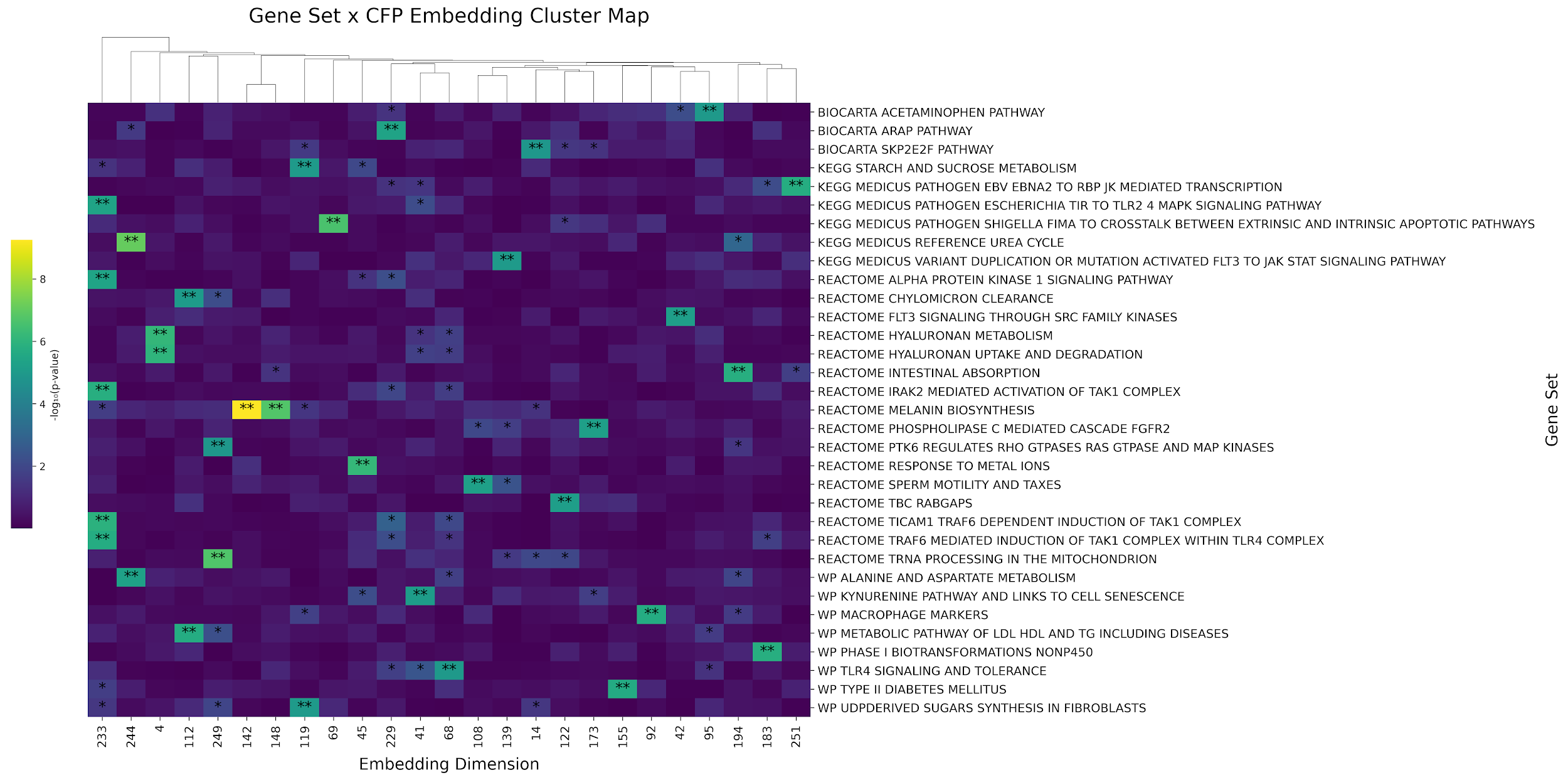

**Supplementary Figure 17:** A cluster plot of MAGMA results for our CFP-derived embedding analysis. A single asterisk indicates nominal significance, whilst a double asterisk indicates multiple testing corrected significance. The figure contains gene sets that did not replicate in the validation study (i.e. these are discovery results). Only gene sets and embeddings with at least one multiple testing significant results are plotted.

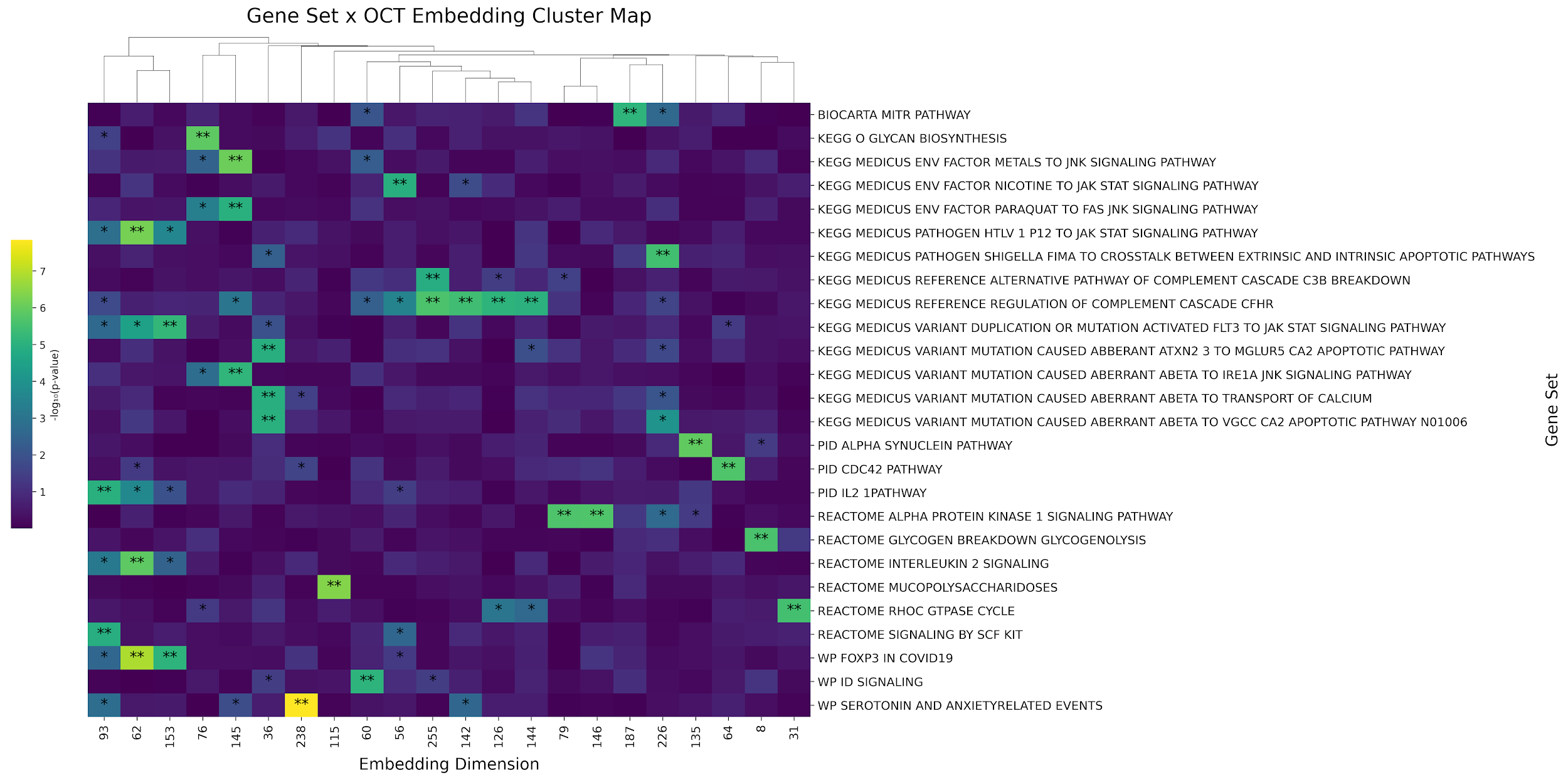

**Supplementary Figure 18:** A cluster plot of MAGMA results for our OCT-derived embedding analysis. A single asterisk indicates nominal significance, whilst a double asterisk indicates multiple testing corrected significance. The figure contains gene sets that did not replicate in the validation study (i.e. these are discovery results). Only gene sets and embeddings with at least one multiple testing significant results are plotted.

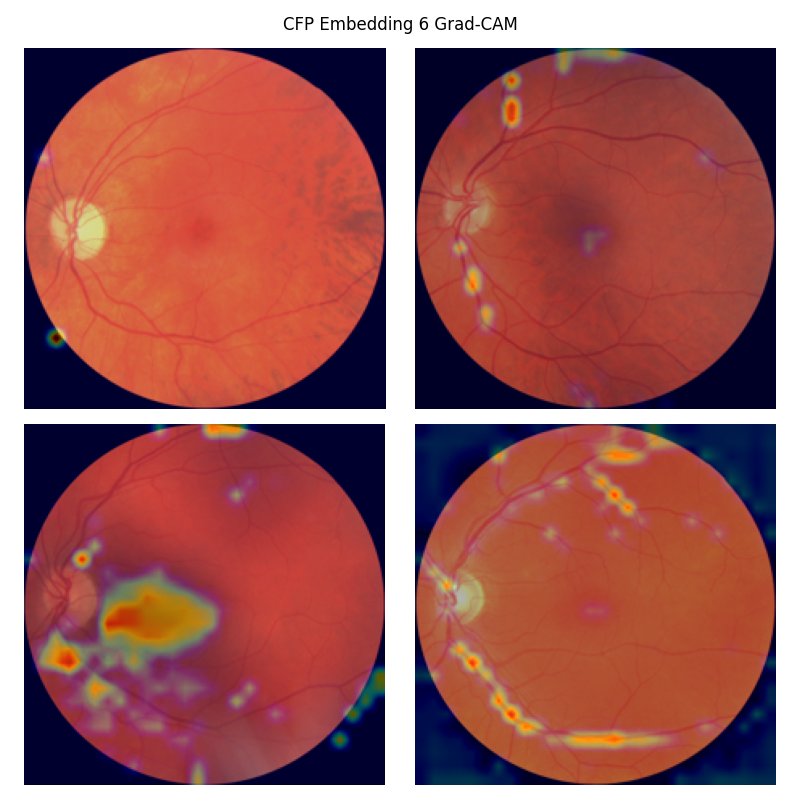
**Supplementary Figure 19:** Grad-CAM saliency map for CFP embedding 6. This was the second most strongly associated with future hypertension risk. The embedding seems to localise to the vasculature and projection artifacts. Reproduced with the permission of UK Biobank.

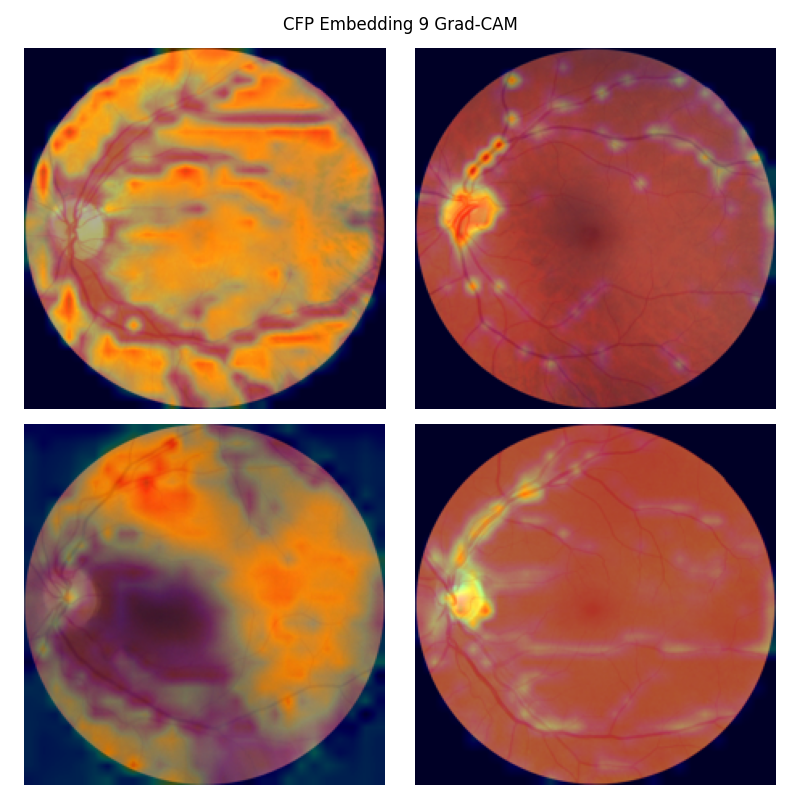
**Supplementary Figure 20:** Grad-CAM saliency map for CFP embedding 9. This was the embedding most strongly associated with heart failure at the time of imaging (baseline). The embedding seems to represent the vasculature, and perhaps to some extent the fundal pigmentation or choroidal features (given the background feature saliency). Reproduced with the permission of UK Biobank.
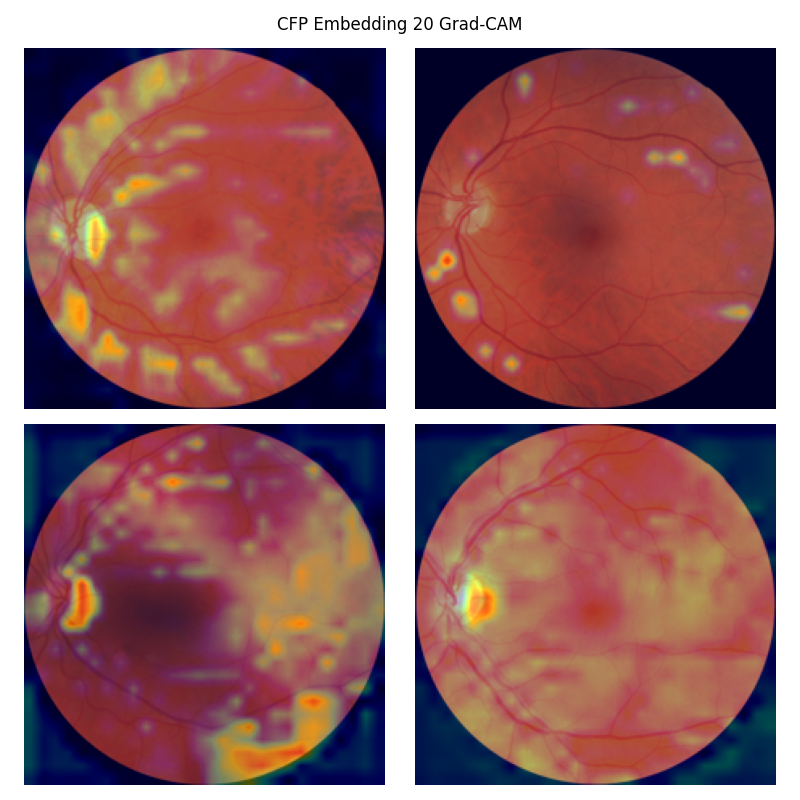
**Supplementary Figure 21:** Grad-CAM saliency map for CFP embedding 20. This was the embedding most strongly associated with chronic ischaemic heart disease at the time of imaging (baseline). The embedding seems to represent the optic nerve head, as well as some scattered background features, which we speculate could be the choroidal vasculature. Reproduced with the permission of UK Biobank.

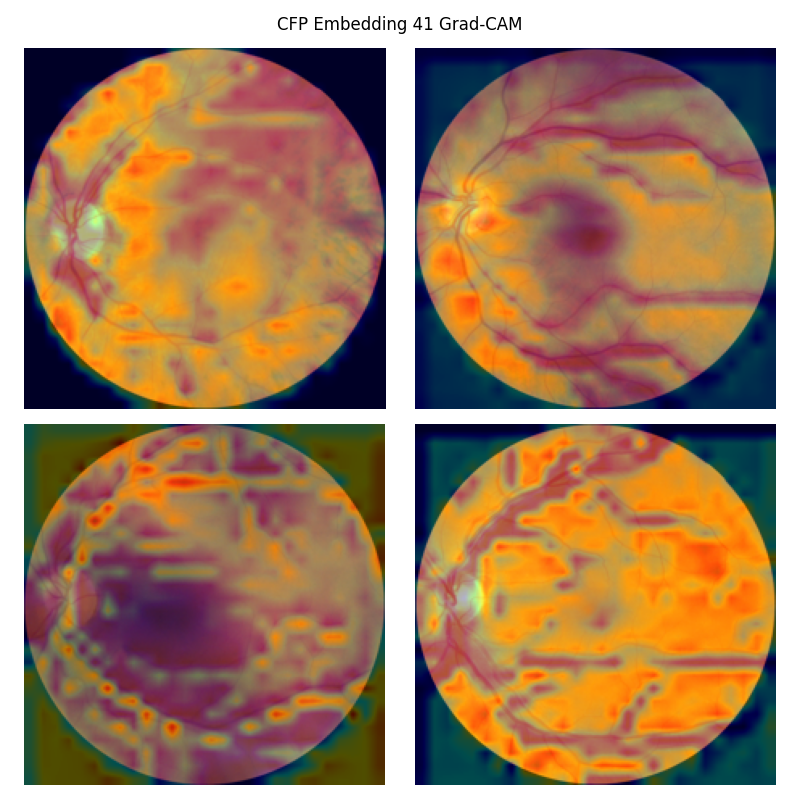

**Supplementary Figure 22:** Grad-CAM saliency map for CFP embedding 41. This was the CFP feature most strongly associated with the gene set ‘WikiPathways kynurenine pathway and links to cell senescence’. As can be seen, the embedding has a diffuse saliency map, and it is not clear what specific trait is represented by the latent feature. Whilst some maps correspond to the vasculature (bottom left), others correspond to background fundal features. Reproduced with the permission of UK Biobank.

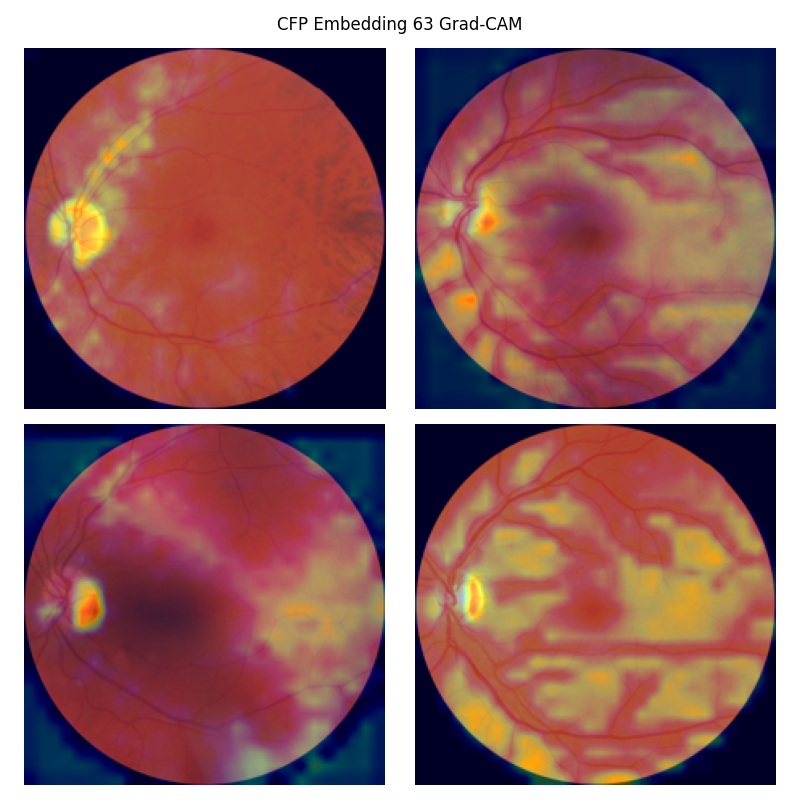
**Supplementary Figure 23:** Grad-CAM saliency map for CFP embedding 63. This was the embedding most strongly associated with acute myocardial infarction at the time of imaging (baseline). As can be seen, the embedding seems to localise to the optic nerve head, with some more inconsistent signals around the vasculature and more diffusely throughout the background. Reproduced with the permission of UK Biobank.

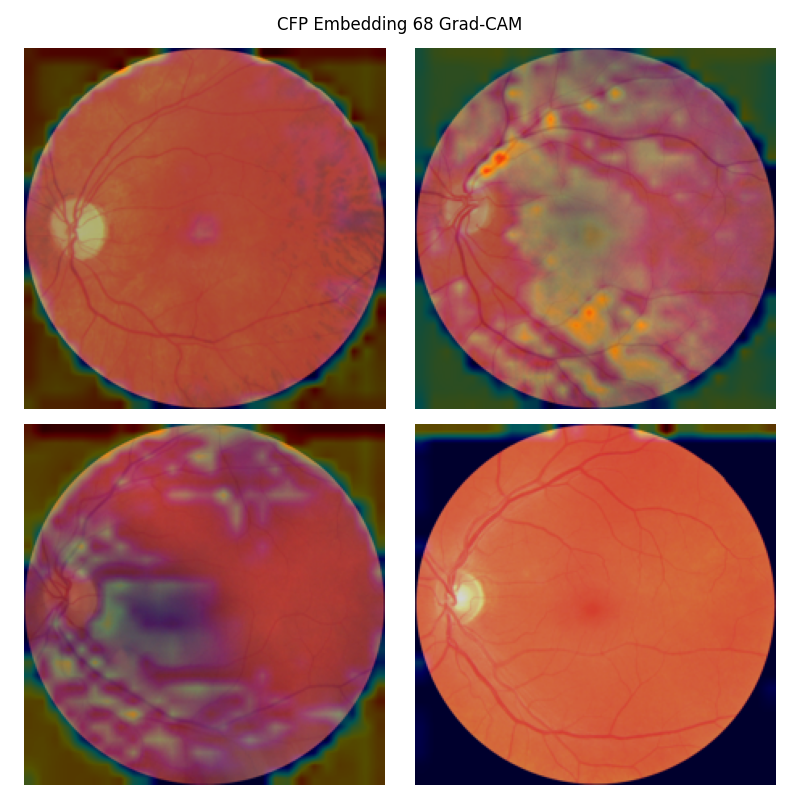
**Supplementary Figure 24:** Grad-CAM saliency map for CFP embedding 68. This embedding was genetically correlated with heart failure (nominally). The feature appears to localise to the vasculature. Reproduced with the permission of UK Biobank.

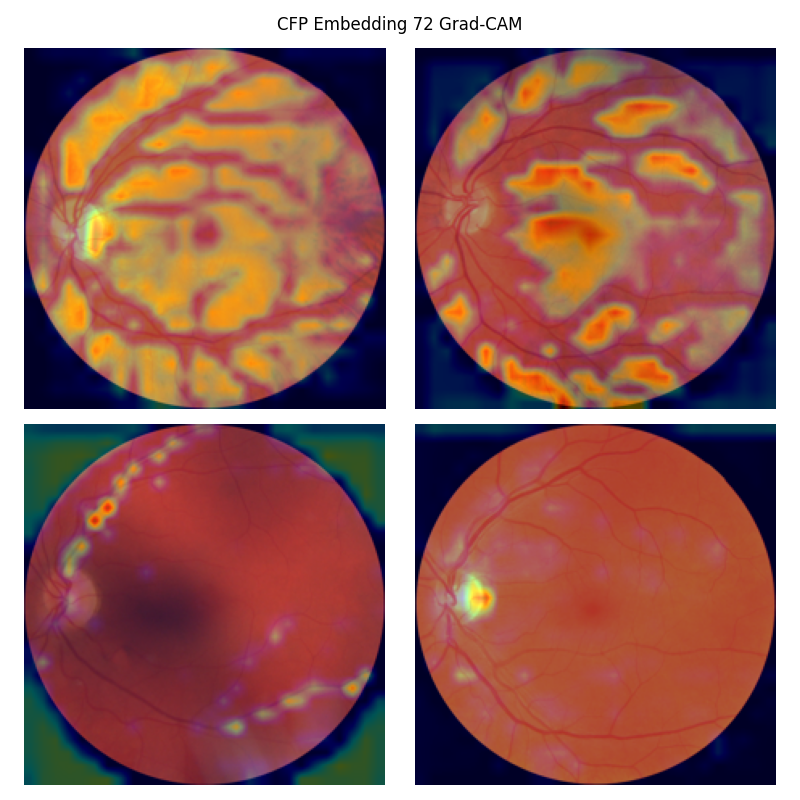
**Supplementary Figure 25:** Grad-CAM saliency map for CFP embedding 72. This embedding was genetically correlated with myocardial infarction (nominally). The feature appears to localise to the vasculature and the optic nerve head. Reproduced with the permission of UK Biobank.

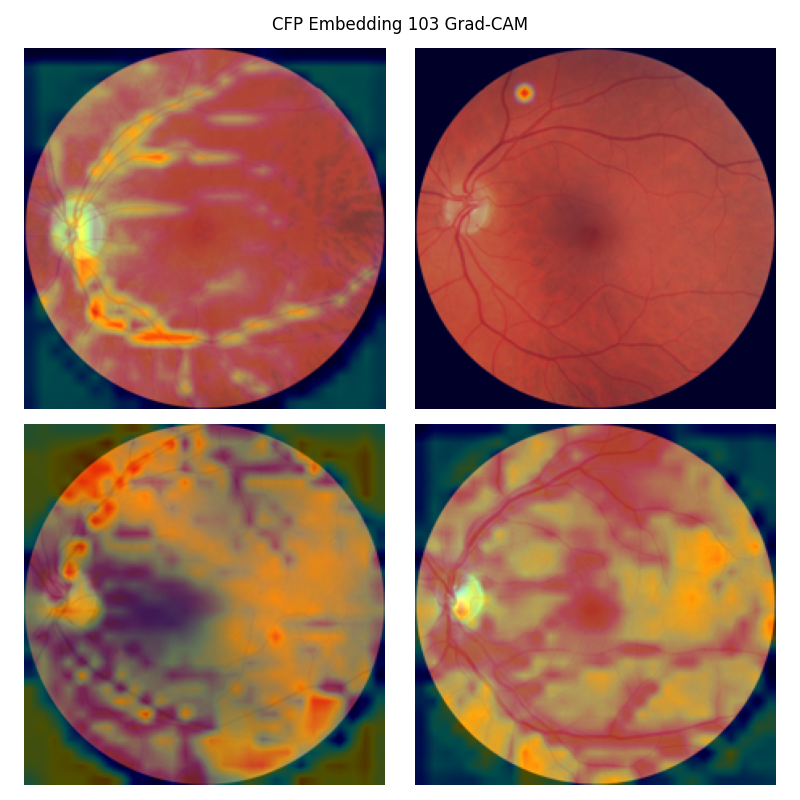
**Supplementary Figure 26:** Grad-CAM saliency map for CFP embedding 103. This was the CFP feature second most strongly associated with future Alzheimer’s disease risk. The embedding seems to localise several features including the optic nerve head, vasculature, and to patchy background features. Reproduced with the permission of UK Biobank.

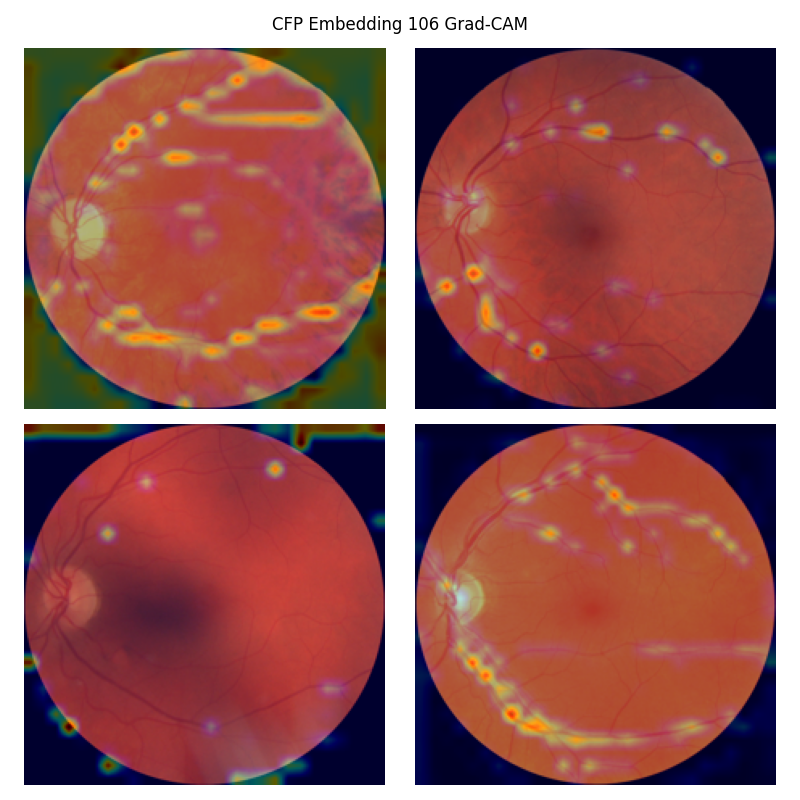
**Supplementary Figure 27:** Grad-CAM saliency map for CFP embedding 106. This embedding was associated with a large number of cardiovascular features in our Pearson correlation analysis. The feature appears to localise to the vascular tree. Reproduced with the permission of UK Biobank.

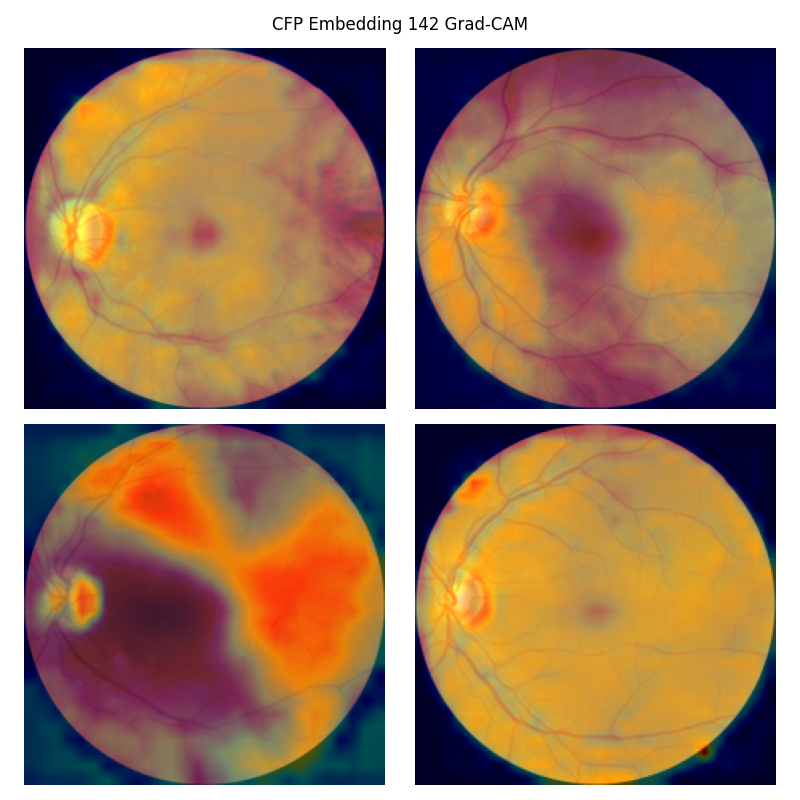

**Supplementary Figure 28:** Grad-CAM saliency map for CFP embedding 142. This was the CFP feature most strongly associated with the gene set for melanin biosynthesis and interestingly was also the embedding which dominated the correlation with lipids in our CCA analysis. The embedding seems to represent the background features in a relatively homogeneous manner and therefore appears to correspond to the extent of fundal pigmentation. Reproduced with the permission of UK Biobank.
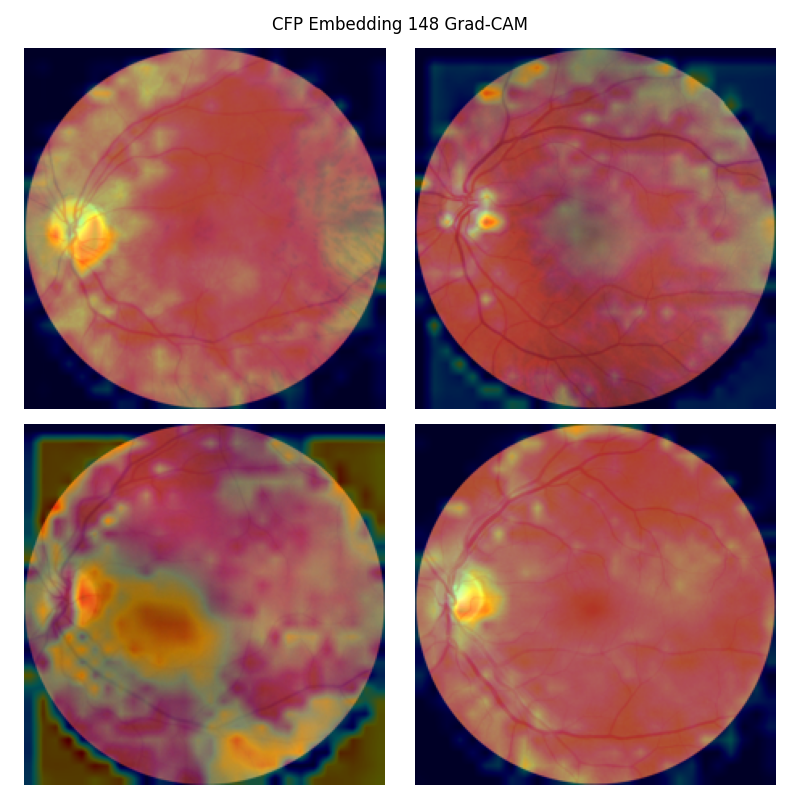
**Supplementary Figure 29:** Grad-CAM saliency map for CFP embedding 148. This was the embedding most strongly associated with hypertension and Parkinson’s disease at the time of imaging (baseline). This was the embedding most strongly associated with future risk of hypertension. Our radiomic analysis revealed this embedding was associated with a large number of cerebral volumes and diffusion MRI features. The genetic analyses revealed this embedding was associated with melanin biosynthesis. The embedding appears to localise to the optic nerve head, pigmented areas, and the projection artifacts. Reproduced with the permission of UK Biobank.
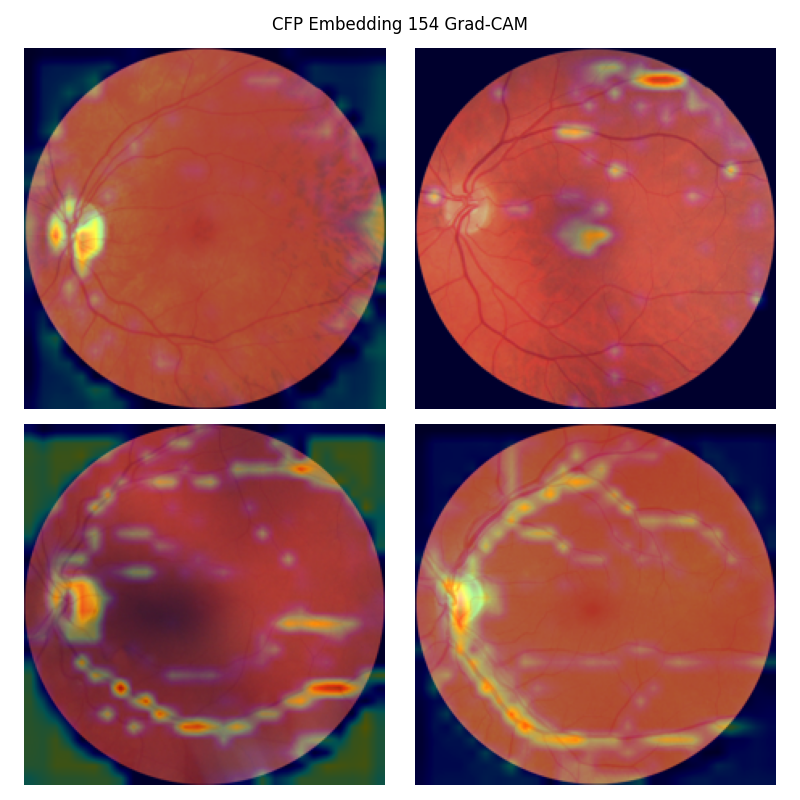
**Supplementary Figure 30:** Grad-CAM saliency map for CFP embedding 154. This was the embedding most strongly associated with transient ischaemic attack at the time of imaging (baseline). The embedding seems to localise to the optic nerve head and vasculature. Reproduced with the permission of UK Biobank.

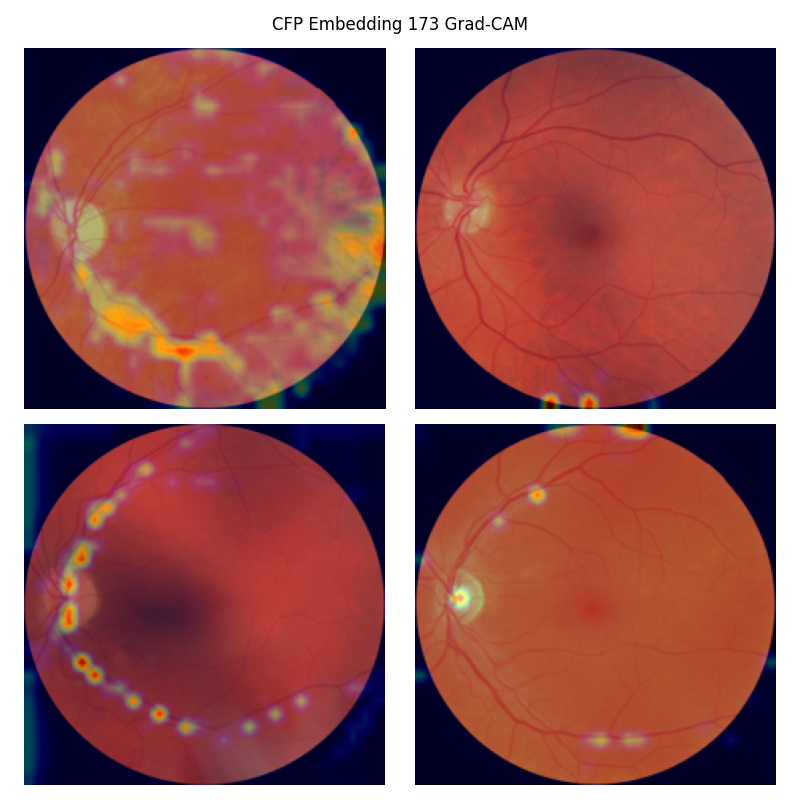
**Supplementary Figure 31:** Grad-CAM saliency map for CFP embedding 173. This was the CFP feature most strongly associated with the gene set ‘Reactome phospholipase C mediated cascade *FGFR2’*. The embedding seems to localise to the vasculature. Reproduced with the permission of UK Biobank.

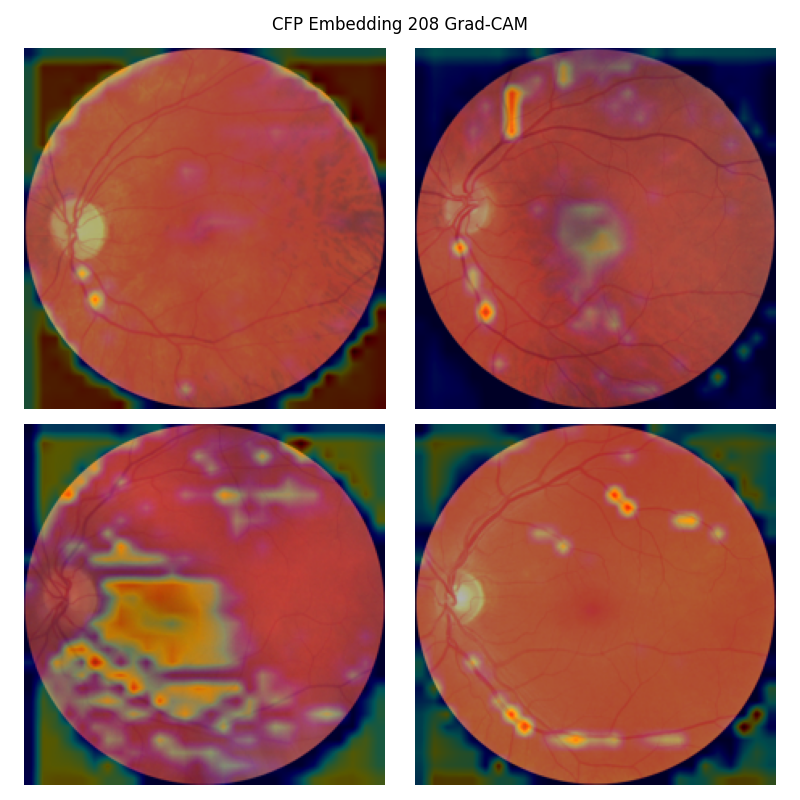

**Supplementary Figure 32:** Grad-CAM saliency map for CFP embedding 208. This was the third most strongly associated with future hypertension risk. The embedding seems to localise to the vasculature and the projection artifact in the bottom left image. Reproduced with the permission of UK Biobank.
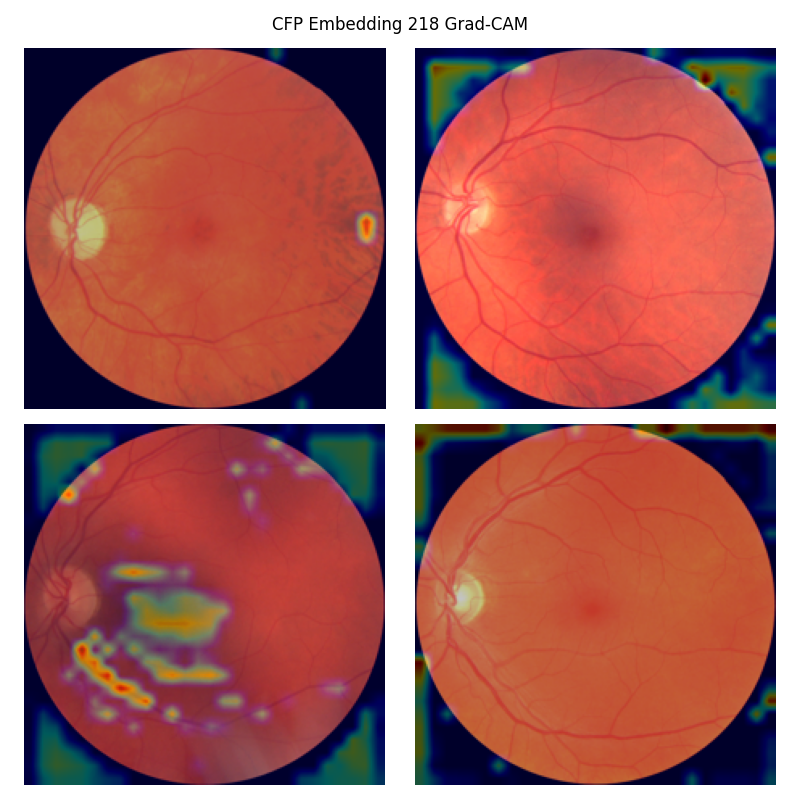

**Supplementary Figure 33:** Grad-CAM saliency map for CFP embedding 218. This was the embedding most strongly associated with angina at the time of imaging (baseline). The embedding seems to localise to the vasculature. Reproduced with the permission of UK Biobank.

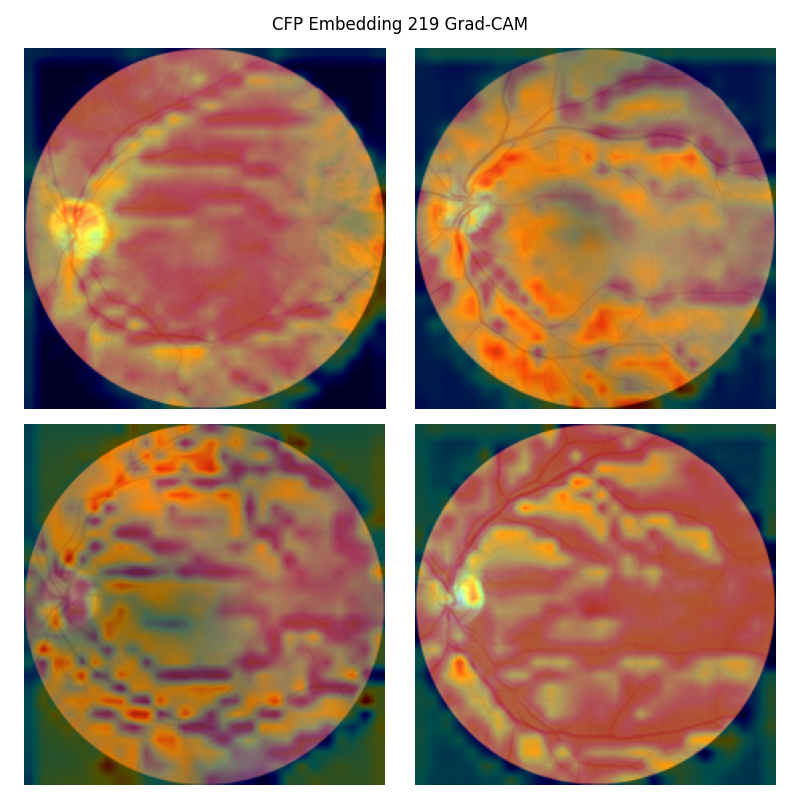

**Supplementary Figure 34:** Grad-CAM saliency map for CFP embedding 219. This embedding was associated with future risk of dementia in diseases classified elsewhere (inclusive of Pick disease, Creutzfeldt-Jakob disease, Parkinson’s disease, Huntington disease, human immunodeficiency virus and others). This embedding appears to localise principally to perivascular regions. Reproduced with the permission of UK Biobank.

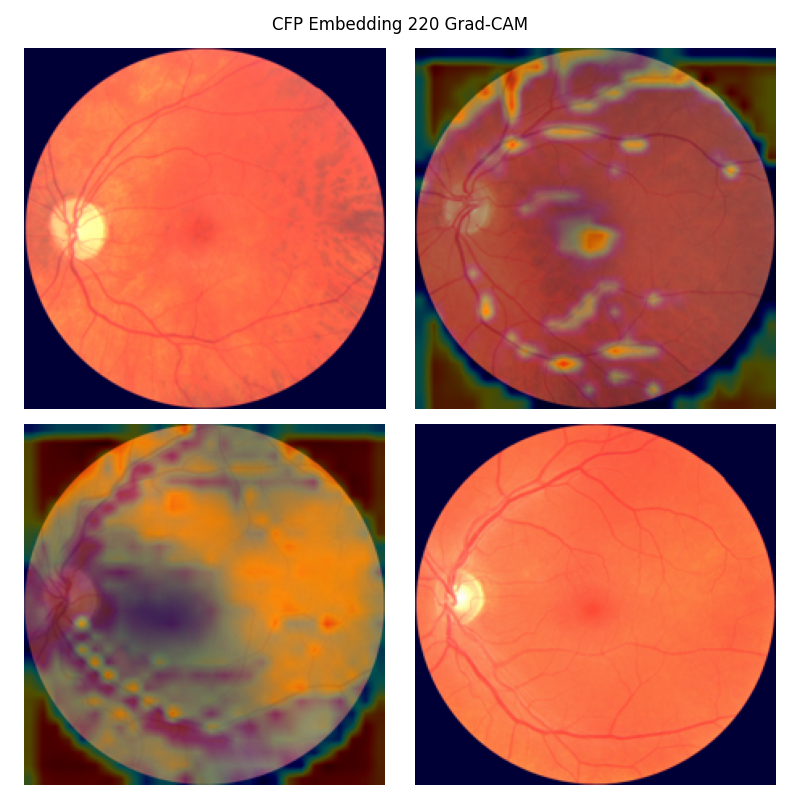

**Supplementary Figure 35:** Grad-CAM saliency map for CFP embedding 220. This embedding was genetically correlated with myocardial infarction. The feature appears to localise to vascular structures and perhaps the fovea.

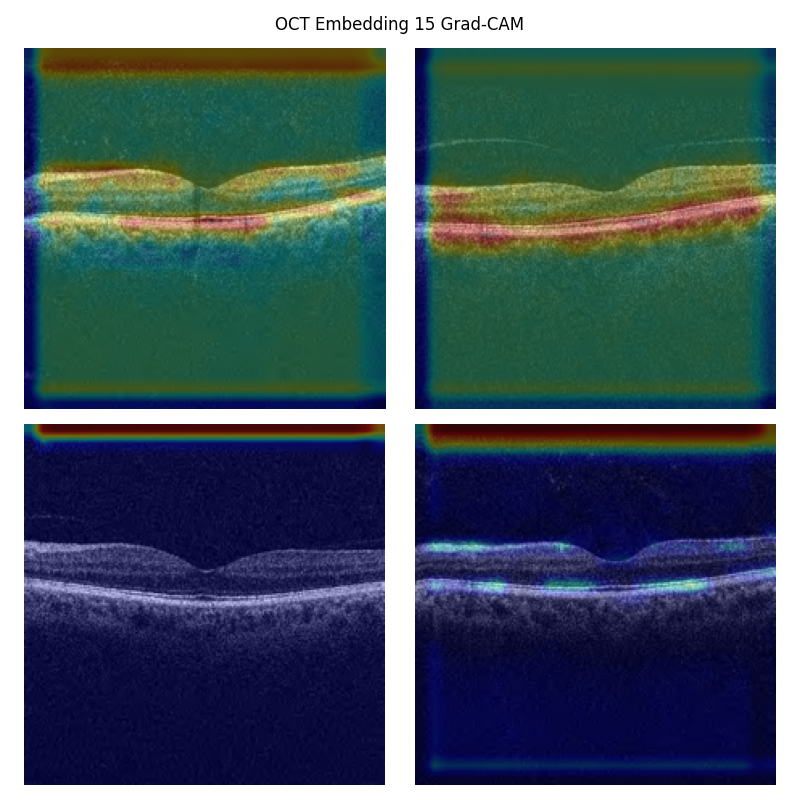
**Supplementary Figure 36:** Grad-CAM saliency map for OCT embedding 15. This embedding was the OCT embedding most strongly associated with baseline Parkinson’s disease. The second convolutional layer was targeted due to poor localisation in the third convolution. The embedding appears to localise to the ellipsoid zone (the location of photoreceptors). Reproduced with the permission of UK Biobank.

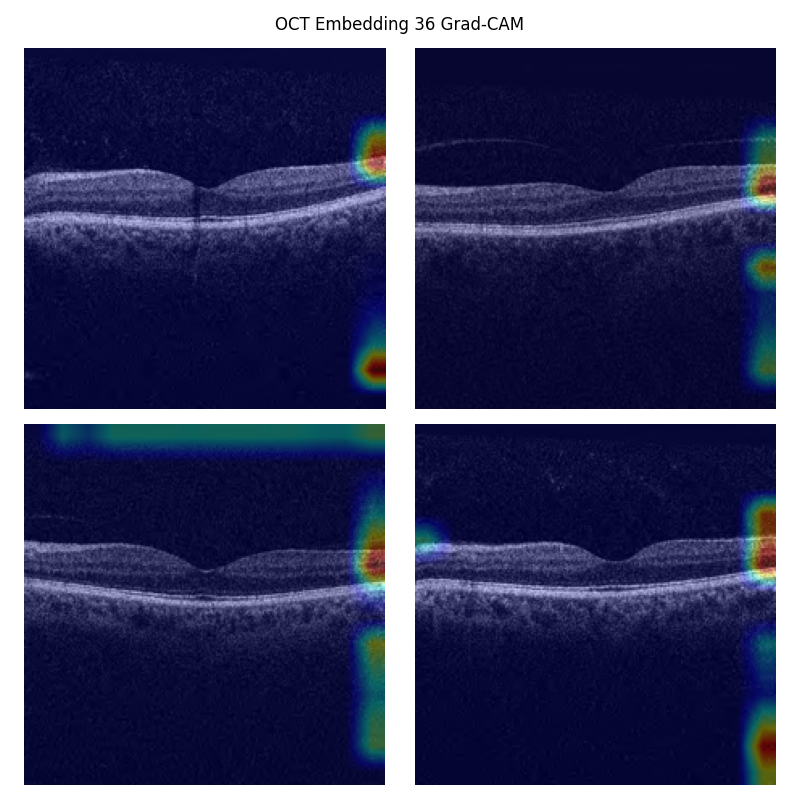
**Supplementary Figure 37:** Grad-CAM saliency map for OCT embedding 36. This was the OCT feature most strongly associated with dementia-related gene sets. The embedding does not clearly localise to a specific ophthalmic feature in Grad-CAM. Reproduced with the permission of UK Biobank.

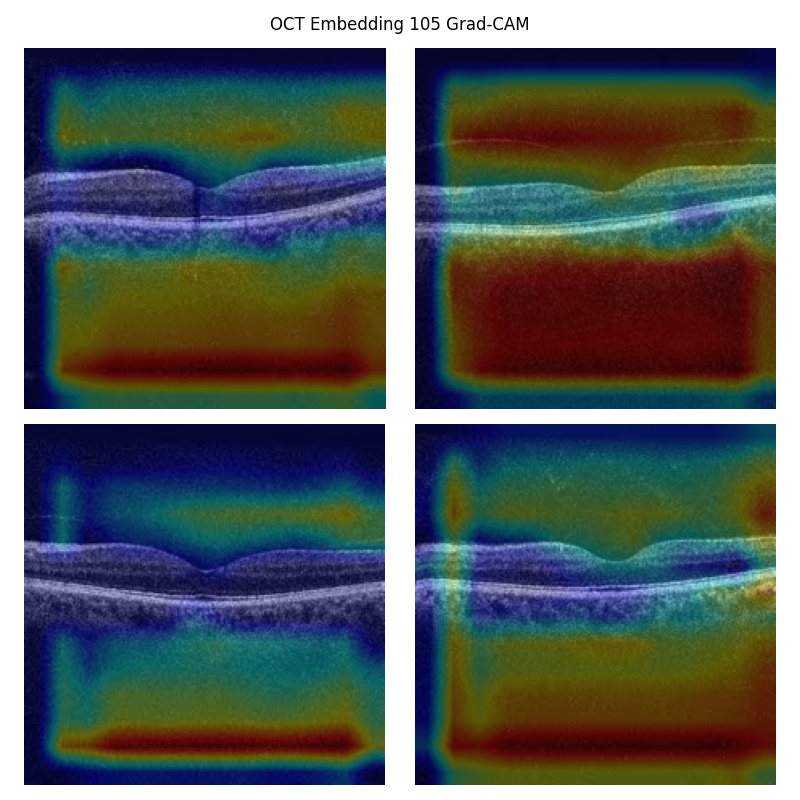
**Supplementary Figure 38:** Grad-CAM saliency map for OCT embedding 105. This was the OCT feature most strongly associated with future hypertension. As can be seen, the embedding seems to localise to superior and inferior borders of the retina, and so perhaps represents total macula thickness. Reproduced with the permission of UK Biobank.

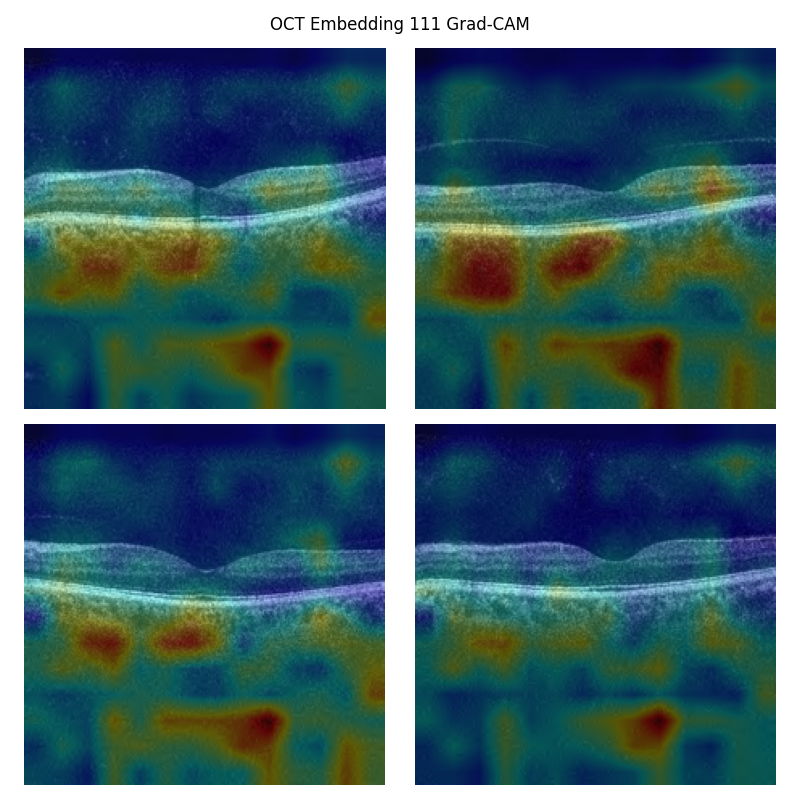
**Supplementary Figure 39:** Grad-CAM saliency map for OCT embedding 111. This embedding was genetically correlated with heart failure (nominally). Grad-CAM failed to localise the feature, and so Layer-CAM was used. The feature appears to localise to choroid. Reproduced with the permission of UK Biobank.

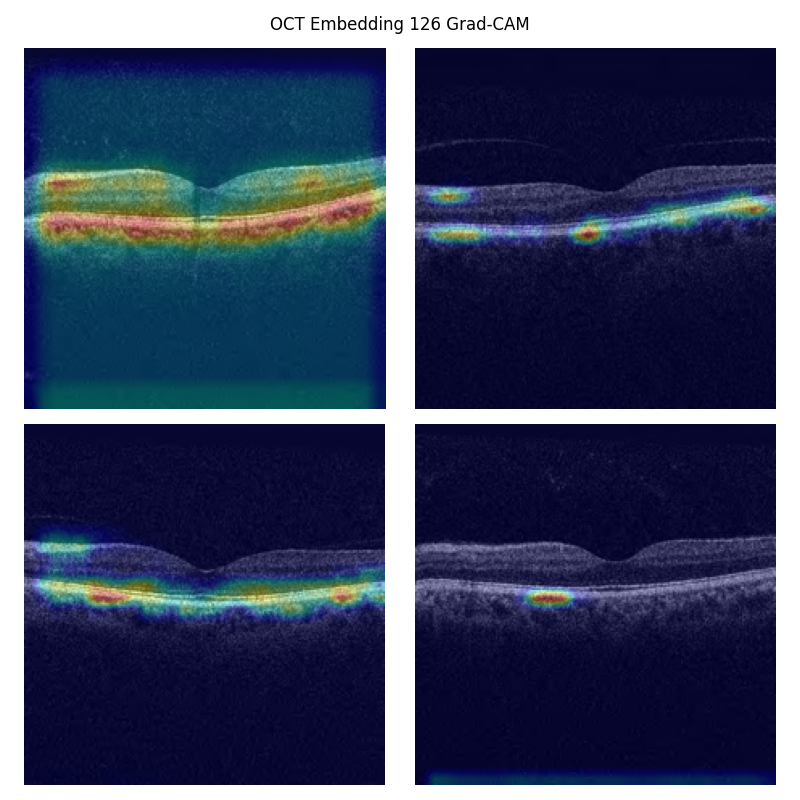
**Supplementary Figure 40:** Grad-CAM saliency map for OCT embedding 126. This was the OCT feature most strongly associated with the gene set ‘KEGG medicus reference regulation of complement cascade *CFHR’*. The third convolutional layer saliency map did not localise clearly, therefore this analysis was performed on the second convolutional layer. The embedding appears to localise to the choroid and retinal pigment epithelium, in keeping with previous work linking complement to disorders of these structures. Reproduced with the permission of UK Biobank.

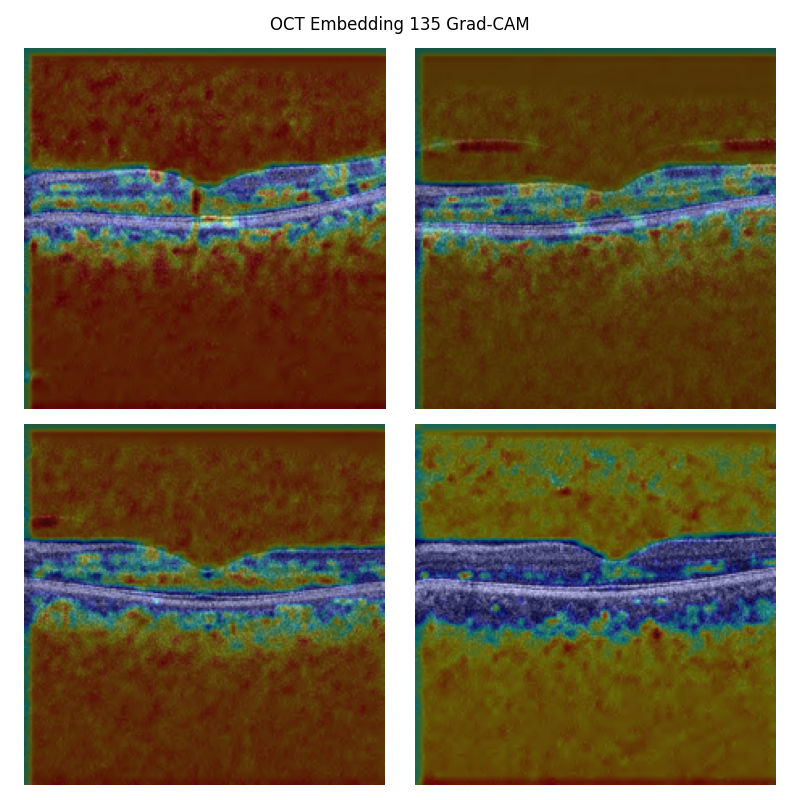
**Supplementary Figure 41:** Grad-CAM saliency map for OCT embedding 135. This was the OCT feature most strongly associated with the gene set ‘Pathway Interaction Database (PID) alpha synuclein pathway*’*. The third and second convolutional layer saliency maps did not localise to any specific area and therefore this analysis was performed on the first convolutional layer. The embedding appears to loosely localise to the outer retinal layers. Reproduced with the permission of UK Biobank.

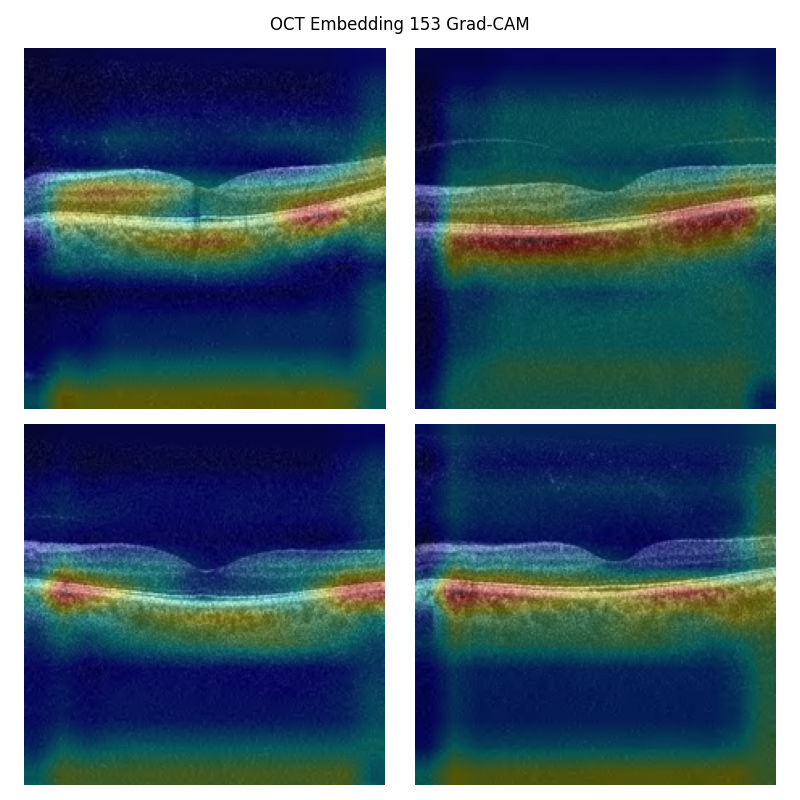

**Supplementary Figure 42:** Grad-CAM saliency map for OCT embedding 153. This was the OCT feature most strongly associated with the gene set ‘KEGG medicus variant duplication or mutation activated *FLT3* to Jak-STAT signaling pathway*’*. The embedding appears to localise to the choroid. Reproduced with the permission of UK Biobank.

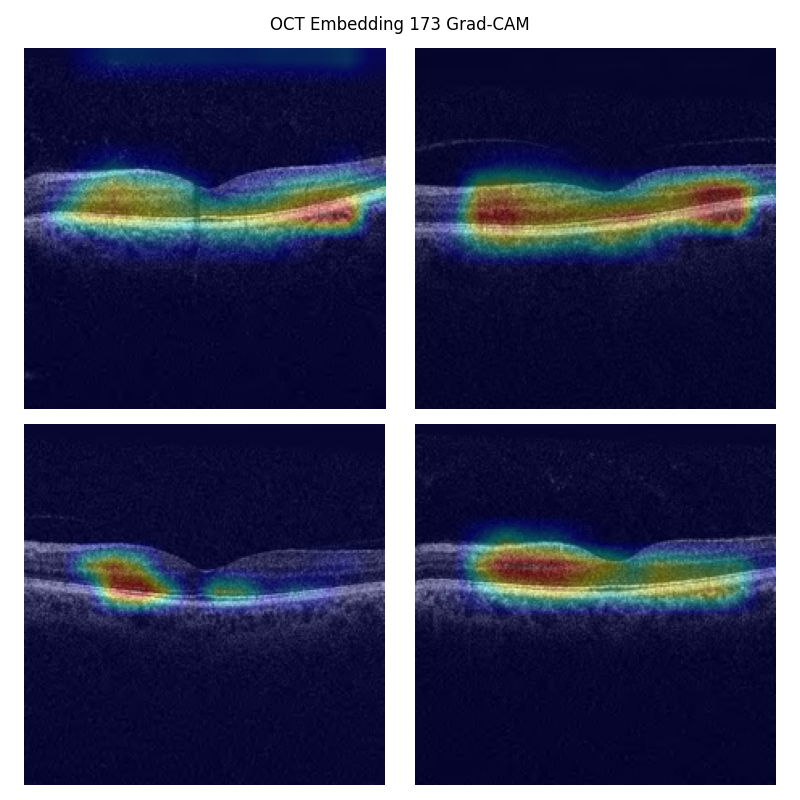
**Supplementary Figure 43:** Grad-CAM saliency map for OCT embedding 173. This was the embedding most strongly associated with acute myocardial infarction at the time of imaging (baseline). The embedding appears to localise to the neurosensory retina. Reproduced with the permission of UK Biobank.

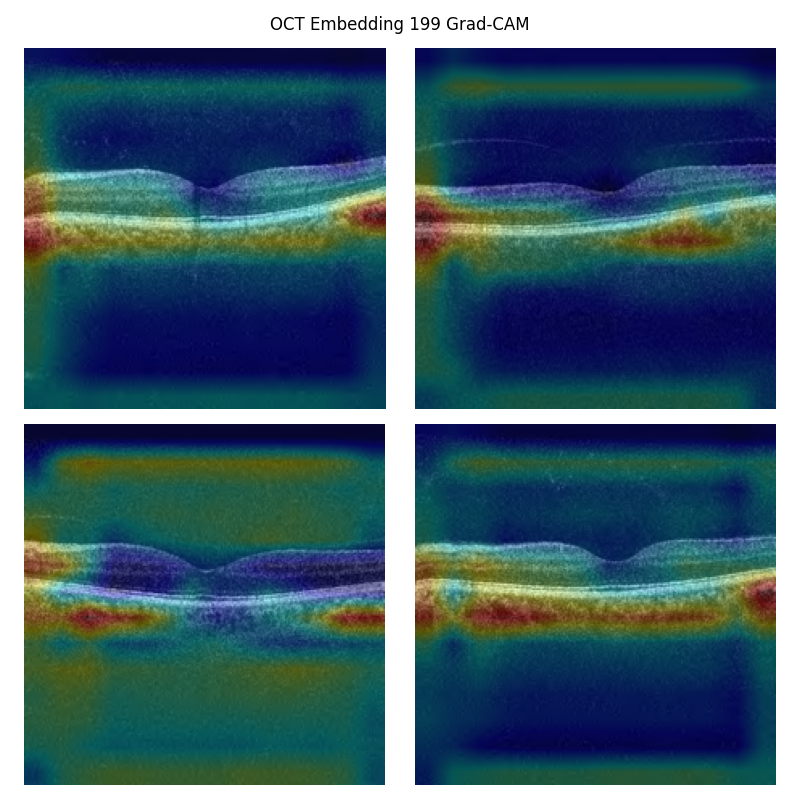
**Supplementary Figure 44:** Grad-CAM saliency map for OCT embedding 199. This embedding was associated with a large number of cardiovascular features in our Pearson correlation analysis. The feature appears to localise to the choroid. Reproduced with the permission of UK Biobank.

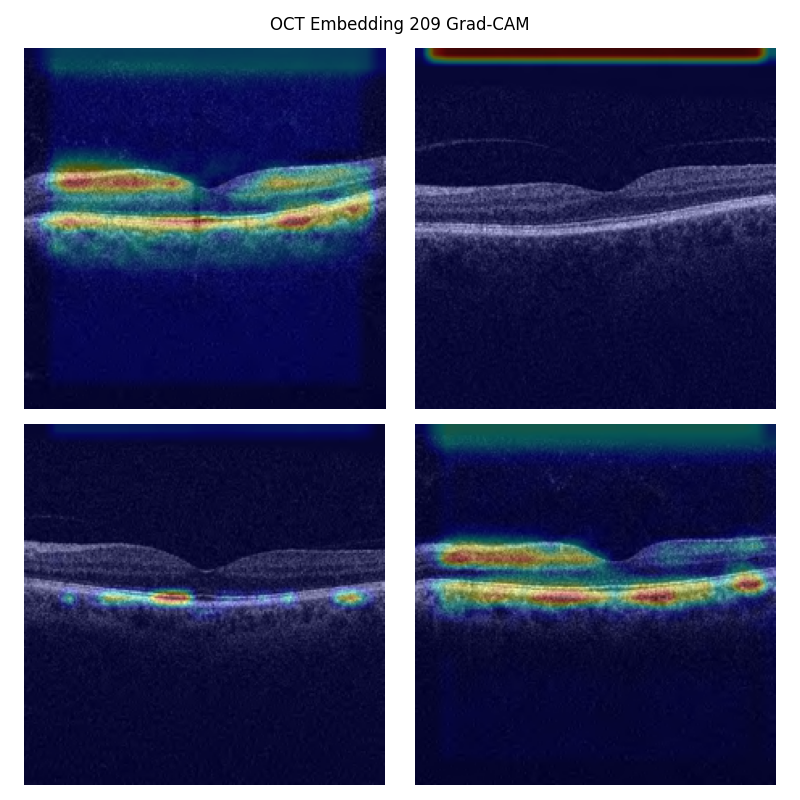
**Supplementary Figure 45:** Grad-CAM saliency map for OCT embedding 209. This was the OCT feature most strongly associated with baseline hypertension, angina, and chronic ischaemic heart disease, and future heart failure. The embedding appears to localise to elipsoid zone, the retinal pigmental epithelium, retinal nerve fibre layer, the ganglion cell layer, and to some extent the choroid. Reproduced with the permission of UK Biobank.

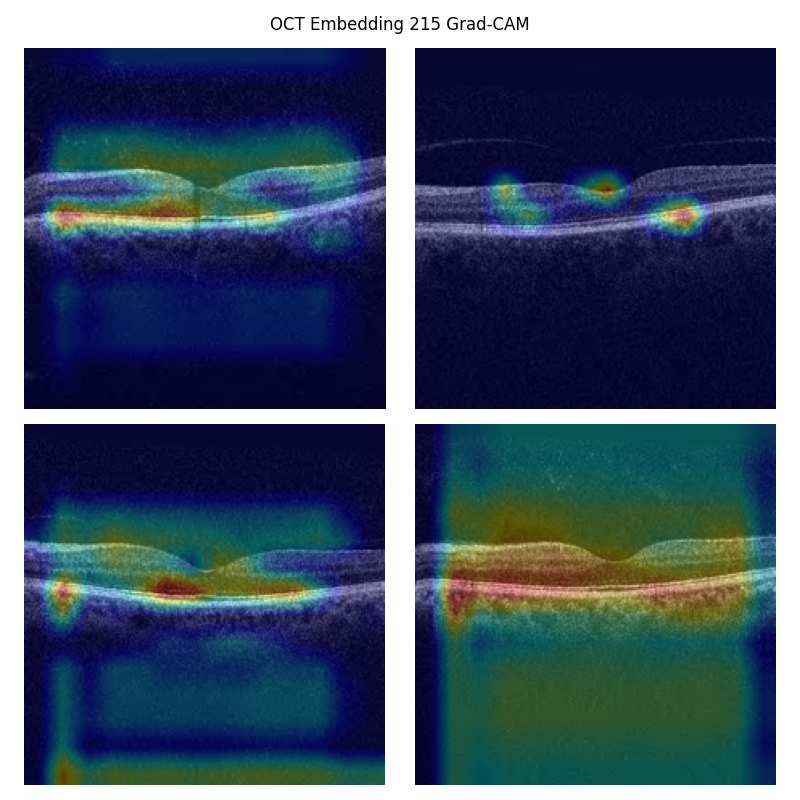

**Supplementary Figure 46:** Grad-CAM saliency map for OCT embedding 215. This embedding was associated with a large number of cerebral radiomic features in our Pearson correlation analysis. The feature is poorly localised. Reproduced with the permission of UK Biobank.

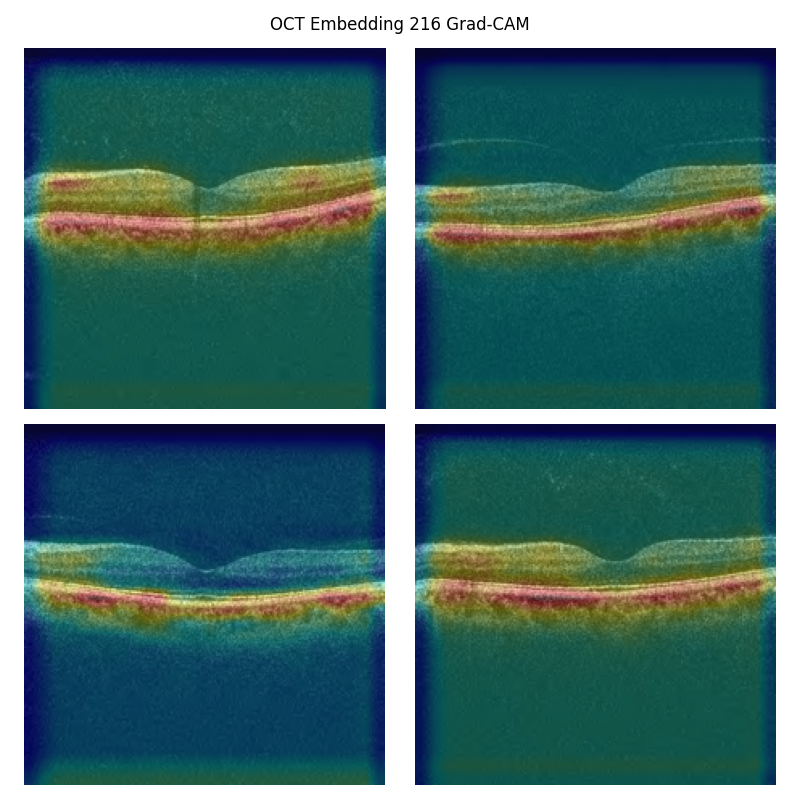

**Supplementary Figure 47:** Grad-CAM saliency map for OCT embedding 216. This was the leading embedding in the embedding-metabolome CCA analysis (i.e., the strongest weighted in relation to lipid measures). The third convolutional layer saliency map did not localise clearly, therefore this analysis was performed on the second convolutional layer. The feature appears to localise to the choroid. Reproduced with the permission of UK Biobank.

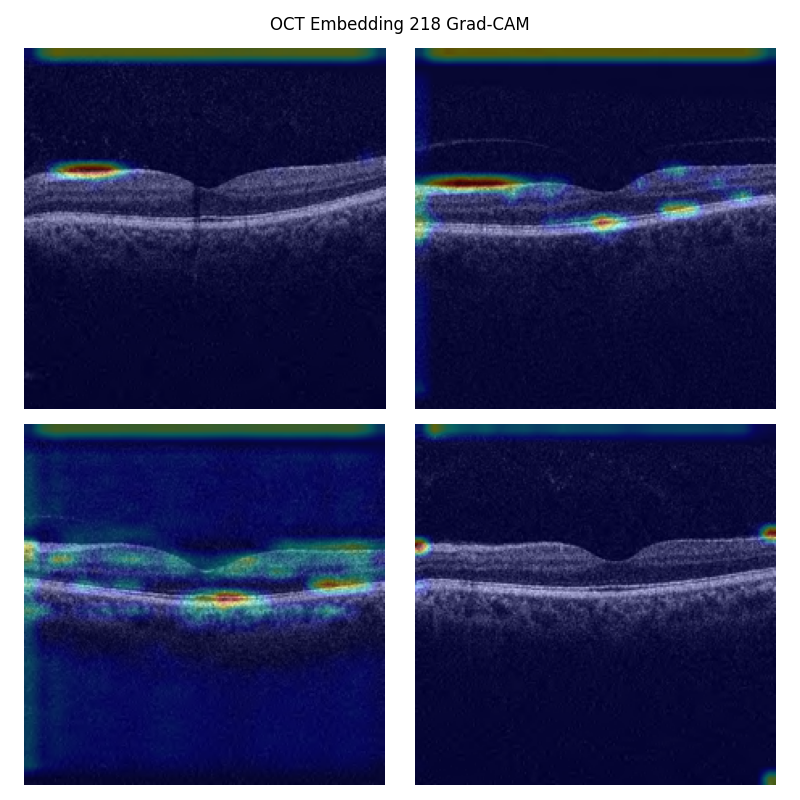
**Supplementary Figure 48:** Grad-CAM saliency map for OCT embedding 218. This embedding was the most strongly associated with baseline angina, baseline heart failure, future heart failure, and a large number of cerebral radiomic features in our Pearson correlation analysis. The second convolutional layer was targeted due to poor localisation in the third convolution. The embedding appears to localise to the retinal nerve fibre layer. Reproduced with the permission of UK Biobank.

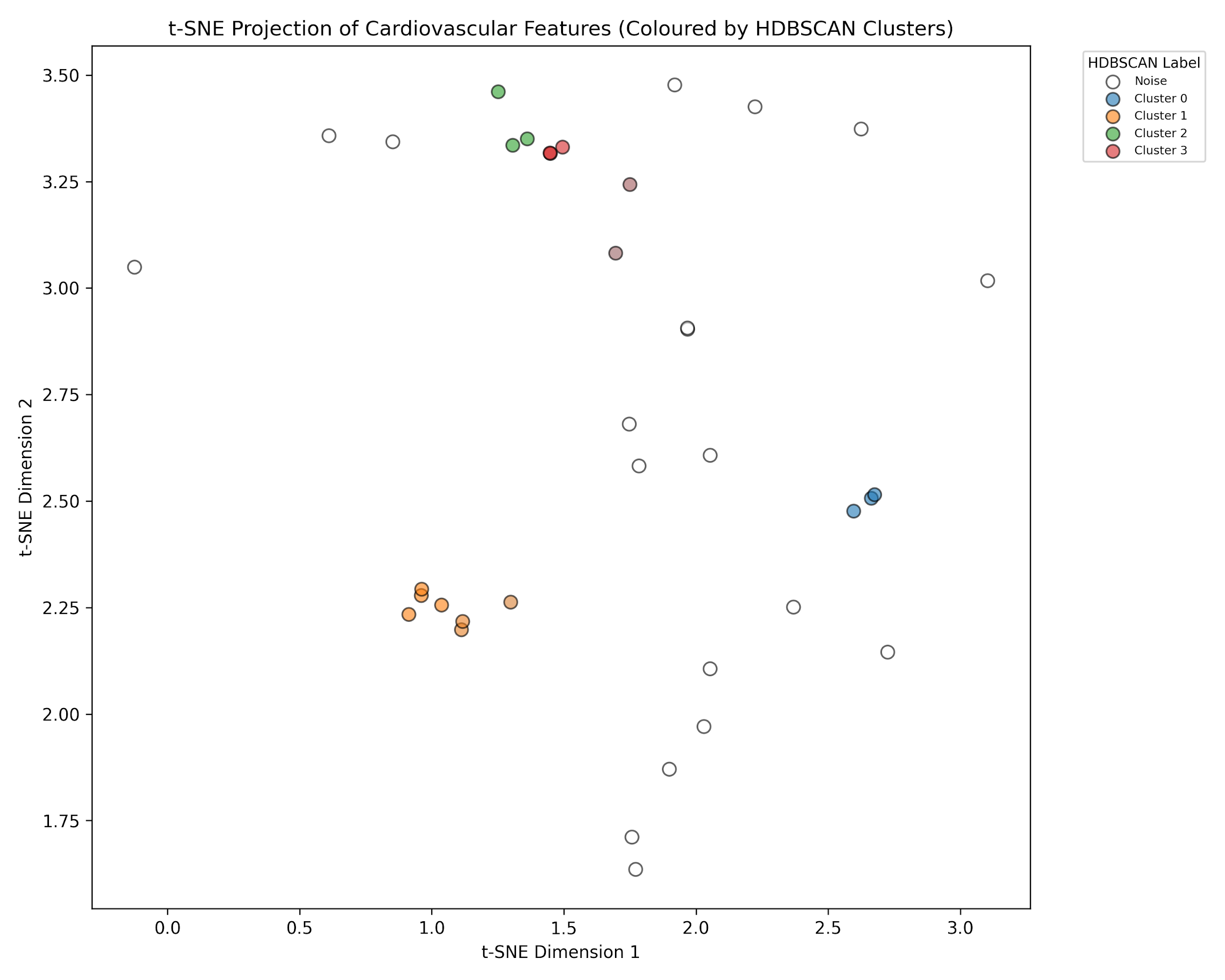

**Supplementary Figure 49:** A t-SNE projection of cardiovascular features clustered using HDBSCAN. The clusters have been color coded according to the HDBSCAN cluster

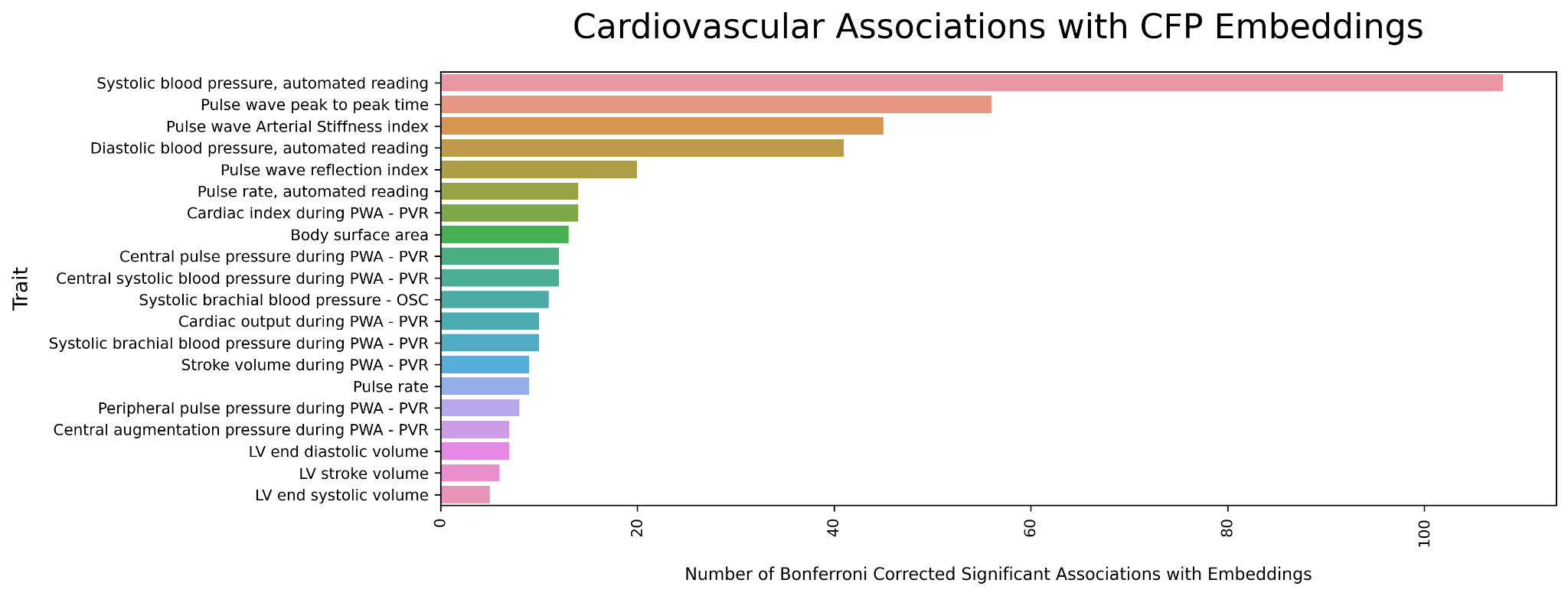

**Supplementary Figure 50**: A bar plot showing the top 20 cardiovascular traits in terms of the number of multiple testing corrective significant associations with CFP embeddings.

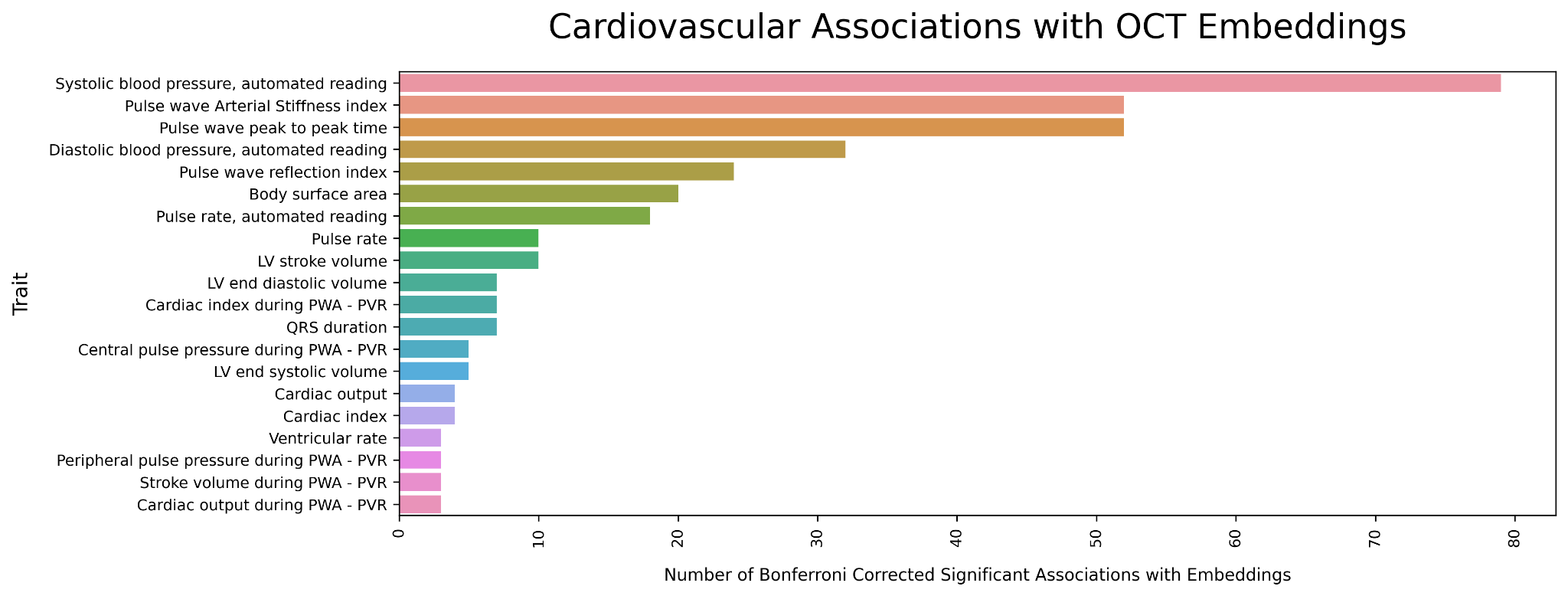
**Supplementary Figure 51**: A bar plot showing the top 20 cardiovascular traits in terms of the number of multiple testing corrective significant associations with OCT embeddings.

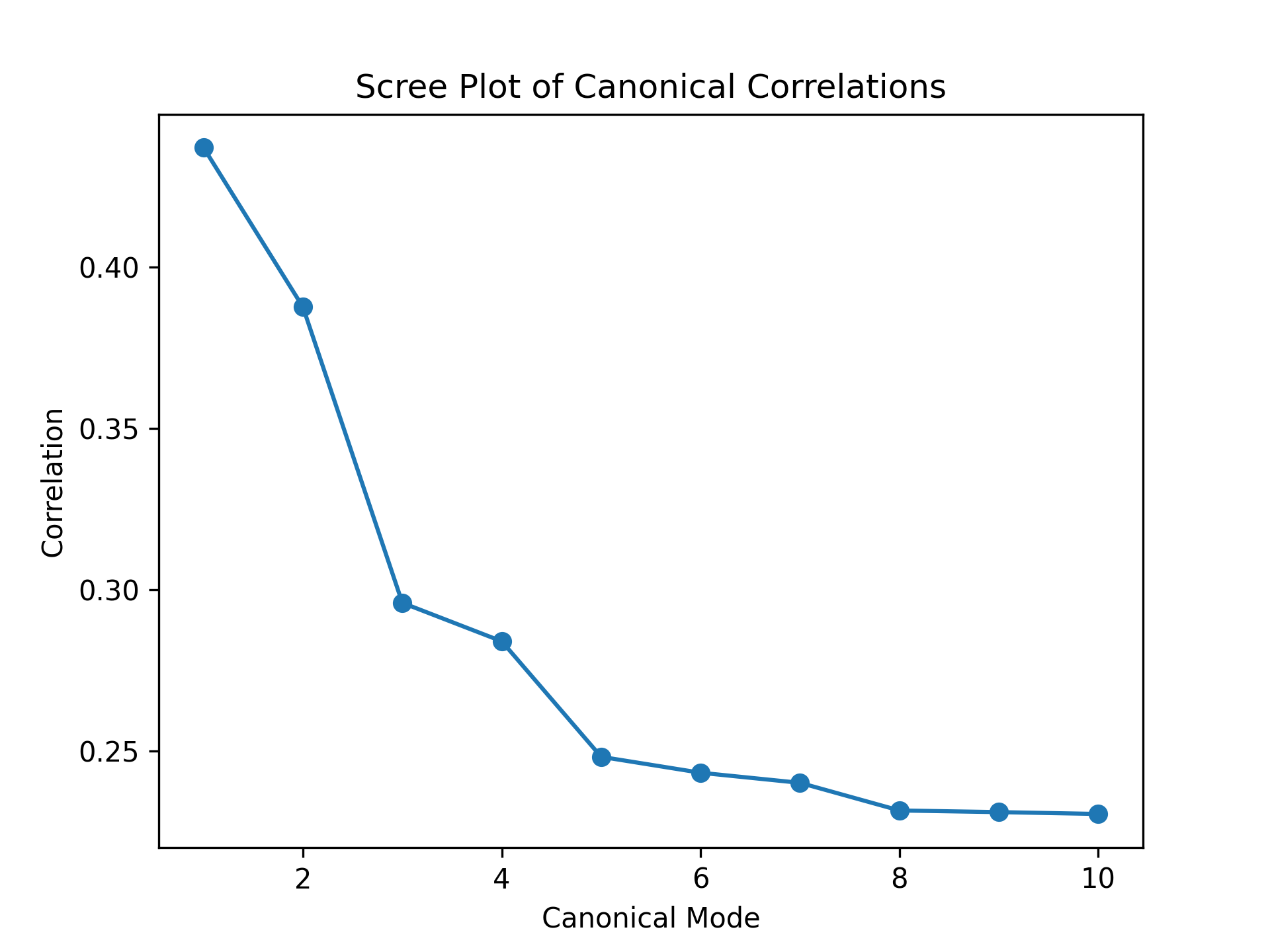

**Supplementary Figure 52:** A scree plot of the first 10 canonical modes in the CFP canonical correlation analysis.

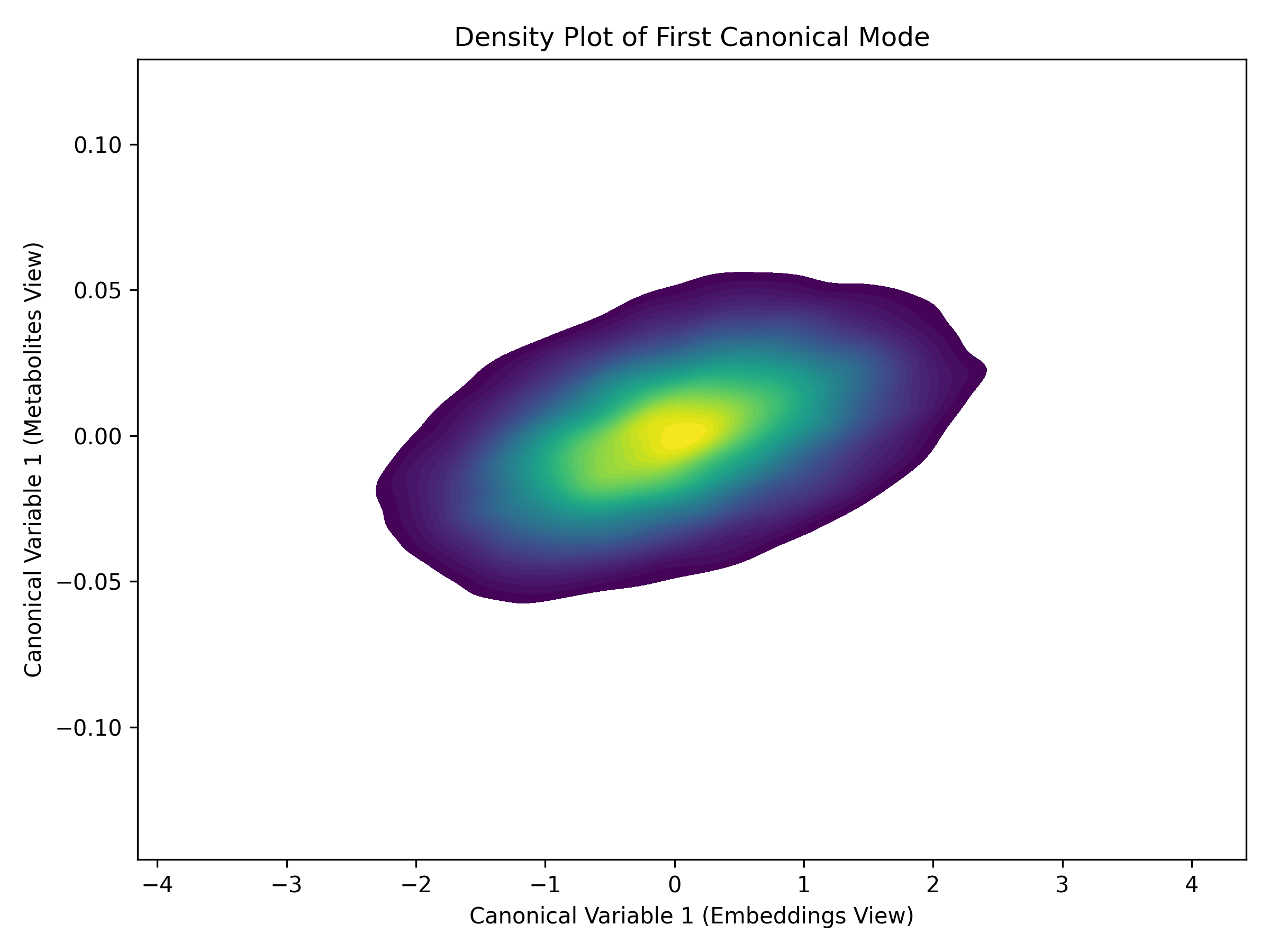

**Supplementary Figure 53:** A density plot of the first canonical mode in the CFP canonical correlation analysis, which is used to visualise the correlation between the canonical variables.

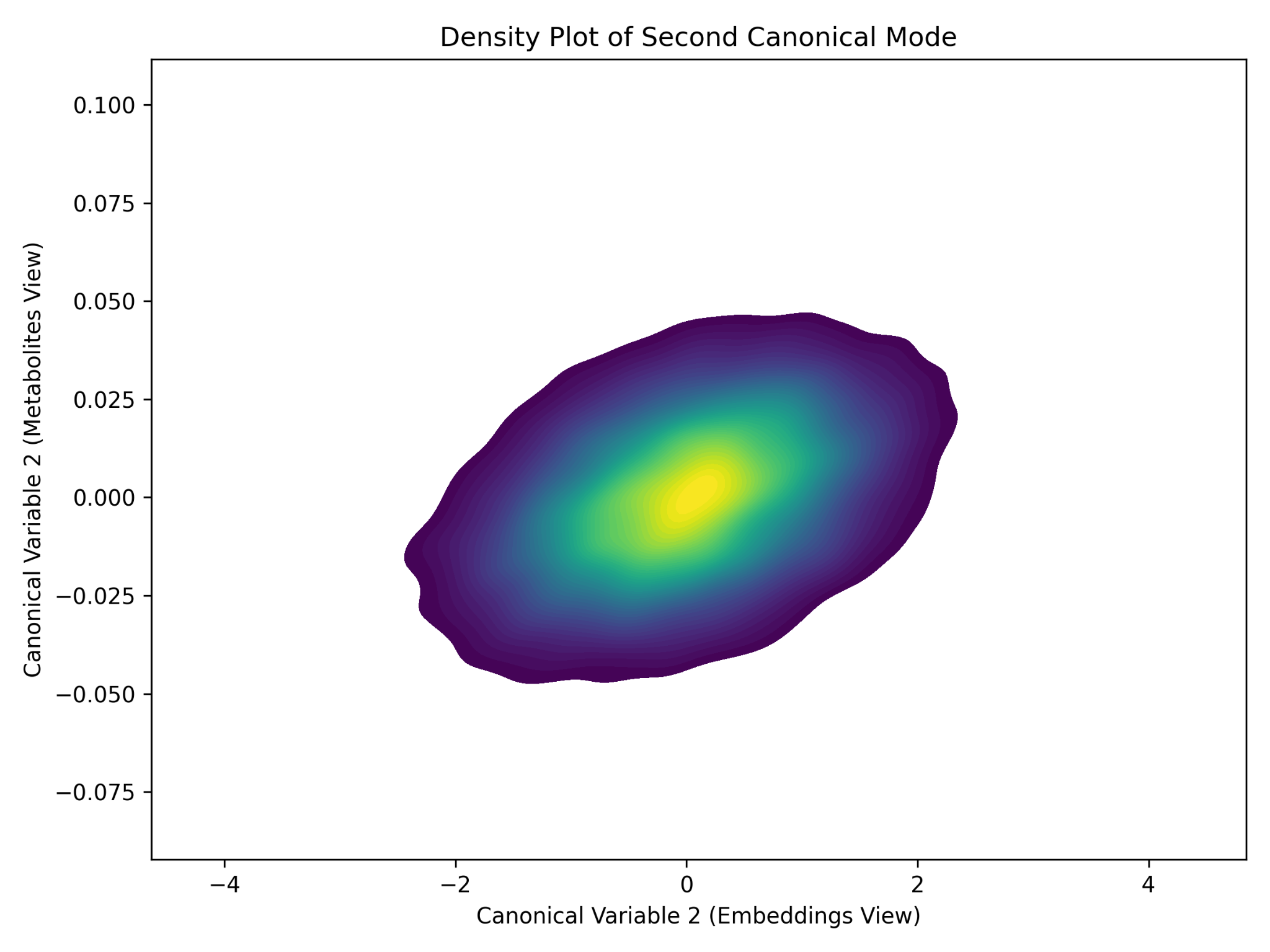

**Supplementary Figure 54:** A density plot of the second canonical mode in the CFP canonical correlation analysis, which is used to visualise the correlation between the canonical variables.

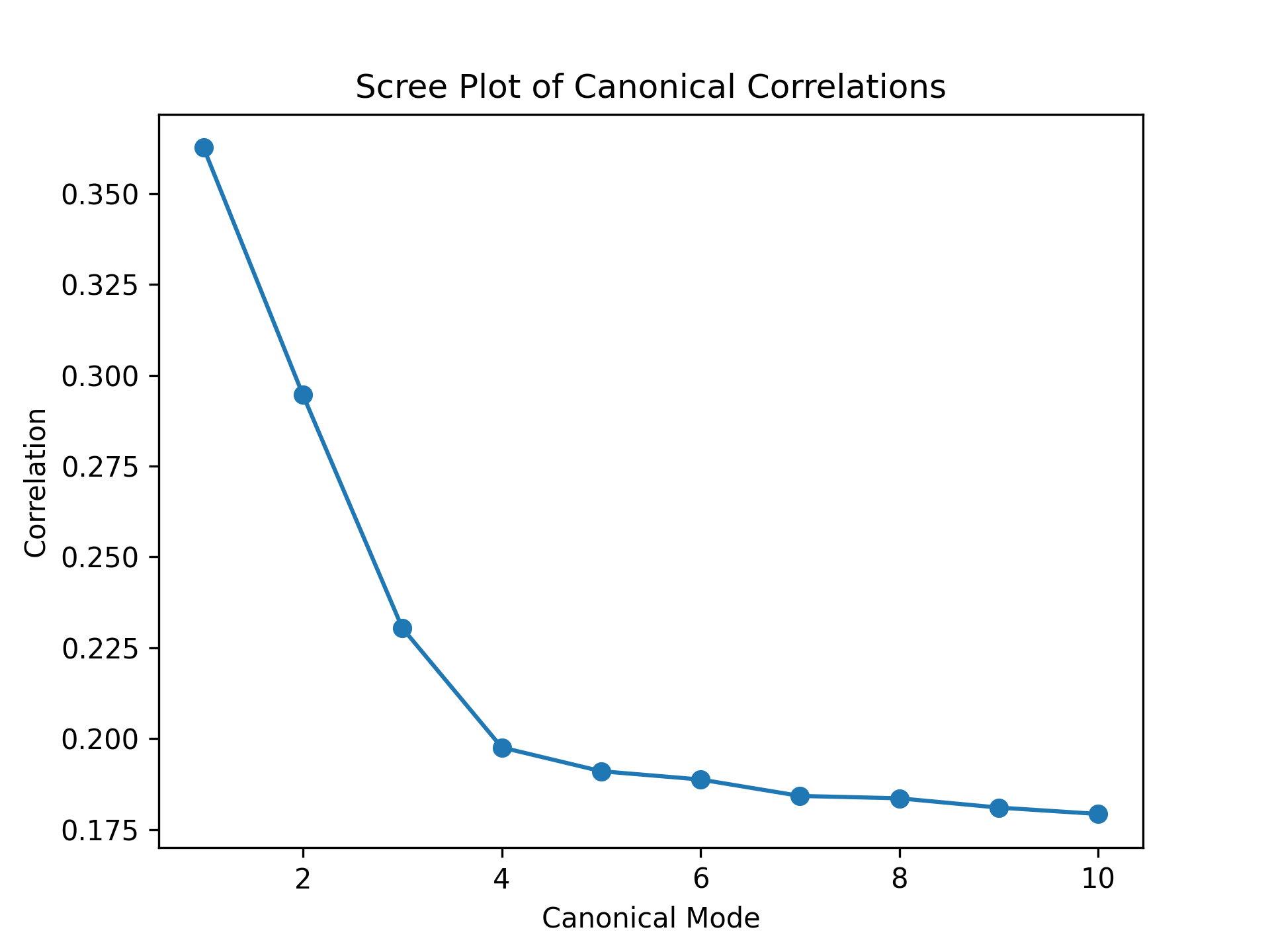

**Supplementary Figure 55:** A scree plot of the first 10 canonical modes in the OCT canonical correlation analysis.

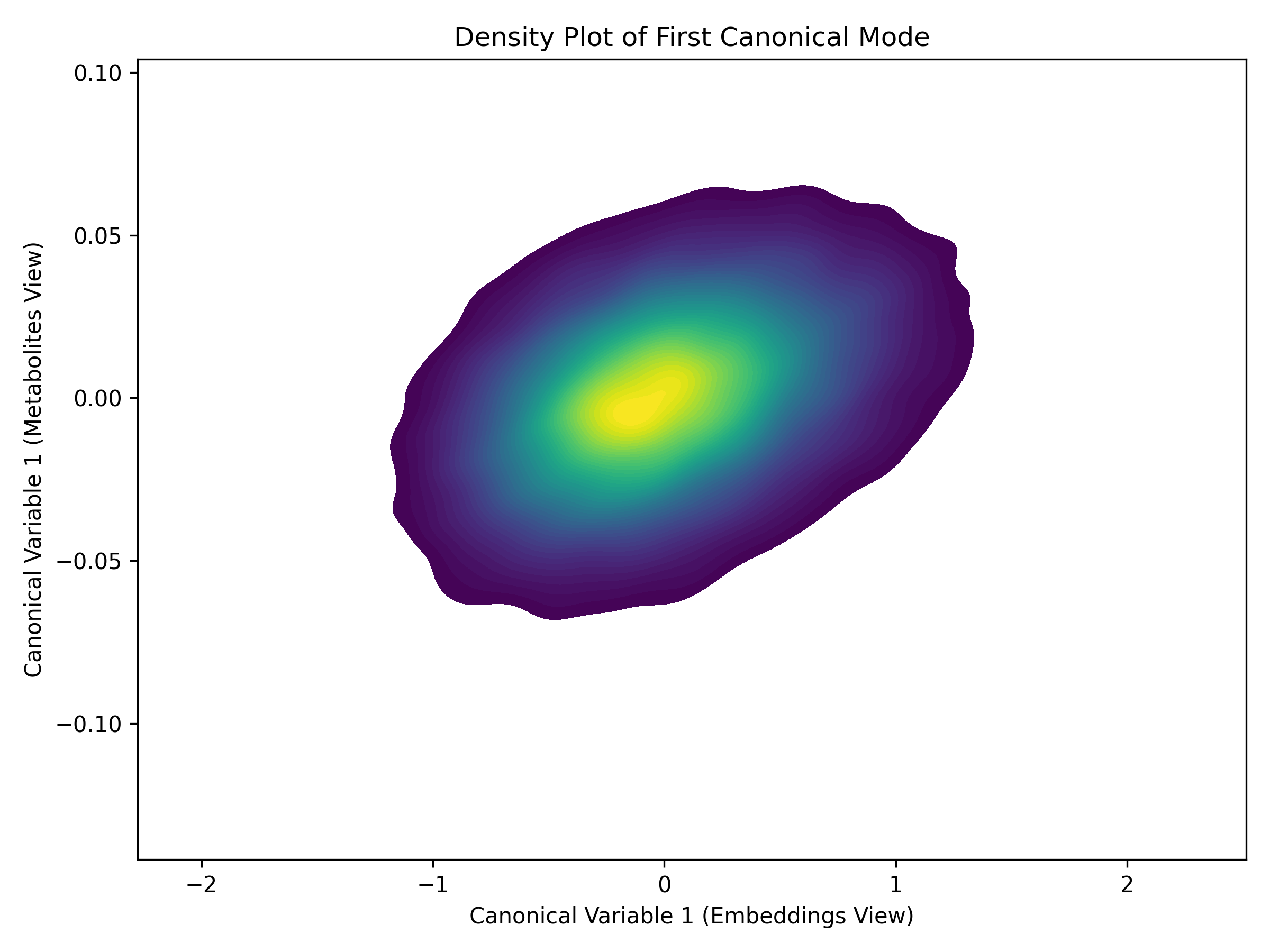

**Supplementary Figure 56:** A density plot of the second canonical mode in the OCT canonical correlation analysis, which is used to visualise the correlation between the canonical variables.

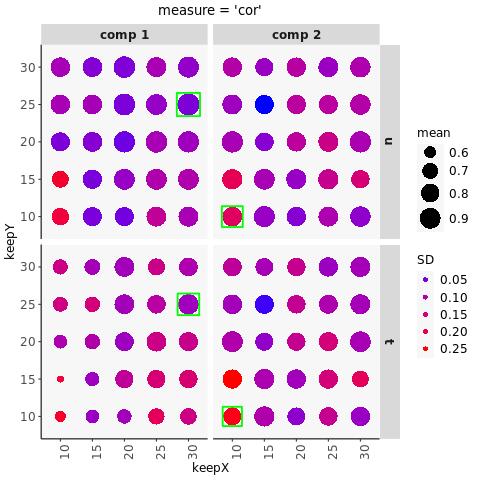

**Supplementary Figure 57:** The results of the colour fundus photograph – metabolite component number search. The green squares indicate the optimal number of components of to use for metabolites and embeddings in the component 1 and component 2 analysis.

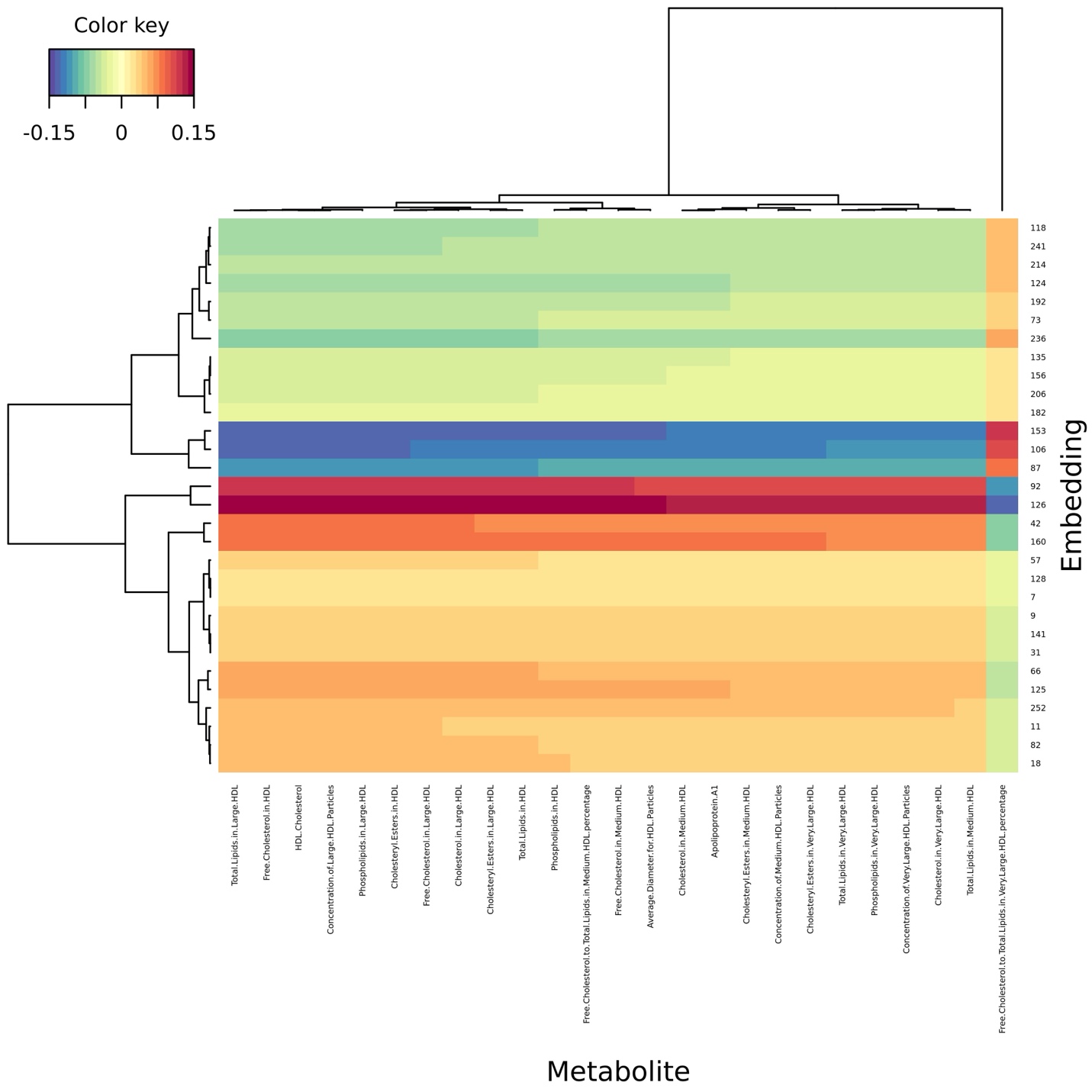
**Supplementary Figure 58**: A clustered heat map of the colour fundus photograph – metabolite sparse partial least squares analysis (SPLS). The plot features component one of the SPLS analysis and illustrates the correlation between features.

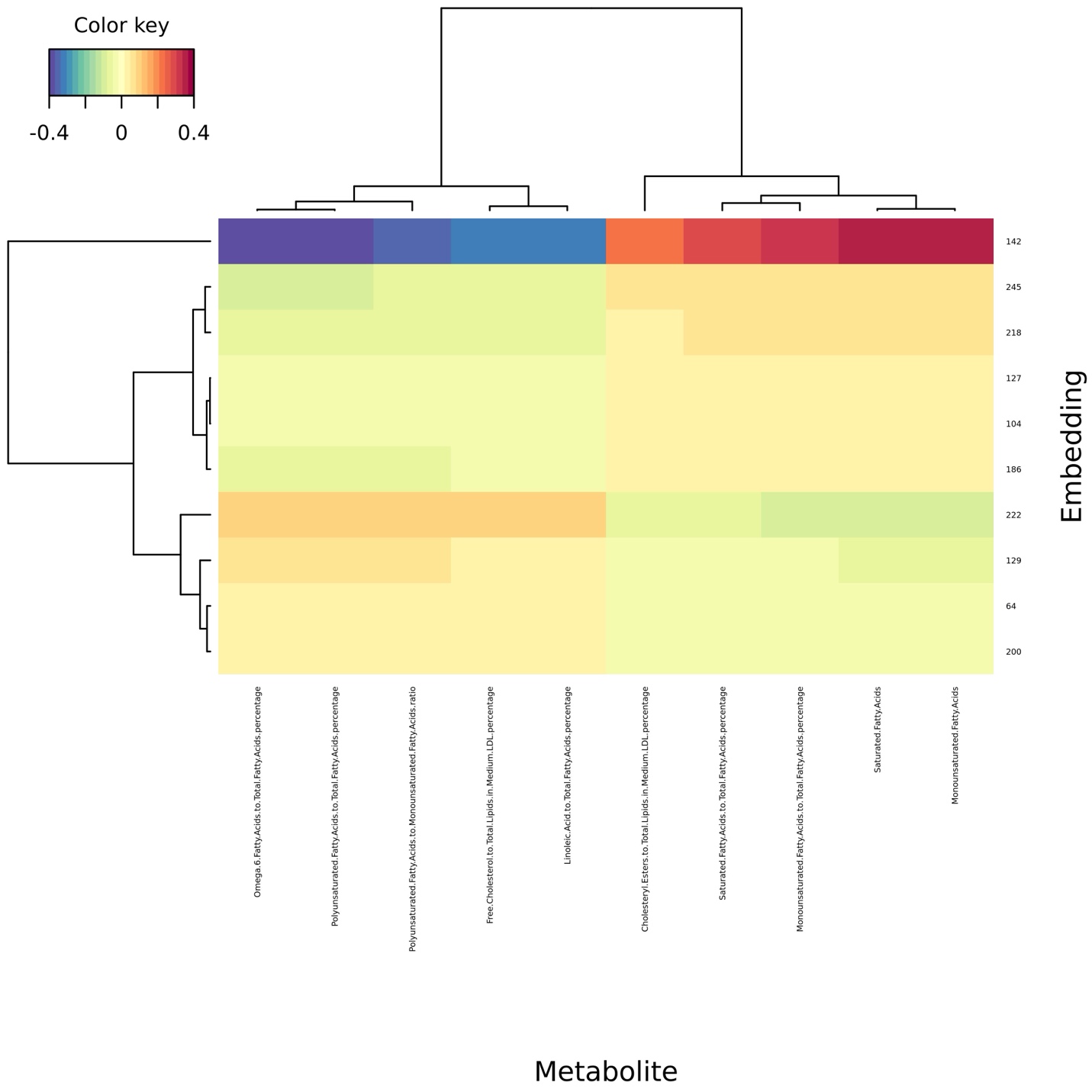
 **Supplementary Figure 59**: A clustered heat map of the colour fundus photograph – metabolite sparse partial least squares analysis (SPLS). The plot features component two of the SPLS analysis and illustrates the correlation between features.
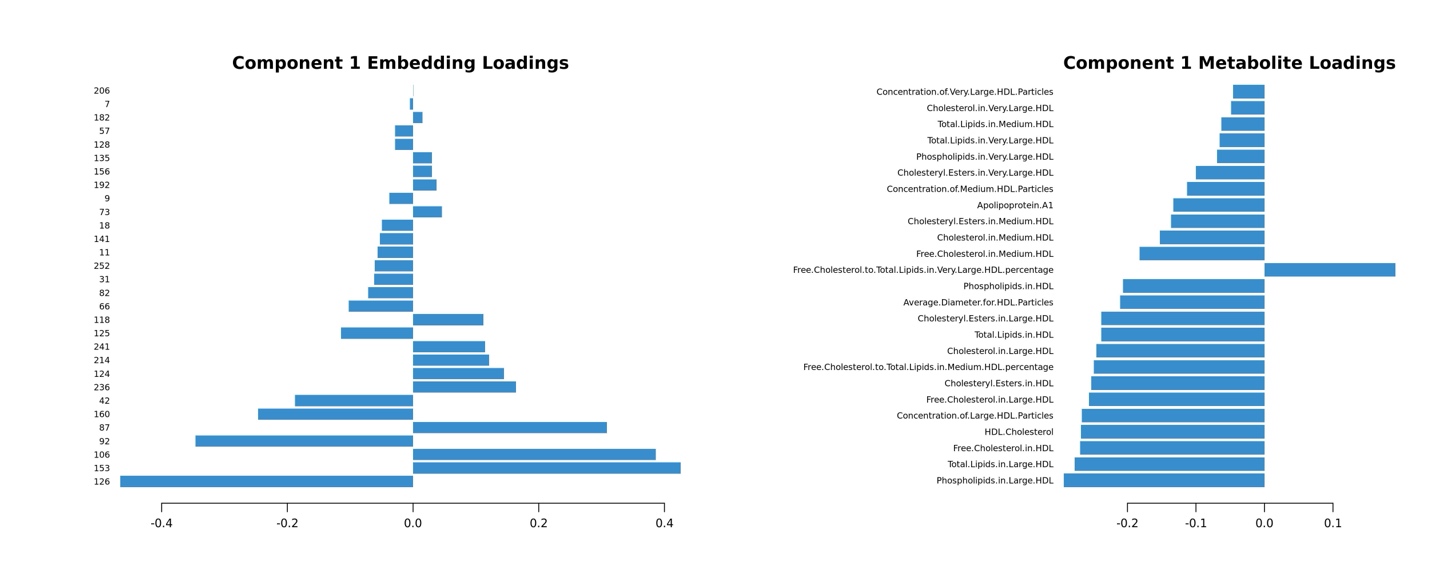

**Supplementary Figure 60:** A plot of each metabolite/embedding contribution to the latent components of the colour fundus photograph – metabolite sparse partial least squares analysis (SPLS). The plot shows which features drive embedding-metabolite associations in component 1.

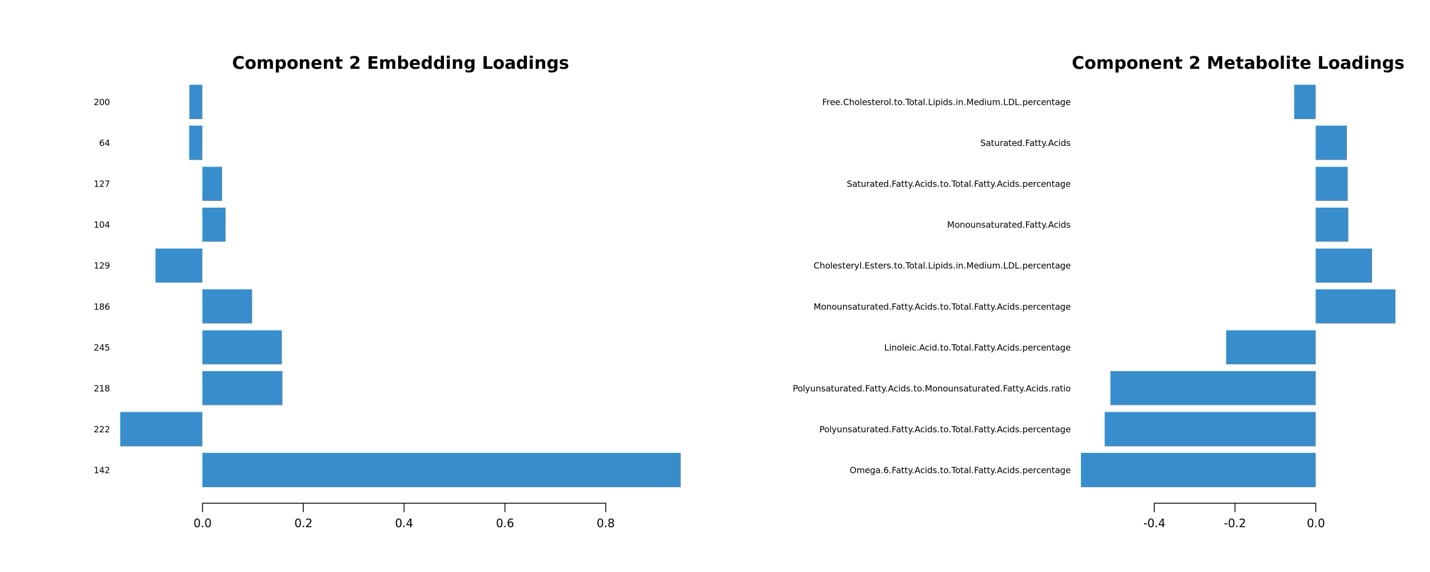

**Supplementary Figure 61:** A plot of each metabolite/embedding contribution to the latent components of the colour fundus photograph – metabolite sparse partial least squares analysis (SPLS). The plot shows which features drive embedding-metabolite associations in component 1.

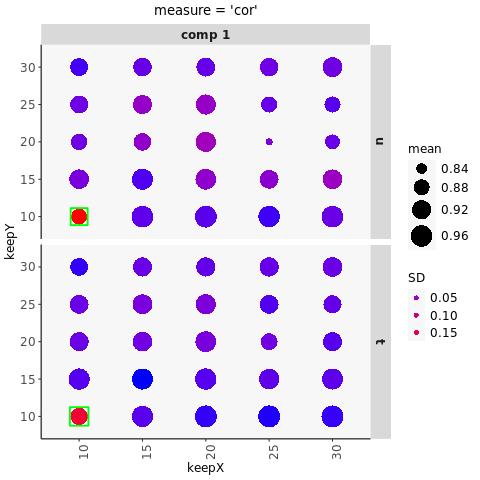

**Supplementary Figure 62:** The results of the optical coherence tomography – metabolite component number search. The green squares indicate the optimal number of components of to use for metabolites and embeddings in the component 1 analysis.

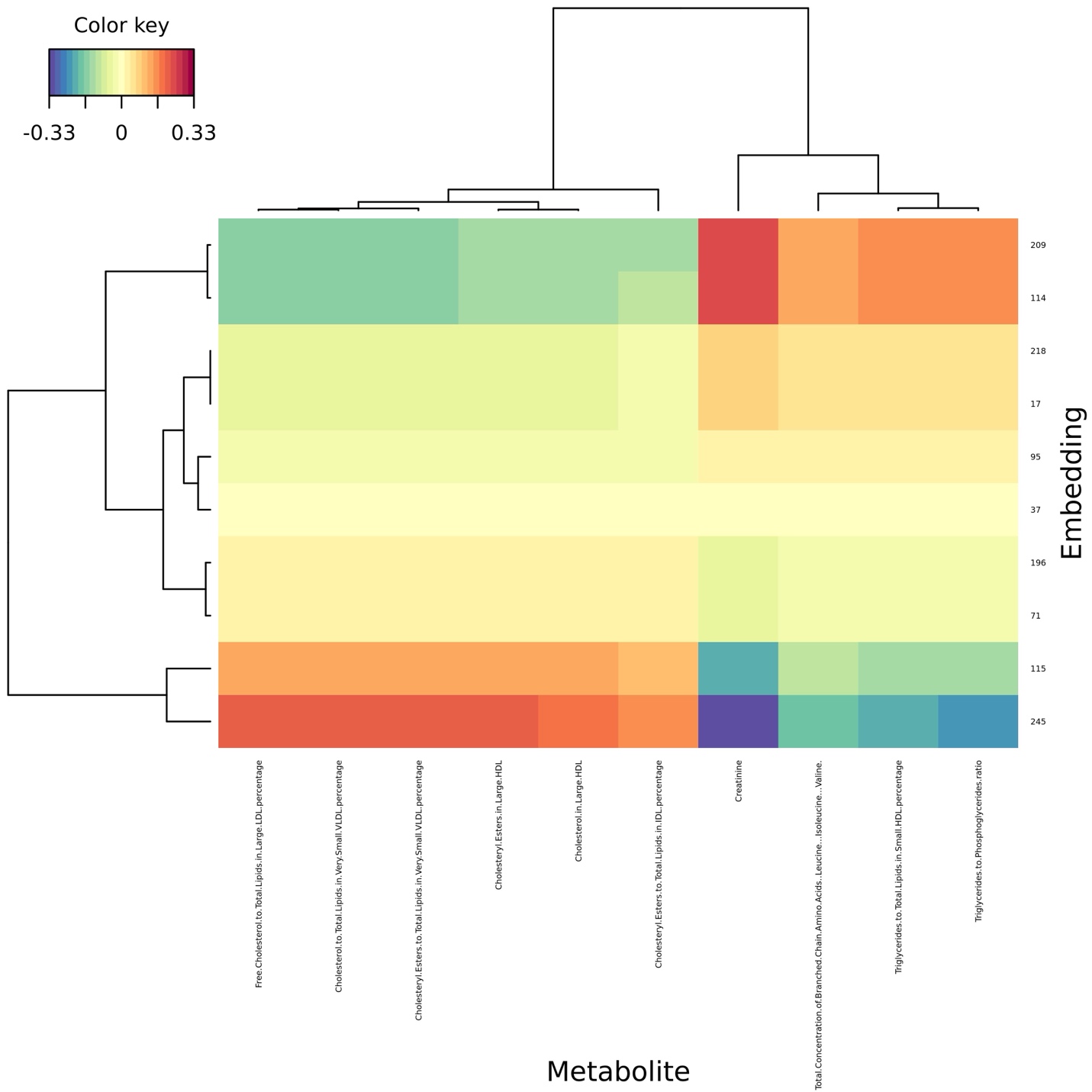
 **Supplementary Figure 63**: A clustered heat map of the optical coherence tomography – metabolite sparse partial least squares analysis (SPLS). The plot features component one of the SPLS analysis and illustrates the correlation between features.

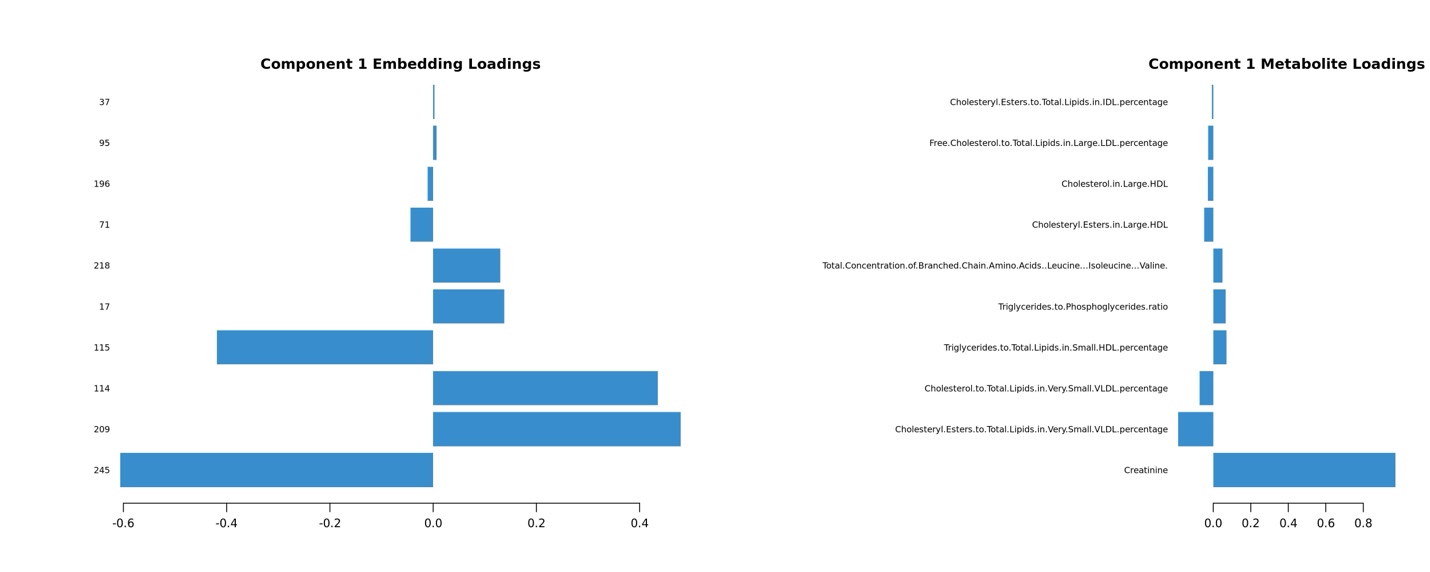

**Supplementary Figure 64:** A plot of each metabolite/embedding contribution to the latent components of the optical coherence tomography – metabolite sparse partial least squares analysis (SPLS). The plot shows which features drive embedding-metabolite associations in component 1.

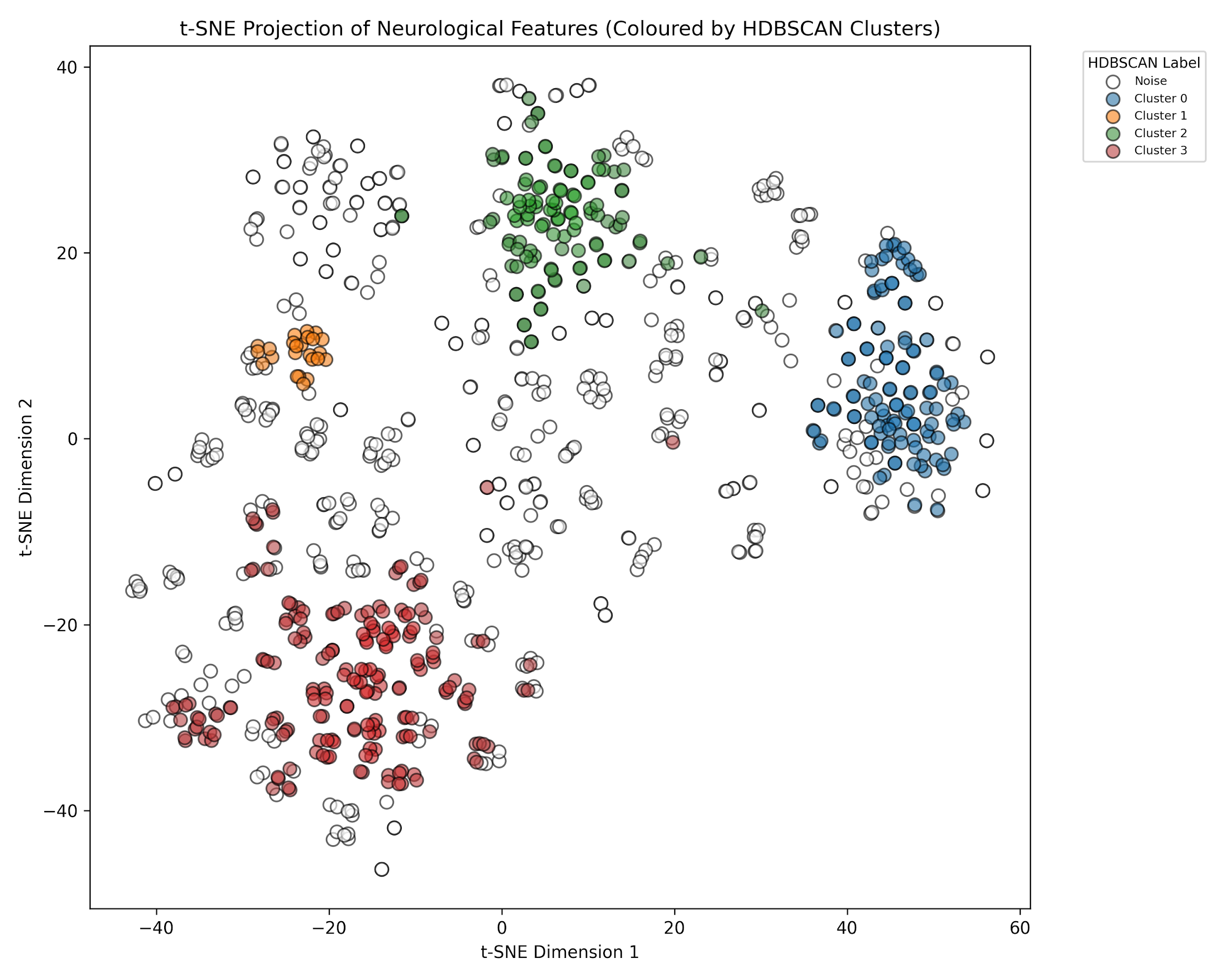

**Supplementary Figure 65:** A t-SNE projection of neurological features clustered using HDBSCAN. The clusters have been color coded according to the HDBSCAN cluster.

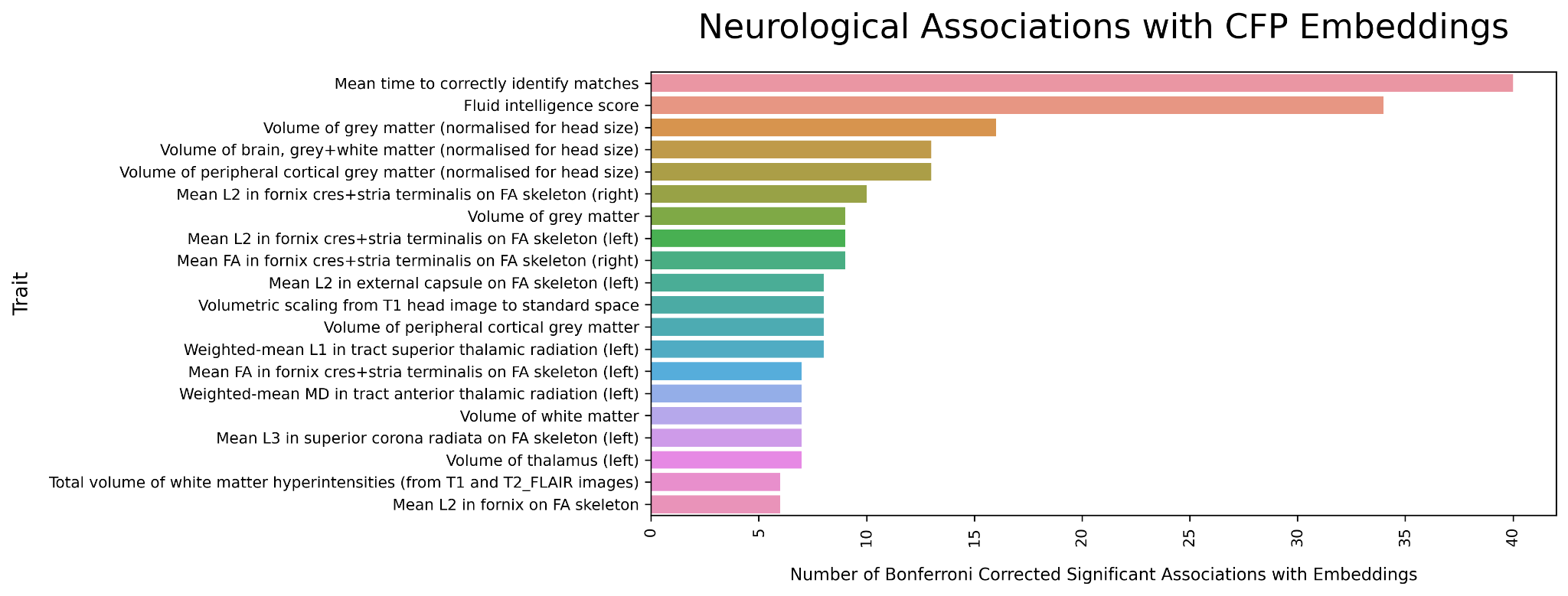

**Supplementary Figure 66**: A bar plot showing the top 20 neurological traits in terms of the number of multiple testing corrective significant associations with CFP embeddings.

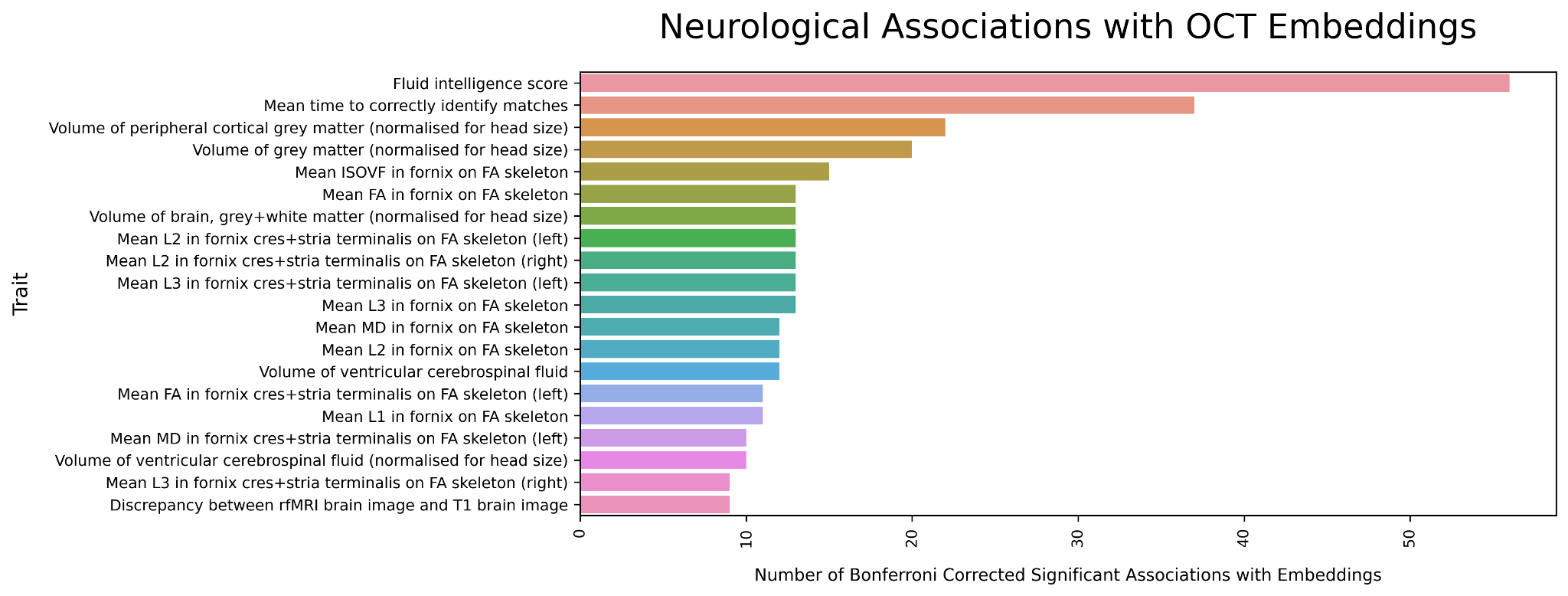

**Supplementary Figure 67**: A bar plot showing the top 20 neurological traits in terms of the number of multiple testing corrective significant associations with OCT embeddings.

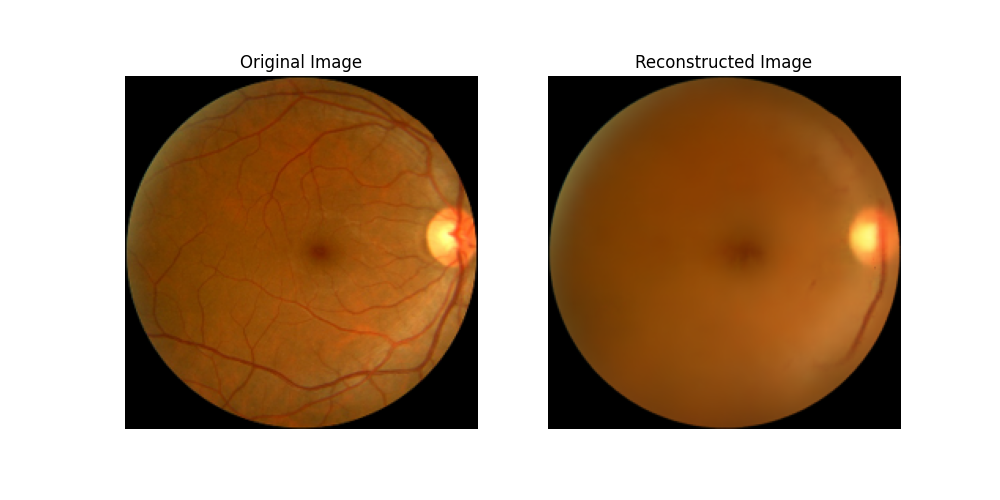
**Supplementary Figure 68:** An example of a CFP reconstruction prior to weighting of the vascular tree in our loss function. As can be seen, the model prioritised the background color over reconstruction of the vasculature, which is amongst the most biologically meaningful features. Reproduced with the permission of UK Biobank.

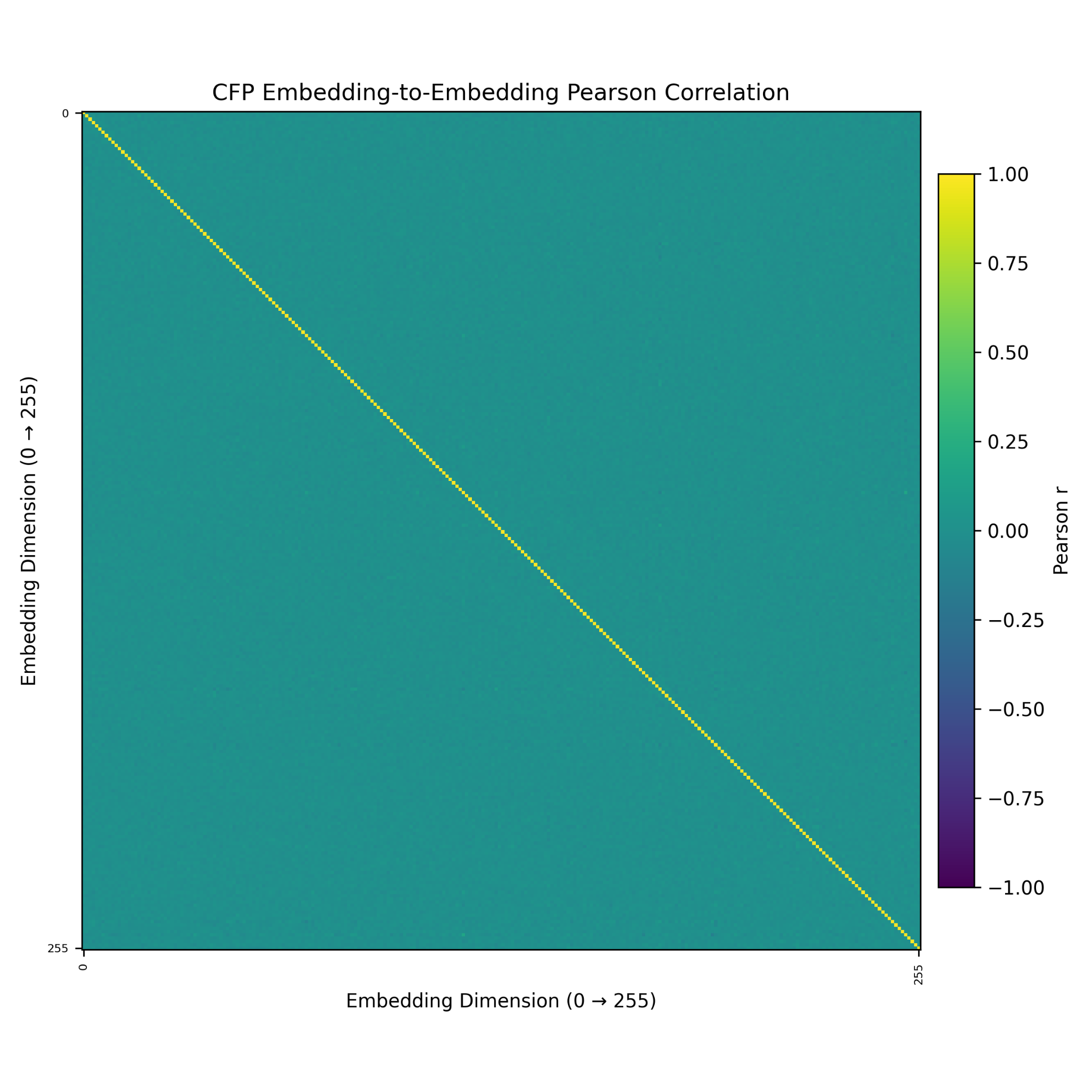
**Supplementary Figure 69:** A heat map of the correlations between CFP embeddings. As can be seen, there is perishingly little correlation between embeddings. This is an expected property given the distributional prior utilised.

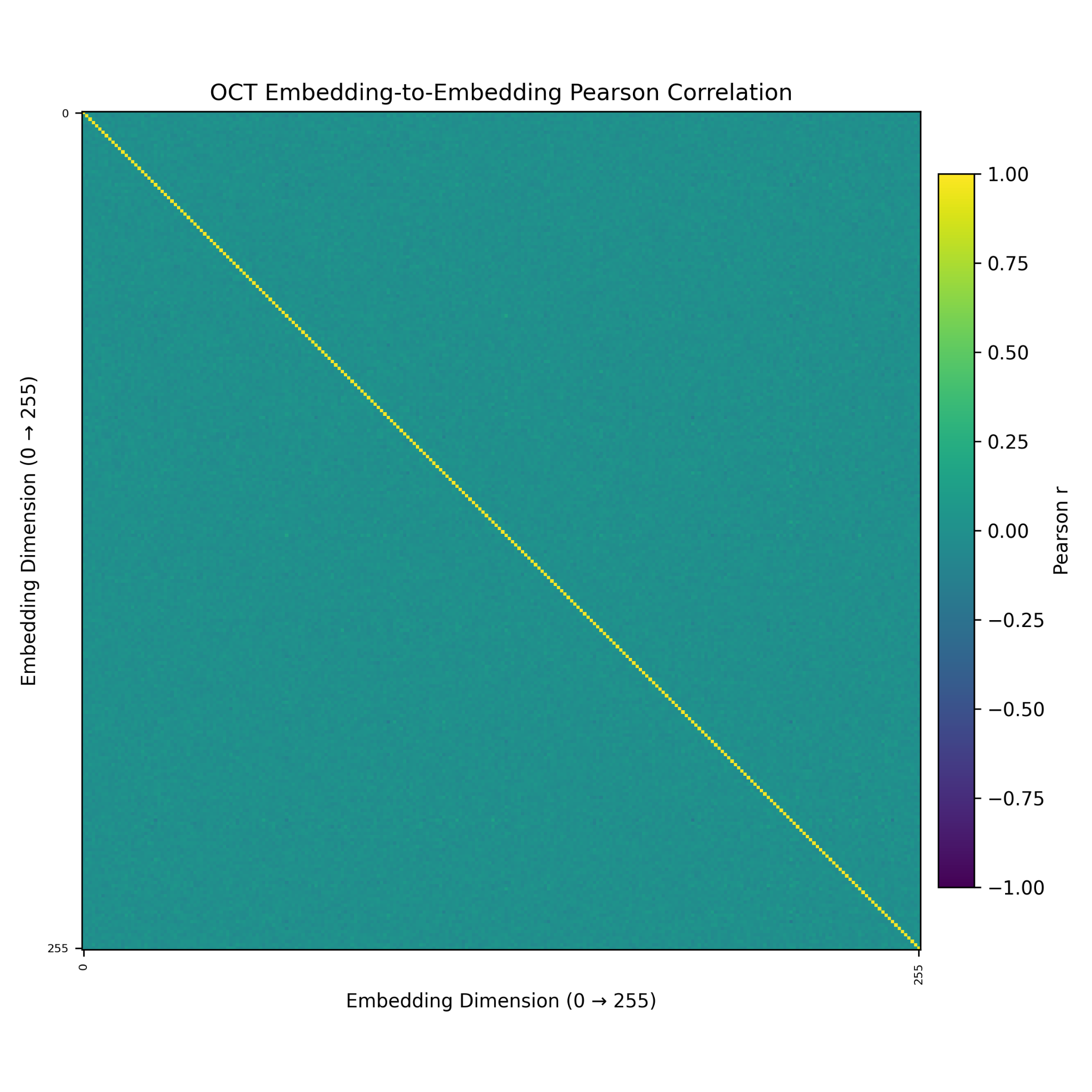
**Supplementary Figure 70:** A heat map of the correlations between OCT embeddings. As can be seen, there is perishingly little correlation between embeddings. This is an expected property given the distributional prior utilised.
